# Rethinking seasonal COVID-19 vaccination in the endemic era: summer timing, eligibility, and frequency in the United States

**DOI:** 10.64898/2026.09.23.26363810

**Authors:** Kate M. Bubar, Hailey J. Park, Samantha J. Bents, Sophia T. Tan, Benjamin J. Singer, J. Daniel Kelly, Tomás M. León, James Watt, Robert Schechter, Nathan C. Lo

## Abstract

**Importance:** With a recent shift towards summer seasonal predominance of COVID-19 across much of the US, debate continues over optimal COVID-19 vaccination strategies regarding timing (including summer vaccination), eligibility, and dosing frequency. We evaluated the population-level impact of alternative vaccine strategies to inform US and state vaccine policy.

**Objective:** To compare reductions in COVID-19 hospitalizations under different vaccination timing, eligibility, and dosing frequency.

**Design:** Decision analytical model using a stochastic individual-based model of SARS-CoV-2 transmission, calibrated to US COVID-19 hospitalization incidence from May 2024-April 2025.

**Setting:** Simulated US national population, with additional scenarios representing state-level seasonal patterns.

**Participants:** Simulated population of 5 million individuals with US age structure, stratified by risk group (immunocompetent, high risk with comorbid conditions, or immunocompromised).

**Exposures:** Vaccination timing, universal or age- and risk-based eligibility, and dosing frequency, including a hybrid strategy of universal annual vaccination with a second dose for adults ≥65 years and immunocompromised adults (US vaccine policy during the study period).

**Main Outcomes and Measures:** COVID-19 hospitalization risk per 100,000 persons by age-risk group, measured among vaccinated individuals and in the total population.

**Results:** If summer COVID-19 vaccination timing is feasible, annual universal vaccination in July minimized risk (441 [95% UI: 423-463] COVID-19 hospitalizations per 100,000 persons among adults ≥65 years) compared to a counterfactual scenario with no additional vaccination given during the study period (578 [553-601]). If timing does not change, hybrid vaccination minimized risk, followed by semiannual risk-based vaccination (460 [440-487] and 477 [454-499] hospitalizations per 100,000 in adults ≥65 years, respectively). The advantage of summer timing diminished if vaccine coverage decreased by 5-10 absolute percentage points (25-50% relative decrease). The magnitude of benefit from summer-timed vaccination and semiannual doses in the highest-risk groups varied regionally.

**Conclusions and Relevance:** Although annual COVID-19 vaccination has historically been timed for winter transmission, summer vaccination may avert more hospitalizations if coverage is maintained, especially in states with predominantly summer transmission including Western and Southern states. The comparative impact of vaccination strategies, especially the benefit of a semiannual dose in the highest-risk groups, may vary regionally. COVID-19 vaccination timing and eligibility may warrant reconsideration.

**Key points:** *Question:* Given increasing summer predominance of COVID-19 in the US, can changing the timing, eligibility, or dosing frequency of vaccination further reduce hospitalizations?

*Findings:* In a decision analytical model calibrated to US COVID-19 hospitalization incidence, annual universal vaccination in July reduced hospitalization risk in adults ≥65 years more than recently recommended hybrid vaccination (universal vaccination with a second dose in the highest-risk groups) with historical fall timing, provided historical vaccine coverage was maintained. The benefit from summer-timed and semiannual vaccination in the highest-risk groups varied regionally.

*Meaning:* Shifting COVID-19 vaccination to summer could further reduce hospitalizations, if coverage is maintained.

## Introduction

COVID-19 continues to cause substantial disease burden in the United States, with an estimated 300,000 to 450,000 hospitalizations and 35,000 to 50,000 deaths from July 2024 through June 2025^1^. Hospitalizations and deaths remain concentrated among adults aged ≥65 years, immunocompromised individuals, and those with underlying medical conditions^2–4^.

During the 2024-25 season, US recommendations included annual COVID-19 vaccination for all individuals aged ≥6 months, with a second dose (i.e., semiannual vaccination) for adults ≥65 years and immunocompromised individuals^5,6^. National recommendations have since become less unified, with major medical societies issuing independent recommendations for the 2026-27 season^7–9^. Historically, production cycles, regulatory review, and national recommendations have concentrated vaccination in the fall to provide peak protection during anticipated winter transmission, in alignment with longstanding seasonal influenza immunization. However, both optimal vaccination timing and the benefit of semiannual doses depend on when SARS-CoV-2 transmission peaks during the year and the durability of vaccine-derived immunity.

Since 2020, COVID-19 has exhibited a semiannual seasonal pattern in the US with distinct summer and winter waves^3^. While winter waves were predominant through the 2023-24 season, summer hospitalization peaks exceeded the winter peaks in the 2024-25 and 2025-26 seasons nationally, though the relative magnitude varies by region^10,11^. This contrasts sharply with historical vaccine uptake occurring predominantly September through November. Given that vaccine-derived immunity wanes within 6 months^12–14^, fall vaccination provides diminished protection during summer peaks, while earlier vaccination protection may wane before winter ends. This pattern of two annual peaks raises two critical policy questions: whether annual vaccination should be retimed to align with evolving seasonal patterns, and, if timing remains unchanged, whether to continue with semiannual vaccination in the highest-risk groups.

Previous modeling studies have evaluated some of these questions, quantifying benefits of semiannual vaccination^15–17^, vaccination timing relative to winter seasonal peaks^17–19^, and optimizing dose intervals in older adults and immunocompromised individuals^18^. However, the comparative benefits of vaccine timing, particularly summer vaccination, eligibility, and dose frequency remain to be evaluated to inform ongoing policy in the US, given recent shifts towards summer seasonal predominance of COVID-19 in some parts of the US. We therefore developed an age- and risk-stratified transmission model to determine which of these policy dimensions offers the greatest reduction in COVID-19 hospitalizations.

## Methods

### Data sources

We obtained data on national- and state-level age-stratified weekly COVID-19 hospitalization incidence from COVID-NET^3^ (Figure 1A, Supplementary Section S1). We estimated the proportion of observed COVID-19 hospitalizations occurring in individuals with underlying medical conditions and immunocompromising conditions based on literature estimates^20,21^. Population demographics matched the national US age distribution and published risk group prevalence (Table S1).

**Figure 1:**
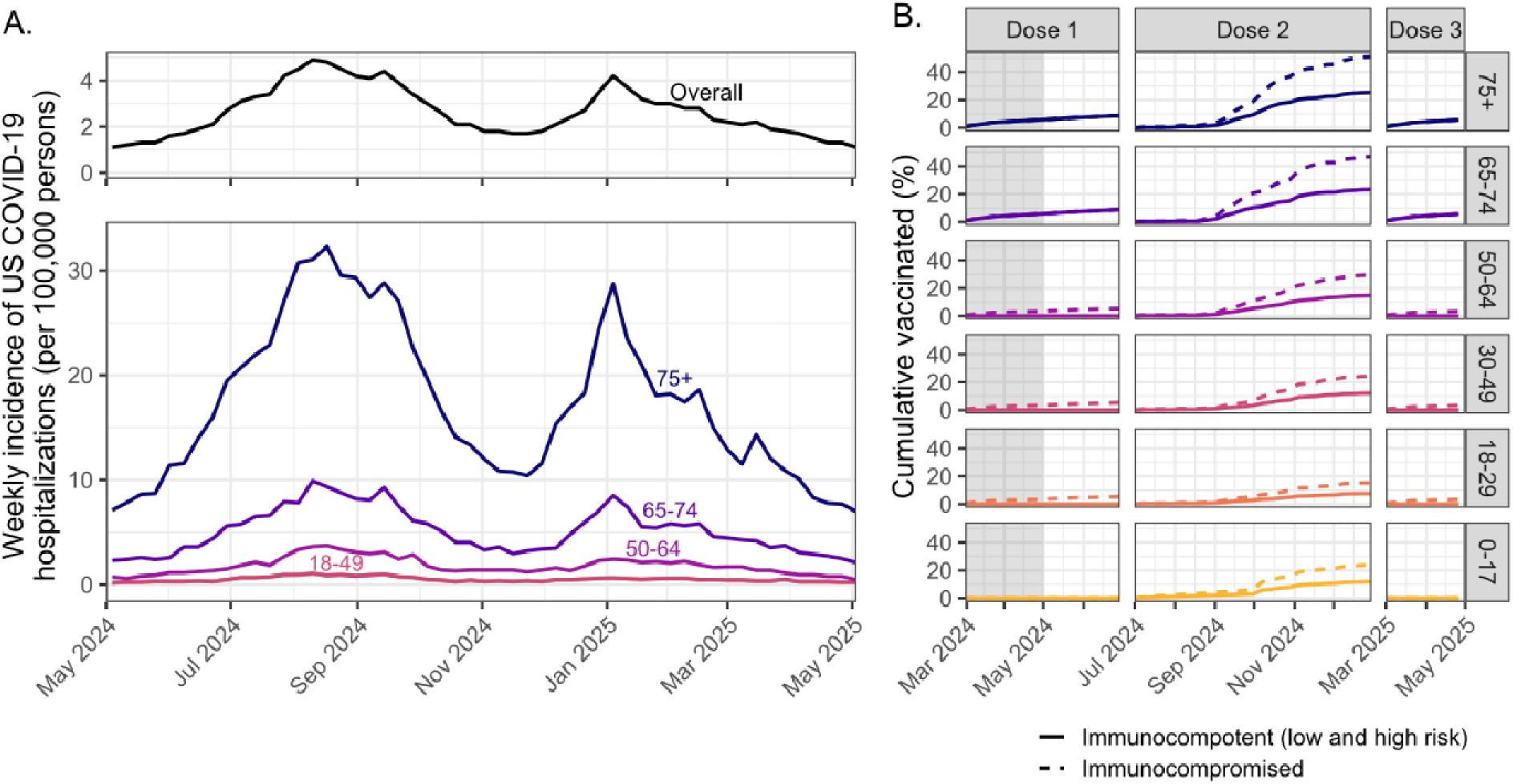
COVID-19 hospitalization incidence and vaccine uptake vary substantially by age. (A) Observed weekly total (top) and age-stratified (bottom) COVID-19 hospitalizations per 100,000 persons in the US (May 2024–April 2025)^3^ (B) Estimated historical COVID-19 vaccine uptake (March 2024-April 2025) by age group (color) and risk status (line style: solid indicates low and high risk group, dashed indicates immunocompromised risk group) used to parameterize historical coverage scenarios. Doses 1 and 3: spring vaccination in those eligible for semiannual dose, Dose 2: fall annual dose.

We defined vaccine coverage as the proportion vaccinated and uptake as the rate at which individuals are vaccinated. We modeled the 2023-24 and 2024-25 COVID-19 vaccine formulations using CDC data on national age-specific vaccine coverage and uptake patterns^22–24^, estimating risk-stratified coverage from published data^25^ (Figure 1B, Table S2). We included protection against infection and hospitalization by modeling protection that wanes over time using linear mixed-effects models as in our prior work^16^. We distinguished between vaccine-acquired, infection-acquired, and hybrid immunity for individuals with both prior infection and vaccination. Protection was parameterized using predominately post-Omicron era studies (2022-2024; Table S3)^26–28^. We validated model-predicted relative vaccine effectiveness against recently observed data for the 2023-24 and 2024-25 formulations (Supplementary Section S2, Figures S1-S2).

### Microsimulation model

We simulated SARS-CoV-2 transmission and COVID-19 hospitalizations in the US from May 2024 through April 2025, using a stochastic individual-based dynamic transmission model, adapted from our prior work^16^. This study period represents the post-Omicron era and contemporary epidemiological patterns for SARS-CoV-2. We modeled a population of 5 million individuals, providing adequate sample sizes for each subgroup while maintaining computational feasibility. We considered three risk groups: immunocompetent (hereafter referred to as low risk), immunocompetent persons with comorbid conditions associated with higher risk of COVID-19 disease^21^ (hereafter referred to as high risk), and immunocompromised (i.e., those with moderate/severe immunocompromising conditions; Supplementary Section S1). Age and risk group assignment was fixed throughout the simulation.

Each day, individuals’ probabilities of SARS-CoV-2 infection or COVID-19 hospitalization were modeled as Bernoulli trials. Infection risk depended on a time-varying force of infection and individual immunity.

Using approximate Bayesian computation with a sequential Monte Carlo algorithm (ABC-SMC), we calibrated monthly transmission coefficients and a single, time-invariant severity parameter to match observed age-specific weekly COVID-19 hospitalization incidence in adults ≥18 years. We retained parameter sets that met a goodness-of-fit criterion on cumulative age-specific incidence and assessed model validity against age-risk incidence and literature estimates not used in calibration (Supplementary Section S3).

### Study outcomes

We aggregated simulated hospitalizations to estimate annual COVID-19 hospitalization incidence per 100,000 persons over the 12 month study period in four reporting groups: adults 18-64 years who are low risk, high risk, or immunocompromised, and all adults ≥65 years, for whom recommendations do not differ by risk status. We report incidence in the total population and among the vaccinated (those who received a vaccine during the study period). Relative to no additional vaccination during the study period, we estimated absolute and relative risk reduction, the number needed to vaccinate to prevent one hospitalization, and workdays missed due to COVID-19 illness or vaccine side effects (Supplementary Section S4).

### Vaccination scenarios

To compare vaccination timing, we considered annual universal vaccination (available to all individuals ≥6 months, in line with vaccine recommendations during the study period without the semiannual dose) with historical vaccine coverage, varying uptake start dates from May to November and the rate of uptake (compressed in one month and gradual over approximately 4 months, matching historical patterns; Figure 1B). We also considered the timing of vaccination under two seasonality archetypes defined by the ratio of cumulative hospitalization burden in May-October versus November-April: (1) a predominant summer/fall peak with a minimal winter peak, resembling patterns observed in California and Oregon, and (2) two annual peaks with higher winter burden, resembling Michigan and Connecticut. These archetypes represent the extremes of this ratio observed among the 12 COVID-NET states with complete data (range of summer / winter ratio: 0.83 to 3.40), with the US-wide model falling between them (ratio = 1.28; Supplementary Section S5). Regional heterogeneity in COVID-19 seasonality beyond COVID-NET catchment areas is consistent with the patterns captured by these archetypes^32,33^.

To compare eligibility and dosing frequency under historical vaccine timing, we evaluated five main strategies: (1) annual universal vaccination (available to all individuals ≥6 months); (2) annual risk-based vaccination in adults ≥65 years and immunocompromised individuals; (3) annual risk-based vaccination, expanded to include higher-risk adults with comorbid conditions; (4) semiannual risk-based vaccination for the same groups as (2); and (5) a hybrid vaccination strategy combining (1) and (4) (Figure 4A). Three additional strategies, which varied the universal age threshold (≥18 years) and applied semiannual dosing to expanded risk groups, are included in the Supplement (Table S4).

### Sensitivity analysis and uncertainty

We conducted sensitivity analyses on waning immunity, vaccine coverage (seasonal variation and a higher-coverage scenario), and alternative evaluation periods (i.e., cumulative incidence six months after vaccination began; Supplementary Section S6). Reported uncertainty intervals are the 2.5th–97.5th percentile range across these iterations, capturing variation across the calibrated parameters and stochastic variation across model runs.

## Results

### Description of COVID-19 burden and vaccination patterns in the US

During the study period from May 2024 to April 2025, COVID-19 hospitalizations in the US exhibited dual seasonal peaks (summer/fall: July-October; winter: December-March), consistent with 2023-24 and 2025-26 patterns (Figures 1A, S3). Nationally, the cumulative hospitalization incidence and peak hospitalization rate in summer 2024 exceeded winter 2024-2025, though this varied by state (Figure S4). Weekly COVID-19 hospitalization incidence increased substantially with age, with weekly minimum-maximum ranging from 0.2-1.1 per 100,000 in adults 18-29 years to 7.1-32.3 per 100,000 in adults ≥75 years (Figure 1A).

Estimated historical vaccine coverage during the 2024-2025 season also increased with age and was higher among immunocompromised individuals (Figure 1B). Estimated fall dose coverage ranged from 11% (adults 18-29 years) to 38% (adults ≥75 years). Coverage of the spring semiannual dose among eligible populations (immunocompromised and adults ≥65 years) was low (5.4-8.9%; Figure 1B, Table S2).

### Optimal timing of annual vaccination

Vaccination in the summer months (June-August) minimized hospitalization risk across all age and risk groups. For example, among vaccinated adults ≥65 years, July vaccination reduced COVID-19 hospitalization risk the most, averting 224 [95% UI: 200-248] hospitalizations per 100,000 persons, followed by vaccination in June (211 [184-235]) and August (206 [176-239]; Figure 2, Table S5). In contrast, September vaccination, which approximates historical timing, had considerably less benefit (179 [150-207] averted COVID-19 hospitalizations per 100,000 persons). These reductions are relative to a counterfactual with no additional vaccination, under which annual hospitalization risk among all adults ≥65 years was 578 [553–601] COVID-19 hospitalizations per 100,000 persons (Table 1). This ranking persisted across all other age-risk groups and the total population (including those not vaccinated during the study period), with July performing better than each other month in a majority of paired simulations, although absolute differences in other groups were often smaller than in adults ≥65 years (Table S6). Summer vaccination remains advantageous under four-month vaccination uptake, which matched historical uptake patterns (Figure S5, Table S7).

**Figure 2:**
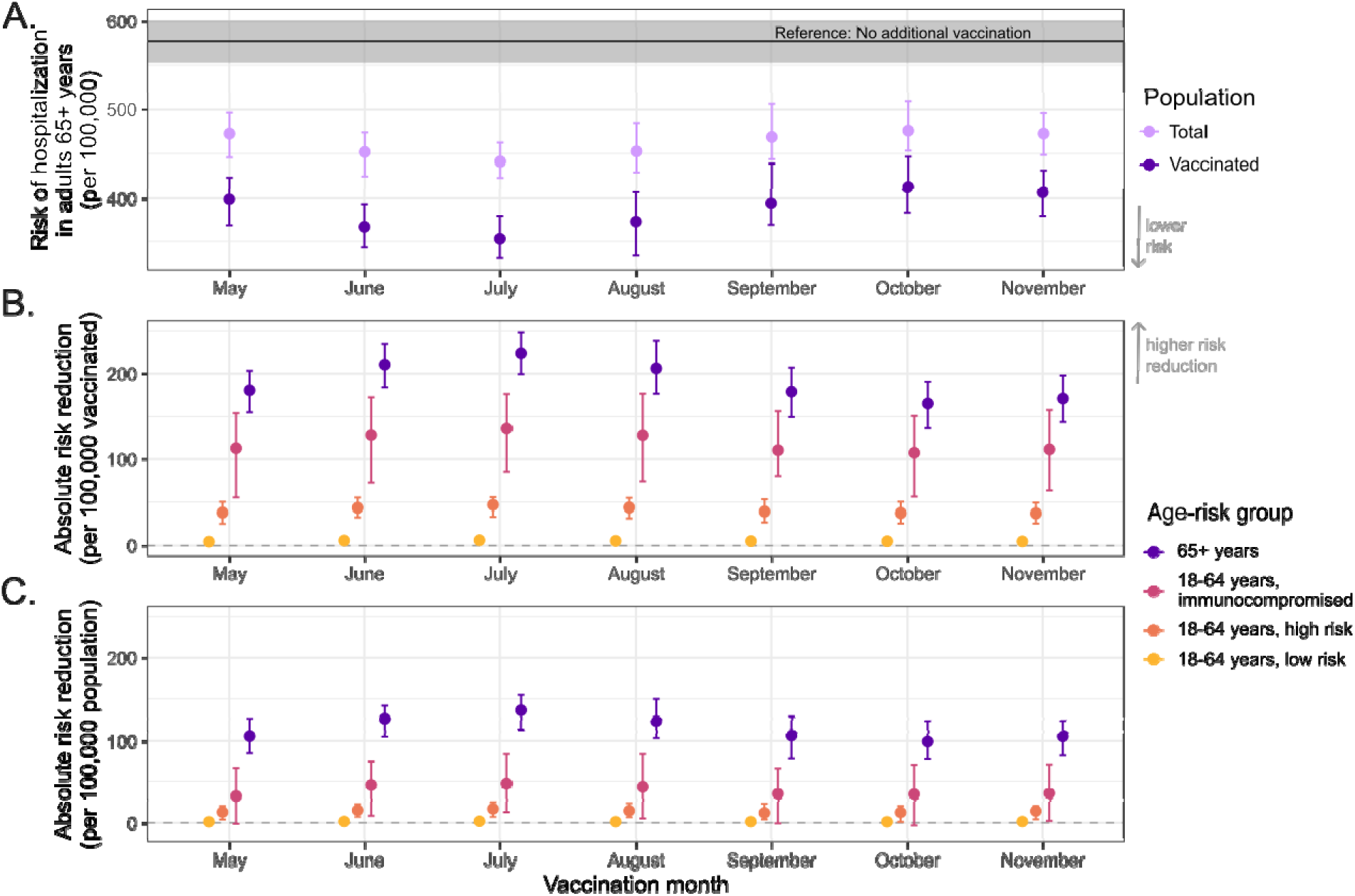
Impact of vaccination timing on COVID-19 hospitalization risk by age-risk group. We modeled universal annual vaccination (≥6 months) with historical vaccine coverage distributed over a single month (x-axis) and assessed COVID-19 hospitalization risk over the 12-month study period May 2024 to April 2025. (A) Annual hospitalization risk per 100,000 in adults ≥65 years: reference group with no additional vaccination during the study period (black line with 95% UI), total population (unvaccinated and vaccinated, light purple), and individuals vaccinated during the study period (dark purple). (B-C) Absolute risk reduction per 100,000 among vaccinated individuals (B) and the total population (C), in comparison to no additional vaccination, by age-risk group: ≥65 years (purple), 18–64 years immunocompromised (pink), 18–64 years high risk (orange), and 18–64 years low risk (yellow). Points show medians with 95% uncertainty intervals.

**Table 1:**
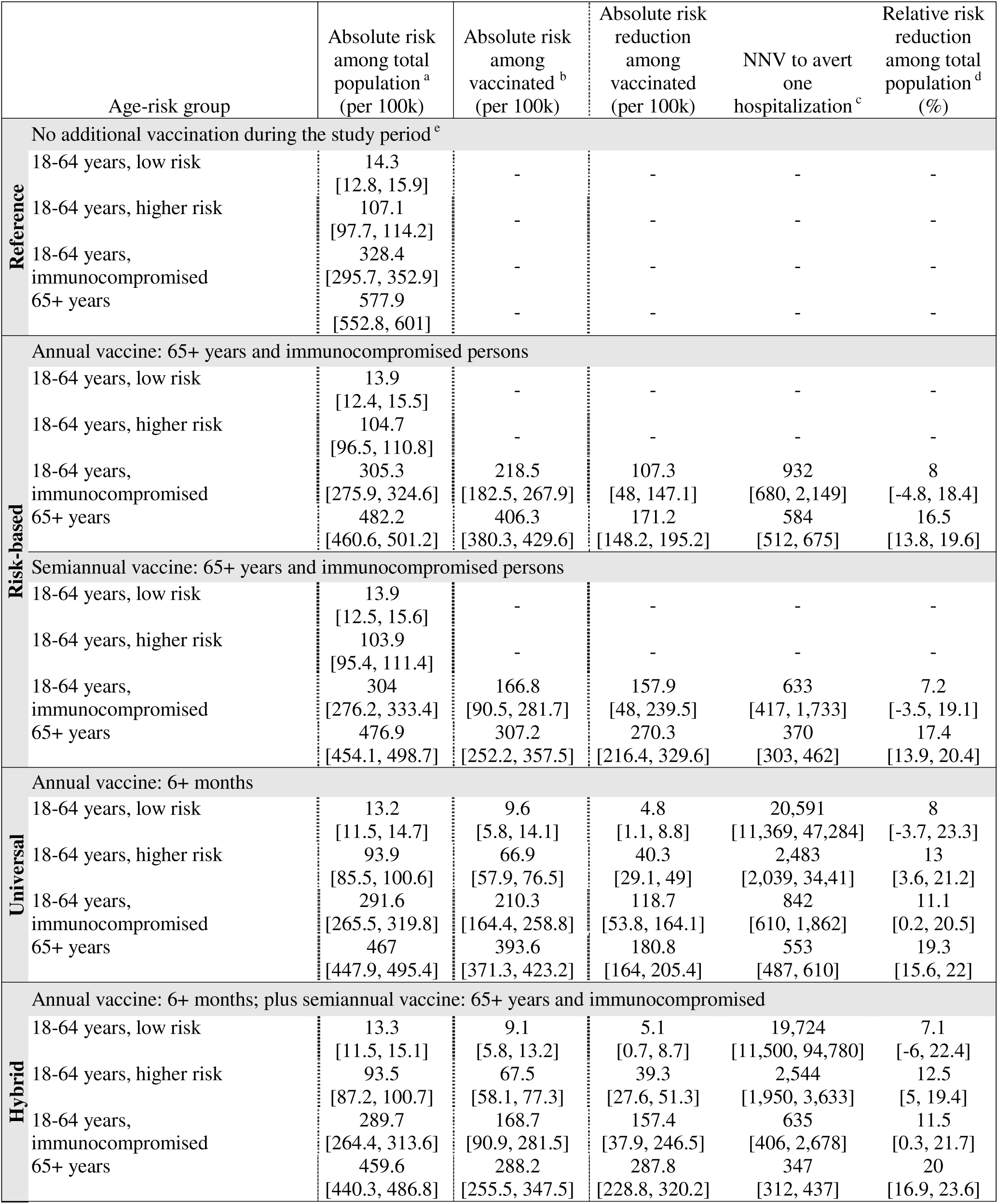
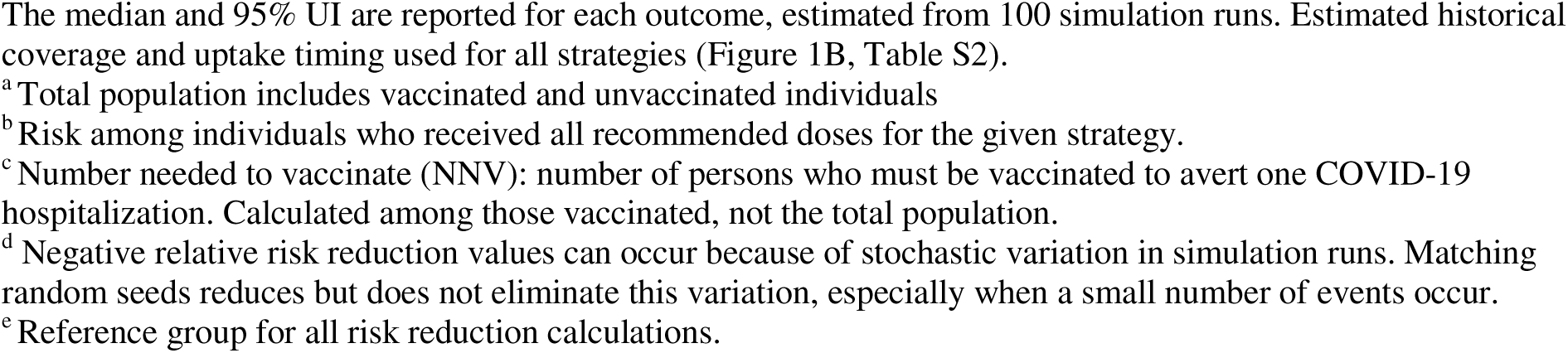
Model-based estimates of COVID-19 hospitalization risk under different vaccination strategies over 12-month study period with historical vaccine uptake.

|  | Age-risk group | Absolute risk among total population<sup>a</sup><br>(per 100k) | Absolute risk among vaccinated<sup>b</sup><br>(per 100k) | Absolute risk reduction among vaccinated<br>(per 100k) | NNV to avert one hospitalization<sup>c</sup> | Relative risk reduction among total population<sup>d</sup><br>(%) |
| --- | --- | --- | --- | --- | --- | --- |
| **Reference** | No additional vaccination during the study period<sup>e</sup> |  |  |  |  |  |
|  | 18-64 years, low risk | 14.3<br>[12.8, 15.9] | - | - | - | - |
|  | 18-64 years, higher risk | 107.1<br>[97.7, 114.2] | - | - | - | - |
|  | 18-64 years, immunocompromised | 328.4<br>[295.7, 352.9] | - | - | - | - |
|  | 65+ years | 577.9<br>[552.8, 601] | - | - | - | - |
| **Risk-based** | Annual vaccine: 65+ years and immunocompromised persons |  |  |  |  |  |
|  | 18-64 years, low risk | 13.9<br>[12.4, 15.5] | - | - | - | - |
|  | 18-64 years, higher risk | 104.7<br>[96.5, 110.8] | - | - | - | - |
|  | 18-64 years, immunocompromised | 305.3<br>[275.9, 324.6] | 218.5<br>[182.5, 267.9] | 107.3<br>[48, 147.1] | 932<br>[680, 2,149] | 8<br>[-4.8, 18.4] |
|  | 65+ years | 482.2<br>[460.6, 501.2] | 406.3<br>[380.3, 429.6] | 171.2<br>[148.2, 195.2] | 584<br>[512, 675] | 16.5<br>[13.8, 19.6] |
|  | Semiannual vaccine: 65+ years and immunocompromised persons |  |  |  |  |  |
|  | 18-64 years, low risk | 13.9<br>[12.5, 15.6] | - | - | - | - |
|  | 18-64 years, higher risk | 103.9<br>[95.4, 111.4] | - | - | - | - |
|  | 18-64 years, immunocompromised | 304<br>[276.2, 333.4] | 166.8<br>[90.5, 281.7] | 157.9<br>[48, 239.5] | 633<br>[417, 1,733] | 7.2<br>[-3.5, 19.1] |
|  | 65+ years | 476.9<br>[454.1, 498.7] | 307.2<br>[252.2, 357.5] | 270.3<br>[216.4, 329.6] | 370<br>[303, 462] | 17.4<br>[13.9, 20.4] |
| **Universal** | Annual vaccine: 6+ months |  |  |  |  |  |
|  | 18-64 years, low risk | 13.2<br>[11.5, 14.7] | 9.6<br>[5.8, 14.1] | 4.8<br>[1.1, 8.8] | 20,591<br>[11,369, 47,284] | 8<br>[-3.7, 23.3] |
|  | 18-64 years, higher risk | 93.9<br>[85.5, 100.6] | 66.9<br>[57.9, 76.5] | 40.3<br>[29.1, 49] | 2,483<br>[2,039, 34,41] | 13<br>[3.6, 21.2] |
|  | 18-64 years, immunocompromised | 291.6<br>[265.5, 319.8] | 210.3<br>[164.4, 258.8] | 118.7<br>[53.8, 164.1] | 842<br>[610, 1,862] | 11.1<br>[0.2, 20.5] |
|  | 65+ years | 467<br>[447.9, 495.4] | 393.6<br>[371.3, 423.2] | 180.8<br>[164, 205.4] | 553<br>[487, 610] | 19.3<br>[15.6, 22] |
| **Hybrid** | Annual vaccine: 6+ months; plus semiannual vaccine: 65+ years and immunocompromised |  |  |  |  |  |
|  | 18-64 years, low risk | 13.3<br>[11.5, 15.1] | 9.1<br>[5.8, 13.2] | 5.1<br>[0.7, 8.7] | 19,724<br>[11,500, 94,780] | 7.1<br>[-6, 22.4] |
|  | 18-64 years, higher risk | 93.5<br>[87.2, 100.7] | 67.5<br>[58.1, 77.3] | 39.3<br>[27.6, 51.3] | 2,544<br>[1,950, 3,633] | 12.5<br>[5, 19.4] |
|  | 18-64 years, immunocompromised | 289.7<br>[264.4, 313.6] | 168.7<br>[90.9, 281.5] | 157.4<br>[37.9, 246.5] | 635<br>[406, 2,678] | 11.5<br>[0.3, 21.7] |
|  | 65+ years | 459.6<br>[440.3, 486.8] | 288.2<br>[255.5, 347.5] | 287.8<br>[228.8, 320.2] | 347<br>[312, 437] | 20<br>[16.9, 23.6] |

Next, we evaluated whether summer vaccination remained optimal under different seasonal trends observed at the state level by generating two state archetypes that deviated from the US national pattern (Figure 3A). We found that the gains from vaccinating in summer (instead of historical fall vaccination) were greater in Archetype 1 than in Archetype 2. For Archetype 1, assuming one-month vaccine uptake, the largest risk reductions were achieved with June and July vaccination (228 [95% UI: 203-256] and 225 [187-263] averted COVID-19 hospitalizations per 100,000 persons among vaccinated adults ≥65 years, respectively), compared with September vaccination (146 [117-179]; Figure 3B-C, Tables S8-S9). For Archetype 2, July and August vaccination performed best (256 [223-284] and 256 [225-302] averted COVID-19 hospitalizations per 100,000 persons among vaccinated adults ≥65 years, respectively), compared with September vaccination (236 [199-279]; Figure 3B-C, Tables S10-S11).

**Figure 3:**
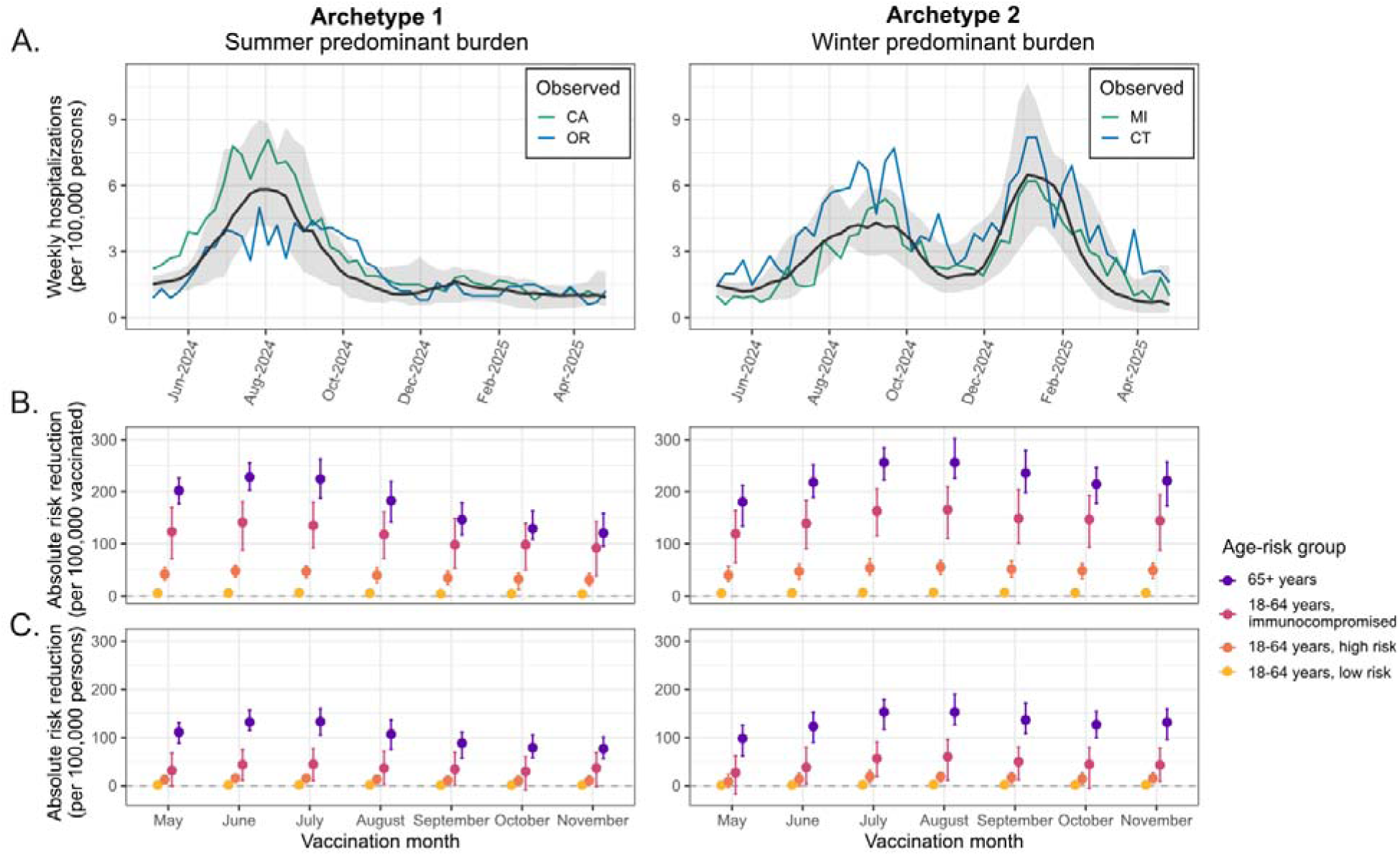
Impact of vaccination timing on COVID-19 hospitalization risk under state-level seasonal patterns. (A) Weekly COVID-19 hospitalizations per 100,000 from May 2024 to April 2025 for individual states (colored lines, see legend) and for the corresponding modeled seasonal archetypes under historical fall vaccination timing (black line, gray shading indicates the 95% uncertainty interval): Archetype 1 (summer-predominant burden; left) and Archetype 2 (winter-predominant burden; right). (B-C) Annual absolute risk reduction per 100,000 among vaccinated individuals (B) and the total population (C) by vaccination month (x-axis) and age-risk group in comparison to no additional vaccination during the study period, for Archetype 1 (left) and Archetype 2 (right): ≥65 years (purple), 18–64 years immunocompromised (pink), 18–64 years high risk (orange), and 18–64 years low risk (yellow). Points show medians with 95% uncertainty intervals. B and C assume annual universal vaccination (≥6 months) with historical vaccine coverage achieved over one month.

We also evaluated whether summer vaccination would remain optimal if summer coverage declined, a plausible consequence of departing from established respiratory virus public health campaigns in the fall. We reduced coverage by 5 or 10 absolute percentage points below historical levels across all age-risk groups, corresponding to approximately 25% and 50% relative reductions overall (Figure S6A). Among vaccinated adults ≥65 years, summer timing under both reductions remained more favorable than September timing with historical coverage (Figure S6B, Table S12). However, among *all* adults ≥65 years (including those not vaccinated during the study period), summer vaccination with 5-percentage-point lower coverage yielded COVID-19 hospitalization risk similar to that under September vaccination with historical coverage, and higher risk under a 10-percentage-point reduction (Figure S6C), largely due to reduced direct protection given reduced vaccine coverage in this scenario.

### Comparison of vaccination schedules based on eligibility and frequency

Because changing vaccination timing presents operational challenges of uncertain feasibility, we next evaluated what could be achieved through eligibility and dosing frequency alone, comparing strategies under historical vaccine coverage and timing (Figure 4A). Hybrid and risk-based semiannual vaccination, the two strategies adding a second annual dose for the highest risk adults, achieved the largest reductions in hospitalization risk among vaccinated adults ≥65 years.

**Figure 4:**
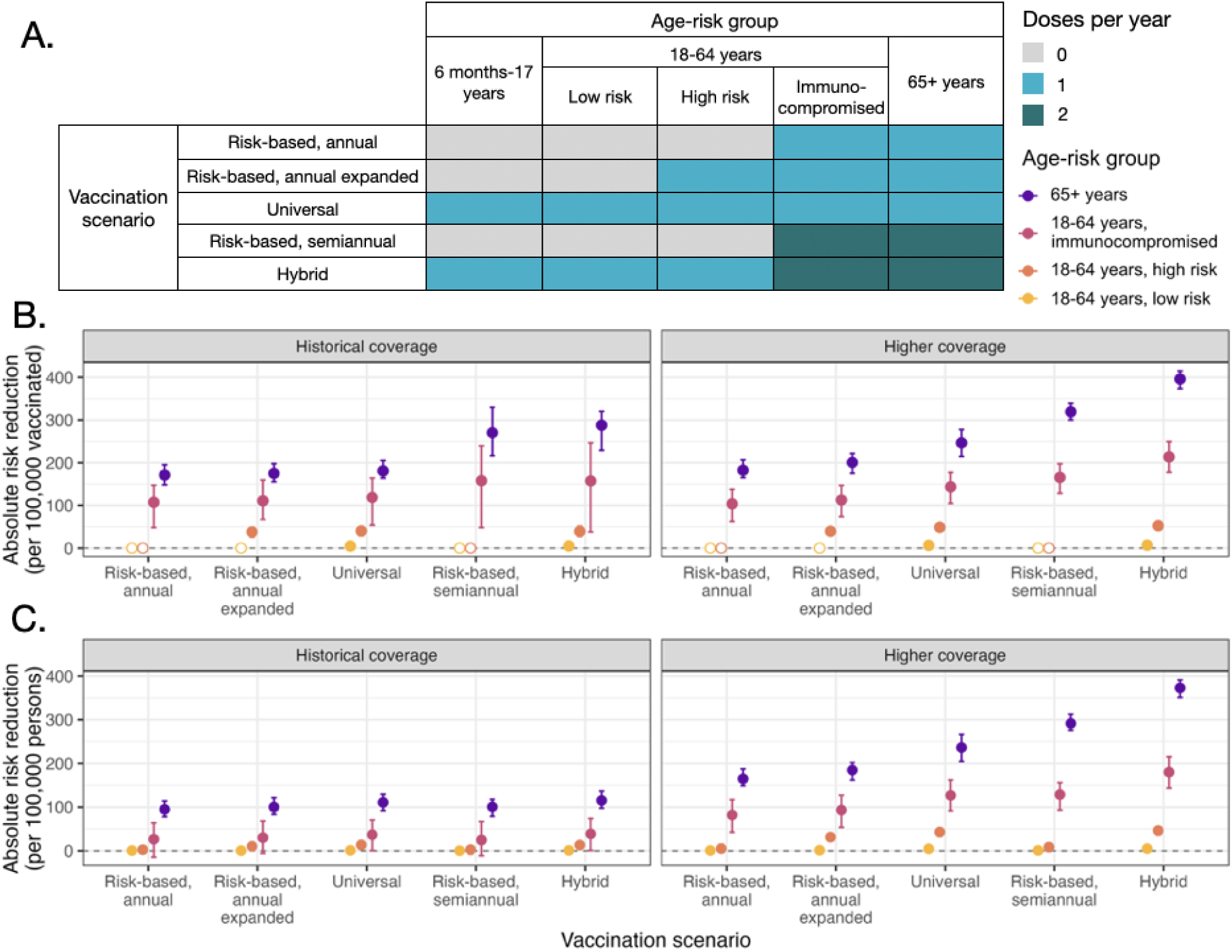
Comparison of COVID-19 vaccination strategies by eligibility and dosing frequency under historical fall timing. (A) Schematic of five vaccination strategies varying by eligibility criteria (universal, risk-based, and hybrid) and dosing frequency (annual vs. semiannual). (B, C) Annual absolute risk reduction per 100,000 among vaccinated individuals (B) and (C) in the total population compared to no additional vaccination, under historical coverage (left) and higher coverage (right) scenarios, by age-risk group: ≥65 years (purple), 18– 64 years immunocompromised (pink), 18–64 years high risk (orange), and 18–64 years low risk (yellow). Open circles in (B) indicate subgroups ineligible for vaccination under that vaccination scenario. Coverage levels defined in Table S2. Points show medians with 95% uncertainty intervals.

Under historical coverage, these strategies averted 288 [95% UI: 228-320] and 270 [216-330] COVID-19 hospitalizations per 100,000 persons among vaccinated adults ≥65 years, respectively, followed by universal annual vaccination (181 [164-205]; Figure 4B, Table 1). This ranking held in the total population for adults ≥65 and immunocompromised adults 18-64 years, although differences between strategies were smaller (Figure 4B-C). No strategy demonstrated clear, clinically meaningful superiority for low-or high-risk adults 18-64 years, due to low vaccine coverage and lower baseline risk of hospitalization. Under higher coverage, the ranking was unchanged, but population-level differences larger, particularly for higher-risk groups (Figure 4B-C, Table S14-S16).

We also considered the number of workdays missed due to COVID-19 infection and vaccination, an outcome particularly relevant for adults with low hospitalization risk. All vaccination strategies reduced total workdays missed compared to no additional vaccination, indicating that vaccination-related absenteeism was more than offset by prevented infections (Table S17). The hybrid strategy performed best, reducing the number of workdays missed by 33,076 [95% UI: 21,029-45,945] in a total population of 5 million. Under this strategy, 96% of the workdays still missed were due to infection and 4% to vaccine side effects.

### Comparison of changing vaccine timing versus vaccination schedules

Compared directly with hybrid and semiannual risk-based vaccination under historical fall timing, universal annual vaccination in July minimized hospitalization risk in the total population across every age-risk group, with the advantage concentrated among adults ≥65 years (averting 19.9 [95% UI: 0.7–40.1] and 36.1 [14.3–54.1] more hospitalizations per 100,000 persons, respectively). Among the vaccinated alone, however, hybrid vaccination remained most favorable for adults ≥65 years, reflecting the benefits of semiannual vaccination since vaccinated individuals only receive one dose under universal annual vaccination (Tables S18-S19).

## Discussion

This modeling study evaluated timing, eligibility, and dosing frequency of COVID-19 vaccination amid changing SARS-CoV-2 seasonality to summer predominance in the US. The most effective vaccine policy depends on whether annual COVID-19 vaccination timing can change. If earlier annual COVID-19 vaccine timing is feasible and coverage is maintained, universal vaccination in the summer (July) prevented more hospitalizations than the status quo hybrid strategy with historical fall timing. If restricted to historical timing, the hybrid vaccination strategy, equivalent to recommendations in place during the study period, minimized hospitalizations among the vaccinated population in the highest-risk groups at the national level. Semiannual risk-based vaccination is a reasonable alternative, requiring far fewer doses and achieving similar protection in those with the highest risk, though it provides less benefit to the wider population than universal vaccination. The magnitude of benefit from summer-timed and semiannual vaccination in the highest-risk groups varied regionally.

At the US national level, the benefits of summer COVID-19 vaccination timing likely generalize beyond our 2024-25 study period in the near term, as 2025-26 COVID-19 hospitalization dynamics closely matched observed patterns in shape and peak timing (Figure S7). Additionally, because the interval from summer to winter peaks (5 months) has been shorter than winter to summer (7 months), summer vaccination is more likely to provide protection across both peaks than fall timing (Figure S3). Under historical coverage, shifting universal annual vaccination to the summer averted 19.9 [95% UI: 0.7-40.1] more COVID-19 hospitalizations per 100,000 adults ≥65 years than the hybrid strategy with historical fall timing (Tables S18-S19), corresponding to several thousand hospitalizations annually among US adults ≥65 years.

However, implementing earlier COVID-19 vaccination timing would present several practical challenges that could undermine its benefits. Shifting from fall timing may reduce vaccine coverage due to operational, regulatory, and messaging barriers, as summer vaccination departs from established respiratory virus public health campaigns and co-administration with influenza vaccines^38,39^. Our sensitivity analyses suggest that a 10-percentage point drop in coverage (corresponding to a 50% relative decrease) would nullify the timing advantage at the national-level and could even worsen outcomes (Figures S8-S9). Earlier vaccination would require either moving strain selection earlier or compressing production timelines. Vaccine effectiveness may also decline over the year due to viral evolution away from the vaccine strain, although this has been difficult for prior studies to distinguish from waning protection^40,41^. Our modeling approach partially accounts for this decline by fitting waning curves to observed effectiveness over time.

If fall vaccination timing is maintained, differences between strategies were small when comparing risk in the total population (i.e., including all persons with and without additional vaccination) under historical coverage. This reflects extremely low uptake of the second dose (5– 9% of eligible individuals) and low overall coverage in lower-risk adults, which limited the population-level indirect effects. Because the hybrid strategy corresponds to US recommendations during the study period, these comparisons indicate limited possibility for further reducing hospitalizations through eligibility or dosing frequency alone. Larger gains would require higher coverage or earlier vaccination timing.

COVID-19 vaccination in July-August performed well under the US national model and both state-level archetypes, suggesting earlier summer-timed vaccination as a pragmatic basis for uniform national guidance. In states with two large peaks over both summer and winter (similar to Archetype 2), the use of a second dose in the highest-risk populations may have additional public health benefit, whereas a second dose in locations similar to Archetype 1 (such as California and Oregon) without a winter peak are unlikely to have substantial benefit. Recent studies attribute these regional differences in disease dynamics to rapidly waning immunity and climate forcing^10,32^, which are expected to persist. However, unlike national dynamics, the relative size of summer and winter waves has not yet stabilized in some states (Figures S10-S11). Region- or state-specific vaccine policy would also be relatively novel and likely operationally challenging to implement, although could potentially further improve the public health impact of vaccination.

Our study has several limitations. Our vaccine effectiveness estimates, based on observational studies from 2022-2024^26–28^, may be subject to bias and confounding, affecting absolute benefit estimates more than relative strategy rankings. Our 12-month evaluation window may slightly favor earlier vaccination timing since earlier doses have longer to demonstrate benefits (Supplementary Section S6). We did not model a state-level archetype with a single winter peak as this was not observed among COVID-NET states, though such a pattern would favor later timing. We modeled immunocompromised and high-risk adults as discrete groups, though substantial heterogeneity exists within these categories^42^. We varied timing of the annual dose but not under semiannual schedules; a summer COVID-19 vaccine dose followed by a second dose later could better protect locations with both a summer and winter peak, although population-level gains would be constrained by low historical second-dose uptake (5–9% of eligible individuals). Finally, we did not model pediatric hospitalizations due to low hospitalization rates, though children were included in transmission dynamics, making findings most relevant for adult vaccination policy.

In conclusion, these findings can inform evidence-based COVID-19 vaccination policy as SARS-CoV-2 transitions to endemic circulation.

## Funding and Acknowledgements

This project is supported by a National Institutes of Health New Innovator Award (DP2AI170485 to NCL). KMB acknowledges support from a National Institutes of Health training grant (T32AI007502) and the Stanford Medicine Sue and Thomas Merigan Postdoctoral Fellowship. The funding source had no role in the design, conduct, or analysis of this study or in the decision to submit the manuscript for publication. The findings and conclusions in this article are those of the authors and do not necessarily represent the views or opinions of the California Department of Public Health, California Health and Human Services Agency, or National Institutes of Health.

## Data and Code Availability

This study used publicly available datasets. Details on these datasets and other epidemiological inputs can be found in the Methods and Supplementary Materials. Model input data and analytic code are publicly available on an online repository: https://github.com/kbubar/comparison-vaccine-schedules. Analysis was conducted in R (version 4.4.2).

## Ethical Approval

This study was not human subjects research given use of computer simulation and publicly available, non-identifiable secondary datasets.

## Authorship contribution

Dr. Kate Bubar, Ms. Hailey Park, and Dr. Nathan Lo had full access to all the data in the study and take responsibility for the integrity of the data and the accuracy of the data analysis.

Study concept and design: RS, HJP, KMB, NCL

Statistical analysis: KMB, HJP, NCL Analytic coding: KMB, HJP

Acquisition, analysis, or interpretation of data: All authors

First draft of the manuscript: KMB, HJP, NCL

Critical revision of the manuscript: All authors

Contributed intellectual material and approved final draft: All authors

## Competing interests

The authors have no conflicts to declare.

## AI declaration

AI tools were only used for routine coding tasks and some minor editorial assistance including grammar, clarity, and formatting. Claude (Sonnet 4 and Opus 5), Cursor (version 3.14.7), and ChatGPT (GPT-5.5) were applied for these applications. All substantive ideas, analysis, methodology, interpretation, and conclusions are the original work of the authors, who retain full responsibility for the content and accuracy of this work.

## Data Availability

Model input data and analytic code are publicly available on an online repository: https://github.com/kbubar/comparison-vaccine-schedules.

## Notes

### Competing Interest Statement

The authors have declared no competing interest.

### Author Declarations

This study used publicly available datasets. Details on these datasets and other epidemiological inputs can be found in the Methods and Supplementary Materials.

## References

1. Silk BJ, Prill MM, Winn AK, et al. Respiratory Virus Activity — United States, July 1, 2024– June 30, 2025. Morb Mortal Wkly Rep. 2026;75(6):77–84. doi:10.15585/mmwr.mm7506a2

2. Taylor CA, Patel K, Pham H, et al. COVID-19-Associated Hospitalizations Among U.S. Adults Aged ≥18 Years - COVID-NET, 12 States, October 2023-April 2024. Morb Mortal Wkly Rep. 2024;73(39):869–875. doi:10.15585/mmwr.mm7339a2

3. Centers for Disease Control and Prevention. Coronavirus Disease 2019 (COVID-19) Hospitalization Surveillance Network (COVID-NET). Published online January 30, 2026. Accessed June 24, 2026. https://www.cdc.gov/covid/php/covid-net/index.html

4. Centers for Disease Control and Prevention. Underlying Conditions and the Higher Risk for Severe COVID-19. Covid. May 12, 2026. Accessed June 24, 2026. https://www.cdc.gov/covid/hcp/clinical-care/underlying-conditions.html

5. Panagiotakopoulos L, Moulia DL, Godfrey M, et al. Use of COVID-19 Vaccines for Persons Aged ≥6 Months: Recommendations of the Advisory Committee on Immunization Practices — United States, 2024–2025. Morb Mortal Wkly Rep. 2024;73(37):819–824. doi:10.15585/mmwr.mm7337e2

6. Roper LE, Godfrey M, Link-Gelles R, et al. Use of Additional Doses of 2024–2025 COVID-19 Vaccine for Adults Aged ≥65 Years and Persons Aged ≥6 Months with Moderate or Severe Immunocompromise: Recommendations of the Advisory Committee on Immunization Practices — United States, 2024. Morb Mortal Wkly Rep. 2024;73(49):1118–1123. doi:10.15585/mmwr.mm7349a2

7. Assistant Secretary for Public Affairs (ASPA). ACIP Recommends COVID-19 Immunization Based on Individual Decision-making. September 19, 2025. Accessed September 8, 2026. https://www.hhs.gov/press-room/acip-recommends-covid19-vaccination-individual-decision-making.html

8. O’Reilly KB. Doctors united on respiratory virus vaccine recommendations. American Medical Association. September 2, 2026. Accessed September 8, 2026. https://www.ama-assn.org/public-health/prevention-wellness/doctors-united-respiratory-virus-vaccine-recommendations

9. Lipson RA, Senerth E, Watson MA, et al. COVID-19 Vaccine Effectiveness and Safety for the 2026-2027 Respiratory Season. JAMA. Published online September 2, 2026. doi:10.1001/jama.2026.18191

10. Bents SJ, Bubar KM, Park HJ, et al. Interplay of Immunity, Climate, and Viral Evolution Explains Semiannual SARS-CoV-2 Dynamics with Implications for Control. *medRxiv*. Preprint posted online March 2, 2026:2026.02.27.26347213. doi:10.64898/2026.02.27.26347213

11. Rubin IN, Bushman M, Lipsitch M, Hanage WP. Seasonal forcing and waning immunity drive the sub-annual periodicity of the COVID-19 epidemic. PLOS Pathog. 2026;22(4):e1014169. doi:10.1371/journal.ppat.1014169

12. Wiegand RE, Payne AB, Mak J, et al. Estimated Effectiveness of 2024-2025 COVID-19 Vaccines in Adults. JAMA Intern Med. Published online June 15, 2026. doi:10.1001/jamainternmed.2026.1936

13. Ma KC, Webber A, Lauring AS, et al. Estimated Effectiveness of 2024-2025 COVID-19 Vaccination Against Severe COVID-19. JAMA Netw Open. 2026;9(2):e2557415. doi:10.1001/jamanetworkopen.2025.57415

14. Kirwan PD, Foulkes S, Munro K, et al. Protection of vaccine boosters and prior infection against mild/asymptomatic and moderate COVID-19 infection in the UK SIREN healthcare worker cohort: October 2023 to March 2024. J Infect. 2024;89(5):106293. doi:10.1016/j.jinf.2024.106293

15. Wells CR, Pandey A, Moghadas SM, Fitzpatrick MC, Singer BH, Galvani AP. Evaluation of strategies for transitioning to annual SARS-CoV-2 vaccination campaigns in the United States. Ann Intern Med. 2024;177(5):609–617. doi:10.7326/M23-2451

16. Park HJ, Gonsalves GS, Tan ST, et al. Comparing frequency of booster vaccination to prevent severe COVID-19 by risk group in the United States. Nat Commun. 2024;15(1):1883. doi:10.1038/s41467-024-45549-9

17. Kelly SL, Le Rutte EA, Richter M, Penny MA, Shattock AJ. COVID-19 Vaccine Booster Strategies in Light of Emerging Viral Variants: Frequency, Timing, and Target Groups. Infect Dis Ther. 2022;11(5):2045–2061. doi:10.1007/s40121-022-00683-z

18. Townsend JP, Hassler HB, Dornburg A. Optimal Annual COVID-19 Vaccine Boosting Dates Following Previous Booster Vaccination or Breakthrough Infection. Clin Infect Dis. 2024;80(2):316–322. doi:10.1093/cid/ciae559

19. Bartsch SM, Weatherwax C, Wasserman MR, et al. How the Timing of Annual COVID-19 Vaccination of Nursing Home Residents and Staff Affects Its Value. J Am Med Dir Assoc. 2024;25(4):639–646.e5. doi:10.1016/j.jamda.2024.02.005

20. Centers for Disease Control and Prevention. Patient Characteristics of Laboratory-Confirmed COVID-19 Hospitalizations from the COVID-NET Surveillance System. Accessed June 24, 2026. https://data.cdc.gov/Public-Health-Surveillance/Patient-Characteristics-of-Laboratory-Confirmed-CO/bigw-pgk2/about_data

21. Taylor CA. COVID-19–Associated Hospitalizations Update — COVID-NET, July 2023– September 2024. Slide presentation presented at: Meeting of the Advisory Committee on Immunization Practices; October 23, 2024; Atlanta, GA. Accessed September 14, 2026. https://www.cdc.gov/acip/downloads/slides-2024-10-23-24/03-COVID-Taylor-508.pdf

22. Centers for Disease Control and Prevention. COVID-19 Vaccination Age and Sex Trends in the United States, National and Jurisdictional. Published online May 11, 2023. Accessed June 24, 2026. https://data.cdc.gov/Vaccinations/COVID-19-Vaccination-Age-and-Sex-Trends-in-the-Uni/5i5k-6cmh/about_data

23. Centers for Disease Control and Prevention. COVID-19 Vaccination Coverage, Overall and by Selected Demographics and Jurisdiction, Among Adults 18 Years and Older, by Season. Published online March 4, 2026. Accessed July 3, 2026. https://data.cdc.gov/Vaccinations/COVID-19-Vaccination-Coverage-Overall-and-by-Selec/ksfb-ug5d/about_data

24. Centers for Disease Control and Prevention. Weekly Parental Intent for Vaccination and Cumulative Percentage of Children 6 Months-17 Years Who are Up to date with the COVID-19 Vaccines by Season, United States. Published online July 2, 2026. Accessed July 3, 2026. https://data.cdc.gov/Child-Vaccinations/Weekly-Parental-Intent-for-Vaccination-and-Cumulat/ker6-gs6z/data_preview

25. Centers for Disease Control and Prevention. National Immunization Survey Adult COVID Module (NIS-ACM). Accessed July 8, 2026. https://data.cdc.gov/Vaccinations/National-Immunization-Survey-Adult-COVID-Module-NI/si7g-c2bs/data_preview

26. Bobrovitz N, Ware H, Ma X, et al. Protective effectiveness of previous SARS-CoV-2 infection and hybrid immunity against the omicron variant and severe disease: a systematic review and meta-regression. Lancet Infect Dis. 2023;23(5):556–567. doi:10.1016/S1473-3099(22)00801-5

27. Carazo S, Skowronski DM, Brisson M, et al. Effectiveness of previous infection-induced and vaccine-induced protection against hospitalisation due to omicron BA subvariants in older adults: a test-negative, case-control study in Quebec, Canada. Lancet Healthy Longev. 2023;4(8):e409–e420. doi:10.1016/S2666-7568(23)00099-5

28. Lau JJ, Cheng SMS, Leung K, et al. Real-world COVID-19 vaccine effectiveness against the Omicron BA.2 variant in a SARS-CoV-2 infection-naive population. Nat Med. 2023;29(2):348–357. doi:10.1038/s41591-023-02219-5

29. Wu Y, Guo Z, Yuan J, et al. Duration of viable virus shedding and polymerase chain reaction positivity of the SARS-CoV-2 Omicron variant in the upper respiratory tract: a systematic review and meta-analysis. Int J Infect Dis. 2023;129:228–235. doi:10.1016/j.ijid.2023.02.011

30. Li Y, Jiang X, Qiu Y, et al. Latent and incubation periods of Delta, BA.1, and BA.2 variant cases and associated factors: a cross-sectional study in China. BMC Infect Dis. 2024;24(1):294. doi:10.1186/s12879-024-09158-7

31. National Center for Immunization and Respiratory Diseases (U.S.) Division of Viral Diseases. Reinfections and COVID 19. September 9, 2022. Accessed August 24, 2026. https://stacks.cdc.gov/view/cdc/121105

32. Stamper A, Baker RE. Spatial patterns and environmental influences of COVID-19 outbreaks, post-Omicron. PLOS ONE. 2026;21(2):e0342510. doi:10.1371/journal.pone.0342510

33. Centers for Disease Control and Prevention. NHSN Hospital Respiratory Data (HRD) Dashboard. Published online September 26, 2025. Accessed August 24, 2026. https://www.cdc.gov/nhsn/psc/hospital-respiratory-dashboard.html

34. Tan ST, Kwan AT, Rodríguez-Barraquer I, et al. Infectiousness of SARS-CoV-2 breakthrough infections and reinfections during the Omicron wave. Nat Med. 2023;29(2):358–365. doi:10.1038/s41591-022-02138-x

35. Tan ST, Rodríguez-Barraquer I, Kwan AT, et al. Strength and durability of indirect protection against SARS-CoV-2 infection through vaccine and infection-acquired immunity. Nat Commun. 2025;16(1):1090. doi:10.1038/s41467-024-55029-9

36. Prunas O, Warren JL, Crawford FW, et al. Vaccination with BNT162b2 reduces transmission of SARS-CoV-2 to household contacts in Israel. Science. 2022;375(6585):1151–1154. doi:10.1126/science.abl4292

37. Lyngse FP, Mortensen LH, Denwood MJ, et al. Household transmission of the SARS-CoV-2 Omicron variant in Denmark. Nat Commun. 2022;13(1):5573. doi:10.1038/s41467-022-33328-3

38. Goswami J, Cardona JF, Hsu DC, et al. Safety and immunogenicity of mRNA-1345 RSV vaccine coadministered with an influenza or COVID-19 vaccine in adults aged 50 years or older: an observer-blinded, placebo-controlled, randomised, phase 3 trial. Lancet Infect Dis. 2025;25(4):411–423. doi:10.1016/S1473-3099(24)00589-9

39. Sanborn J, Robertson MM, Penrose K, et al. Gaps between willingness and uptake of influenza and COVID-19 vaccines during the 2025–26 respiratory virus season in a U.S. adult cohort. Vaccine. 2026;92:129126. doi:10.1016/j.vaccine.2026.129126

40. Payne AB. Updates on COVID-19 Vaccine Effectiveness. Slide presentation presented at: Meeting of the Vaccines and Related Biological Products Advisory Committee (VRBPAC), US Food and Drug Administration; May 28, 2026; Silver Spring, MD. Accessed August 28, 2026. https://www.fda.gov/media/192771/download

41. Alhumaid S, Alkhars O, Algrafi AS, et al. Vaccine effectiveness across the Omicron evolutionary spectrum (BA.2, BA.5, XBB, JN.1, KP.3): a systematic review of studies published 2022–2025. BMC Infect Dis. 2026;26(1):1014. doi:10.1186/s12879-026-13281-y

42. Antinori A, Bausch-Jurken M. The Burden of COVID-19 in the Immunocompromised Patient: Implications for Vaccination and Needs for the Future. J Infect Dis. 2023;228(Suppl 1):S4–S12. doi:10.1093/infdis/jiad181

