## Supplement for "Rethinking seasonal COVID-19 vaccination in the endemic era: summer timing, eligibility, and frequency in the United States"

**Table of Contents**

**Supplementary Text:**

- **Section S1: Data processing**
- Definition of risk groups
- **Section S2: Model of waning immunity**
- **Section S3: SARS-CoV-2 transmission model**
- Model equations and implementation
- Model initialization
- Model calibration
- Model validation
- **Section S4: Study outcomes definitions and calculations**
- **Section S5: Archetypes representing state-level heterogeneity**
- **Section S6: Sensitivity analyses**

**Supplementary Tables**

**Supplementary Figures**

#### Section S1: Data processing

We obtained national age-stratified weekly COVID-19 hospitalization rates per 100,000 from the COVID-NET surveillance system (version downloaded on February 18, 2026 used for calibration and model analysis, version downloaded on July 12, 2026 used for most recent plots (e.g., Figure S3, S7, S10-11))^1^. We adjusted incidence estimates to only include the subset of hospitalizations where COVID-19 was the primary indication for hospital admission^2^. We considered 6 age groups: 0-17, 18-29, 30-49, 50-64, 65-74, and ≥75 years.

We estimated nonsevere (non-hospitalized) incidence from observed hospitalization incidence using published age-specific infection-severity risk (ISR) relationships with an additional calibration step to align to contemporary risk. We defined ISR as the proportion of infections resulting in severe disease, which we defined as hospitalization^3^. Reported values are [0.155, 0.4283, 1.545, 5.9, 13, 30] percent of infections for ages {0–17, 18–29, 30–49, 50–64, 65–74, 75+ years}. We assumed the relative gradient in severity across age groups follows the reported ISRs, normalized to the 50–64 year group $(rₐ = ISRₐ/ISR₅₀₋₆₄$), and calibrated a single severity scalar$\phi$ to set its absolute level, giving$\thetaₐ = rₐ \cdot\phi$. This allowed for an attenuation of overall clinical severity since the ISR was from data collected largely before the Omicron era and before widespread vaccination while maintaining age-specific risk relationships. Nonsevere incidence was then estimated as observed severe incidence divided by $\thetaₐ$. We assumed age-specific nonsevere incidence was consistent across risk groups (i.e., all risk groups of the same age had the same probability of infection). These estimates were used to set model initial conditions and to define the age- and risk-group-specific multipliers converting nonsevere to severe infection risk (Section S3). Because ϕ is a single time-invariant scalar applied across all age groups, it cannot absorb age- or time-varying components of the change in disease severity. Age-specific fits are described in Section S3.

##### Definition of risk groups

We consider three risk groups: immunocompetent (referred to as low risk throughout the main text), high risk, and immunocompromised. We define the high-risk group as persons with 1+ comorbid conditions that are associated with a higher risk of severe COVID-19. Comorbid conditions include one or more of the following conditions based on published lists: chronic lung disease, chronic metabolic disease, blood disorder/hemoglobinopathy, cardiovascular disease, neurologic disorder, renal disease, gastrointestinal/liver disease, rheumatologic/autoimmune/inflammatory condition, and obesity^4–7^. The immunocompromised group included those with moderate to severe immunocompromising conditions (e.g., hematologic malignancy with active treatment or poor response to vaccines, solid organ or bone marrow transplant, high-dose corticosteroids or other moderate/severely immunosuppressive medications)^8,9^.

#### Section S2: Model of waning immunity

Our model of waning immunity describes how protection against SARS-CoV-2 infection and COVID-19 related hospitalization wanes over time since the last vaccine dose or prior infection, following our prior work^10^.

We defined individual protection as absolute protection relative to an immunologically naïve baseline in the model itself for its technical implementation, although we use diverse data inputs including absolute and relative vaccine effectiveness data estimates to inform the model. We parameterized our model of waning immunity using predominately post-Omicron vaccine effectiveness estimates against circulating variants antigenically similar to recent vaccine formulations (Table S3). Absolute vaccine effectiveness data were derived from earlier periods of the pandemic and were higher than recent estimates given comparison to an immunologically naïve population. Relative vaccine effectiveness data came from recent studies that use comparators who lack the most recent dose but have substantial immunity from prior vaccination and infection.

We modeled twelve distinct protection curves that wane over time, varying by outcome (infection versus COVID-19 related hospitalization), immunity types (vaccine-acquired, infection-acquired, and hybrid being immunity from both previous vaccination and infection), and risk group (low risk/high risk versus immunocompromised).

To generate the protection curves, we fitted linear mixed-effects models with the log of (1 - protective effectiveness) as the outcome variable. Predictor variables included prior infection status and the log of months since the most recent immune event (either vaccination or COVID-19 infection). We modeled three estimates simultaneously (mean, lower bound, upper bound) by treating them as an ordinal variable, calibrating against literature values using the mean and 95% confidence interval of protective effectiveness (Tables S3, Figure S1). The model included random effects for each study and weighted studies by the inverse of their 95% confidence interval width to account for varying sample sizes and precision. We assumed protection was on average 15 percentage points lower than the immunocompetent population based on literature estimates^11,12^, and adjusted protection curves accordingly (Figure S1).

To validate this approach, we assessed whether fitted waning curves imply plausible relative protection from additional vaccination observed in the recent literature. We derived relative protective effectiveness from model-predicted absolute protective effectiveness ($PE$) using the standard relative-risk formulation ($1 - \frac{RR_{t}}{RR_{t+7}}$, where $RR = 1 - PE$). Protection at time $t$ represents individuals who received an additional dose, while protection at $t+7$months represents those that did not, assuming an average of 7 months since their last immune event and informed by literature estimating significant protection when vaccinated within a 6-month period. The model-derived relative $PE$ (vaccine-derived immunity for those without prior infection; hybrid immunity for those with prior infection) aligned well with published relative $VE$ estimates from observational studies of updated 2023-2024 and 2024-2025 boosters (Figure S2).

#### Section S3: SARS-CoV-2 transmission model

##### Model equations and implementation

Each day, each individual *k* (age group *i*, risk group *r*) had independent probabilities of nonsevere or severe COVID-19 infection, modeled as Bernoulli trials:

$$p_{nonsevere, k}\left( t \right)=1-\exp\left( -\lambda_{i}\left( t \right)\left( 1-{PE}_{nonsevere, r, v}\left( \tau\right) \right) \right),$$

$$p_{severe, k}\left( t \right)=1-\exp\left( -\lambda_{i}(t)\left( 1-{PE}_{severe, r, v}\left( \tau\right) \right)\theta_{i,r} \right)$$

where $\lambda_{i}(t)$ is the age-specific force of infection, ${PE}_{nonsevere,r,v}$ and ${PE}_{severe,r,v}$ are the protective effectiveness against SARS-CoV-2 infection and COVID-19 hospitalization for risk group *r* with immunity type $v$ (vaccine-acquired, infection-acquired, or hybrid) at time $\tau$ since last immune event, and $\theta_{i, r}$ is the age- and risk-specific severity ratio (i.e., ratio of severe to nonsevere incidence scaled by $\phi$, as described in Section S1). In rare instances where both events occurred on the same day, we classified the infection as severe.

We define the force of infection as:

$$\lambda_{i}(t)=\kappa_{t}\beta_{i}\sum_{j=1}^{6} C_{ij}\frac{I_{j}(t)}{N_{j}},$$

where $\kappa_{t}$ is a time-varying transmission coefficient, $\beta_{i}$ is an age-specific scaling constant, $C$ is an age-stratified contact matrix (Table A1), $I_{j}(t)$ is the number of infectious individuals in age group $j$, and $N_{j}$ is the total number of people in age group $j$. Notably, $\kappaₜ\betaᵢ$ is multiplicative in the force of infection, so $\betaᵢ$ determines the relative age pattern of transmission while $\kappaₜ$ determines its overall level and temporal variation.

Upon infection, individuals entered a 3-day latent period, followed by a 5-day infectious period^13,14^. After the infectious period, individuals recovered and their level of protection against future infection was reset. Individuals could not be reinfected within 90 days of a previous infection, in line with a commonly applied CDC definition of reinfection^15^. We did not model long COVID due to limited data on impact of repeated vaccination on risk and progression.

##### Table A1. Age-based contact matrix

| **Contacts** ^a^ | 0-17 years | 8.22 | 3.41 | 2.85 | 0.83 | 0.31 | 0.14 |
| --- | --- | --- | --- | --- | --- | --- | --- |
|  | 18-29 years | 2.02 | 6.60 | 2.71 | 1.05 | 0.12 | 0.07 |
|  | 30-49 years | 3.07 | 4.22 | 6.83 | 2.17 | 0.36 | 0.17 |
|  | 50-64 years | 1.91 | 2.75 | 3.11 | 2.93 | 0.56 | 0.21 |
|  | 65-74 years | 0.60 | 0.53 | 0.87 | 0.79 | 1.17 | 0.31 |
|  | 75+ years | 0.31 | 0.19 | 0.32 | 0.26 | 0.24 | 0.40 |
|  |  | 0-17 years | 18-29 years | 30-49 years | 50-64 years | 65-74 years | 75+ years |
|  |  | **Individual** | | | | | |

^a^ These are average contacts per day.

Contact matrix based on published study^16^.

The model was implemented using a daily time step, with an initial 10-month pre-study period followed by a one-year study period. Individual $PE$ values were updated immediately upon vaccination or infection and otherwise recalculated every two weeks for computational efficiency. Four vaccine doses were distributed throughout the simulation: 2023-2024 formulation in fall 2023 (Dose 0 during the pre-study period) and spring 2024 (Dose 1), and 2024-2025 formulation in fall 2024 (Dose 2) and spring 2025 (Dose 3) following observed uptake and coverage data.

##### Model initialization

We initialized our simulated population's immune history to broadly match the immune history estimated in the United States around the start of the pre-study period, July 2023. Individuals were assigned both a vaccination status (unvaccinated, primary series, one booster, or 2+ boosters), and prior infection status (0, 1, or 2+), and a time since last vaccination and/or infection.

We assigned these immune histories using age-specific joint distributions (Table A4), derived by solving equations that synthesize population-level immunity estimates (Table A2) with age-specific data (Table A3). We assumed all individuals had either prior vaccination, infection, or both. In rare cases of discrepancies, we deferred to CDC data. We used Klaassen's infection estimates (due to known seroreversion in CDC nucleocapsid assays^17^) but CDC vaccination data (closer to reported administration records).

**Table A2: SARS-CoV-2 immune history in the United States in July 2023.**

| **Immunity status** | **Prevalence (%)** |
| --- | --- |
| Immune naive | 2.8 |
| Infected | 3.7 |
| Reinfected | 14.3 |
| Infected + Vaccinated | 12 |
| Reinfected + Vaccinated | 29.8 |
| Infected + Boosted | 14.3 |
| Reinfected + Boosted | 15.9 |
| Boosted | 4.4 |
| Vaccinated | 2.8 |
| Prior infection | 90 |
| Booster | 34.6 |
| Fully vaccinated only | 44.6 |

Reference: estimates reported on covidestim.org, which are a continuation of Klaassen et al.^18^

**Table A3: Age-specific prevalence (%) of SARS-CoV-2 prior infection and COVID-19 vaccination status by age group in the United States, July 2023**

| **Immunity status** | **0-17**  **years** | **18-29**  **years** | **30-49**  **years** | **50-64**  **years** | **65+**  **years** | **Reference** |
| --- | --- | --- | --- | --- | --- | --- |
| Prior infection | 86.3 | 83 | 77.9 | 70.3 | 46.6 | US CDC national serologic surveys, estimates from July 2023*^19,20^ |
| Boosted | 15 | 41.7 | 60 | 76.5 | 75.3 | CDC national vaccine administration coverage data^21^ |
| Primary vaccine series only | 43.5 | 37 | 28.8 | 19.8 | 13.6 |  |

*National serologic surveys with the nucleocapsid antibody (suggesting prior infection, not vaccine-induced antibody response). Data originally accessed at <https://covid.cdc.gov/covid-data-tracker/#pediatric-seroprevalence> and <https://covid.cdc.gov/covid-data-tracker/#nationwide-blood-donor-seroprevalence-2022>.

**Table A4: Estimated age-specific prevalence (%) of prior immune history in the United States, July 2023**

| **Immunity status** | **0-17 years** | **18-29**  **years** | **30-49**  **years** | **50-64**  **years** | **65+**  **years** |
| --- | --- | --- | --- | --- | --- |
| Immune naive | 0 | 0 | 0 | 0 | 0 |
| Infected | 8.6 | 4.6 | 2.6 | 2.2 | 1.6 |
| Reinfected | 33.2 | 17.7 | 10.1 | 8.5 | 6 |
| Infected + Vaccinated | 12.1 | 10 | 7.4 | 4.8 | 3.5 |
| Reinfected + Vaccinated | 30 | 24.6 | 18.5 | 11.8 | 8.6 |
| Infected + Boosted | 6.6 | 17.5 | 23.2 | 27.4 | 27.2 |
| Reinfected + Boosted | 7.4 | 19.4 | 25.8 | 30.4 | 30.3 |
| Boosted | 0.9 | 4.3 | 10 | 13.1 | 20.8 |
| Vaccinated | 1.2 | 1.9 | 2.4 | 1.8 | 2 |
| Prior infection | 97.9 | 93.8 | 87.6 | 85.1 | 77.2 |
| Boosted | 14.9 | 41.2 | 59 | 70.9 | 78.3 |
| Fully vaccinated only | 43.3 | 36.5 | 28.3 | 18.4 | 14.1 |

We assigned each individual a time since their most recent COVID-19 event (vaccination or infection) by generating separate distributions for vaccination and infection timing, then taking the most recent event. We assumed vaccine timing was independent of recent infection history, potentially overestimating impact among recently infected individuals, though this effect is likely modest given waning immunity. For time since vaccination, we used national CDC data on cumulative daily vaccine doses (primary series completion, 1 booster, 2+ boosters) from December 15, 2020 to July 1, 2023 (Figure A1A). Vaccination timing was assigned based on vaccine status: primary series recipients sampled from the primary distribution, while boosted individuals sampled from booster distributions conditional on their booster count (1 booster vs 2+ boosters). For individuals with prior infection, we simulated time since last COVID-19 infection by sampling from the distribution of estimated cases over time [accessed on <https://covidestim.org/>, which is a continuation of Klaassen et al.^18^] (Figure A1B). We constrained sampling to January 1, 2022 through July 1, 2023 to focus on the Omicron era. For reinfected individuals, we sampled their most recent infection from the period between their first infection date and July 1, 2023.

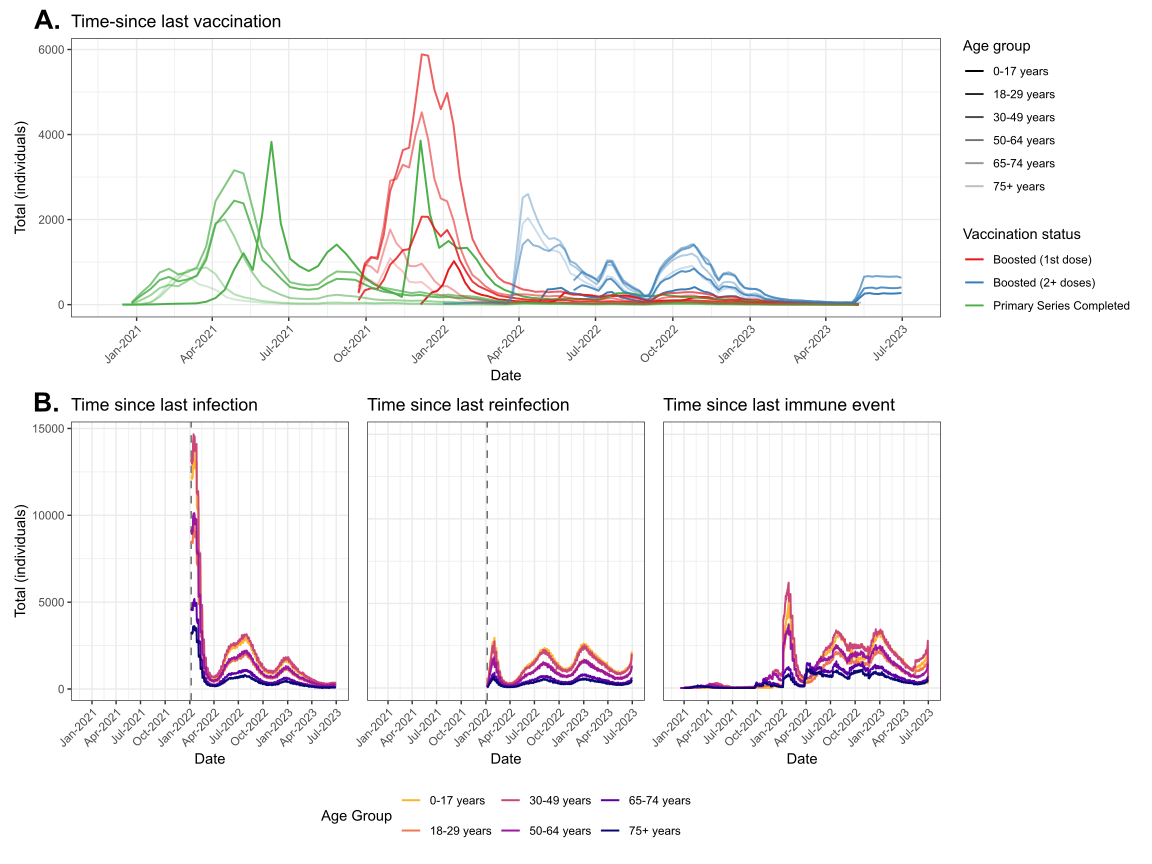
**Figure A1: Distributions used to assign time since last COVID-19 event in the simulated population.** (A) COVID-19 vaccine administration by vaccination status and age group. (B) COVID-19 cases: initial infections (left), reinfections (middle), and combined distribution for sampling time since last immune event (right). Colors represent vaccination status (A) or age groups (B); transparency represents age groups (A).

We initialized the number of infectious individuals in each age group as:

$$I_{i}\left( 0 \right)=5\left( \frac{1}{\theta_{i}}I_{severe,i}\left( 0 \right) \right)$$

where $I_{severe,i}\left( 0 \right)$ is the observed daily severe incidence in age group $i$ in early July 2023, $\theta_{i}$ is the age-specific severity ratio, multiplied by a 5-day infectious period.

We then solved for $\beta_{i}$ analytically using the force of infection equation, so that the initial force of infection reproduced observed age-specific hospitalization rates in early July 2023. Because we did not model pediatric COVID-19 hospitalizations as discussed in the Discussion, we set $\beta₀₋₁₇$ equal to $\beta₁₈₋₂₉$. Because $\betaᵢ$ is solved from a single time point and held fixed thereafter, and $\phi$ is a single scalar applied across all age groups, neither can accommodate drift in the age relationship of incidence over the 22-month period. Preliminary runs showed some overestimation of incidence in adults aged 50–64 years, and we therefore applied a fixed 0.80 multiplier to $\beta₅₀₋₆₄$, held constant across all simulations. No other age group was adjusted.

##### $\boldsymbol{\betaᵢ}$ is solved so that the initial force of infection reproduces observed age-specific hospitalization rates at $\boldsymbol{\kappa}_{\boldsymbol{0}}\boldsymbol{= 1}$, whereas $\boldsymbol{\kappa}\boldsymbol{ₜ}$ is estimated to reproduce the observed trajectory of incidence over time. These are distinct requirements on the product $\boldsymbol{\kappa}\boldsymbol{ₜ}\boldsymbol{\betaᵢ}$, and the calibrated $\boldsymbol{\kappa}\boldsymbol{ₜ}$ therefore absorb a scaling offset relative to the analytic solution. The absolute magnitude of $\boldsymbol{\kappa}\boldsymbol{ₜ}$is not interpretable as a transmission probability; only relative variation across months is meaningful.

##### Model calibration

We calibrated monthly transmission coefficients $\kappa_{1}-\kappa_{22}$ and a single severity scale parameter $\phi$ to observed weekly age-stratified COVID-19 hospitalization incidence data in adults 18+ years old from July 2023 through April 2025. Given the high dimensionality of this parameter space and computational constraints from stochastic model simulations, calibration proceeded in sequential stages: a preliminary grid search providing starting values for subsequent steps, calibrating a 10-month pre-study period (July 2023-April 2024) whose parameters were fixed thereafter, and two six-month phases during the study period. Phase boundaries were placed at the late-October seasonal trough so that each phase spans a single epidemic wave: the summer 2024 wave (May–October) in phase 1 and the winter 2024–25 wave (November 2024–April 2025) in phase 2. We used Approximate Bayesian Computation Sequential Monte Carlo (ABC-SMC) calibration using the Del Moral adaptive algorithm^22^ as implemented in the EasyABC R package^23^.

We first conducted a coarse grid search advancing sequentially through the time series in overlapping blocks of one to three months, evaluating adjacent monthly transmission coefficients together with $\phi$ and scoring parameter sets by sum of squared errors against age-specific incidence. This established plausible ranges spanning epidemic extinction to explosive growth, constraining $\phi$ so the severity in adults 50-64 years old was less than or equal to that observed earlier in the pandemic (Section S1).

The pre-study period entailed calibrating $\kappa₁-\kappa₁₀$ and the severity parameter $\phi$ (Table A5). We ran a preliminary ABC-SMC, then refined its estimates by a coarse search until simulated cumulative hospitalization incidence fell within 30% of observed values for adults 18-29 years, 20% for adults aged 30–49 years, and 10% for adults aged ≥50 years. More lenient thresholds were used for younger groups due to much lower absolute numbers. We simulated stochastic realizations of the pre-study period, retained the end states at the end of the pre-study period (day 304), including each individual’s immune history and infection status, as the initial condition for all subsequent calibration and analysis.

**Table A5: Pre-study period parameters**

| $\kappa_{1}$ | $\kappa_{2}$ | $\kappa_{3}$ | $\kappa_{4}$ | $\kappa_{5}$ | $\kappa_{6}$ | $\kappa_{7}$ | $\kappa_{8}$ | $\kappa_{9}$ | $\kappa_{10}$ | $\phi$ |
| --- | --- | --- | --- | --- | --- | --- | --- | --- | --- | --- |
| 0.49 | 0.43 | 0.43 | 0.50 | 0.63 | 0.67 | 0.65 | 0.65 | 0.60 | 0.56 | 0.0115 |

Study-period calibration used ABC-SMC with 200 particles per generation and α = 0.9, and a target tolerance of ε = 35 (based on preliminary runs, where tolerance had flattened) on the Euclidean distance between simulated and observed summary statistics after normalization by their standard deviation across the initial generation; this internal normalization accounts for differences in magnitude across age groups without manual weighting. For each phase, summary statistics comprised COVID-19 hospitalization incidence per 100,000 persons aggregated into seven four-week intervals for each of five adult age groups (18–29, 30–49, 50–64, 65–74 and 75+ years), together with cumulative hospitalization incidence by age over the phase window (40 statistics in total). Priors were independent uniform distributions of half-width 0.07 centered on the grid-search estimates, chosen to span the plausible range identified in the grid search while remaining computationally feasible. Simulated statistics were averaged over two stochastic replicates per particle in phase 1; a paired comparison of one- versus two-replicate runs showed no material difference in the accepted parameter sets, so phase 2 used a single replicate.

Phase 1 estimated $\kappa₁₁-\kappa₁₆$ (May–October 2024), with tolerance of 34.4 reached after 32 generations. We filtered the final generation to particles whose cumulative hospitalization incidence fell within 30% of observed values for the two youngest age groups and within 20% for the three oldest, yielding 32 unique accepted parameter sets. Phase 2 estimated $\kappa₁₇-\kappa₂₂$ (November 2024–April 2025) conditional on these, with tolerance of 34.0 reached after 34 generations. Rather than fixing phase-1 values at a point estimate, we treated the identity of the inherited phase-1 particle as an additional parameter with a uniform prior, sampled and reweighted alongside the transmission coefficients within the SMC, so that phase-1 parameter and stochastic uncertainty propagated into the phase 2.

We then applied the same acceptance criteria to cumulative hospitalization incidence over the full study period to ensure goodness-of-fit. Importantly, monthly transmission coefficients are not independently identified: a higher value in one month can be compensated by a lower value in the next, so the data constrain the joint trajectory rather than each coefficient separately. Sampling from each coefficient's marginal distribution independently would therefore generate combinations that were never accepted and that do not reproduce observed incidence. We consequently retained complete parameter sets rather than resampling individual coefficients, which limits the number of distinct parameterizations to those meeting the acceptance criteria. These criteria are stringent, and a single run yielded few sets meeting them: seven in the final calibration run (200 particles) and five in an independent earlier run (100 particles). Both runs used identical priors, summary statistics, and algorithm settings and therefore explored the same parameter space, and retained sets from both were held to the same acceptance criteria. We pooled these accepted sets, using five from each run. These ten sets were treated as equally likely and each simulated ten times, yielding 100 stochastic iterations per scenario.

The ten sets of monthly transmission coefficients used in projections are shown in Figure A2, together with the prior bounds, and the age-stratified model fits are shown in Figure A3. Note that the largest relative error (36.5%, ages 30–49 years) corresponds to 9.5 hospitalizations per 100,000 against an observed 26.0 per 100,000 (Figure A4). Also note that COVID-NET reports hospitalization rates for adults aged 18–49 years as a single category, which we assigned to both the 18–29 and 30–49 year groups; observed incidence is therefore identical for these two groups. Because the model generates an age gradient within 18–49 years that the surveillance data cannot resolve, this gradient appears as error in one group or the other. The underestimate in adults aged ≥75 years is consistent with the structural limitations described above (Section S3, Model Initialization). This underestimation makes estimated hospitalizations averted in older adults conservative.

ABC-SMC yields an approximate posterior distribution. The additional acceptance criterion we applied is not part of that procedure, so the retained sets are a filtered subset rather than a posterior sample.

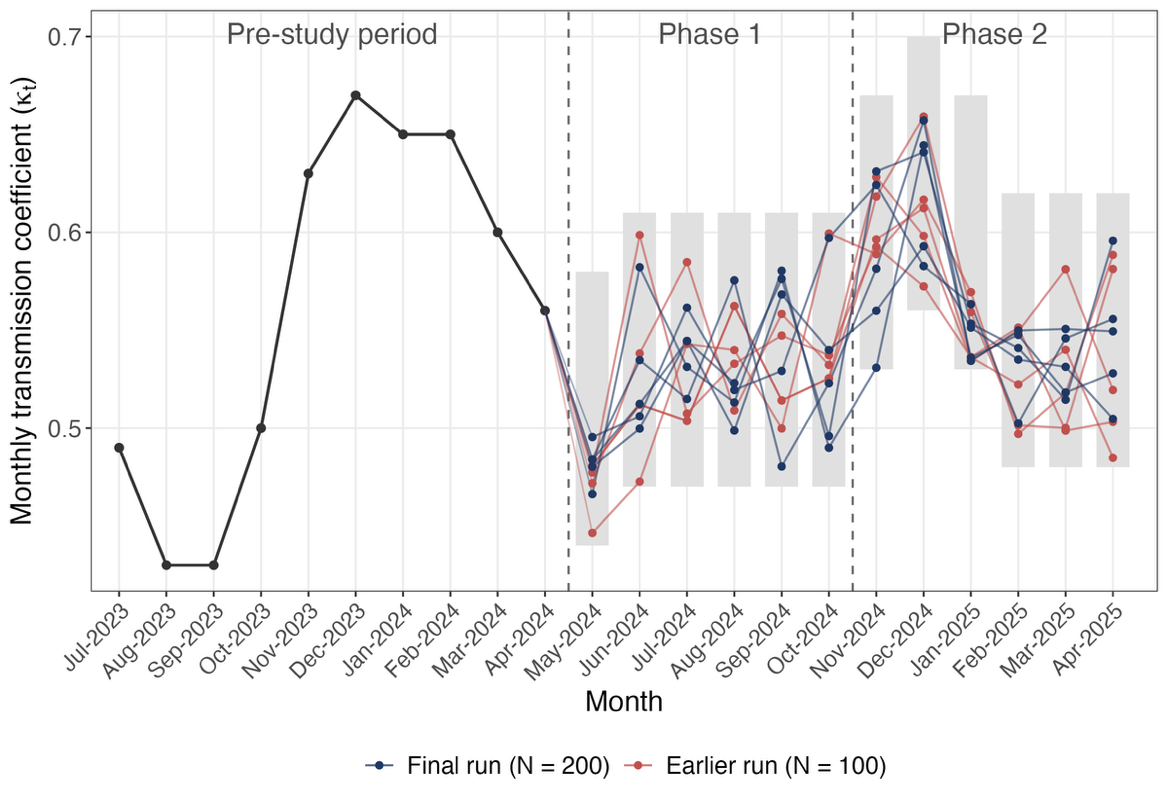

**Figure A2: Monthly transmission coefficients (κₜ) estimated by ABC-SMC.** Points and connecting lines show the ten accepted parameter sets used in model projections, colored by calibration run. Grey bands show the uniform prior support (± 0.07 around grid-search estimates). The dashed line marks the boundary between phase 1 (κ₁₁–κ₁₆) and phase 2 (κ₁₇–κ₂₂) of calibration.

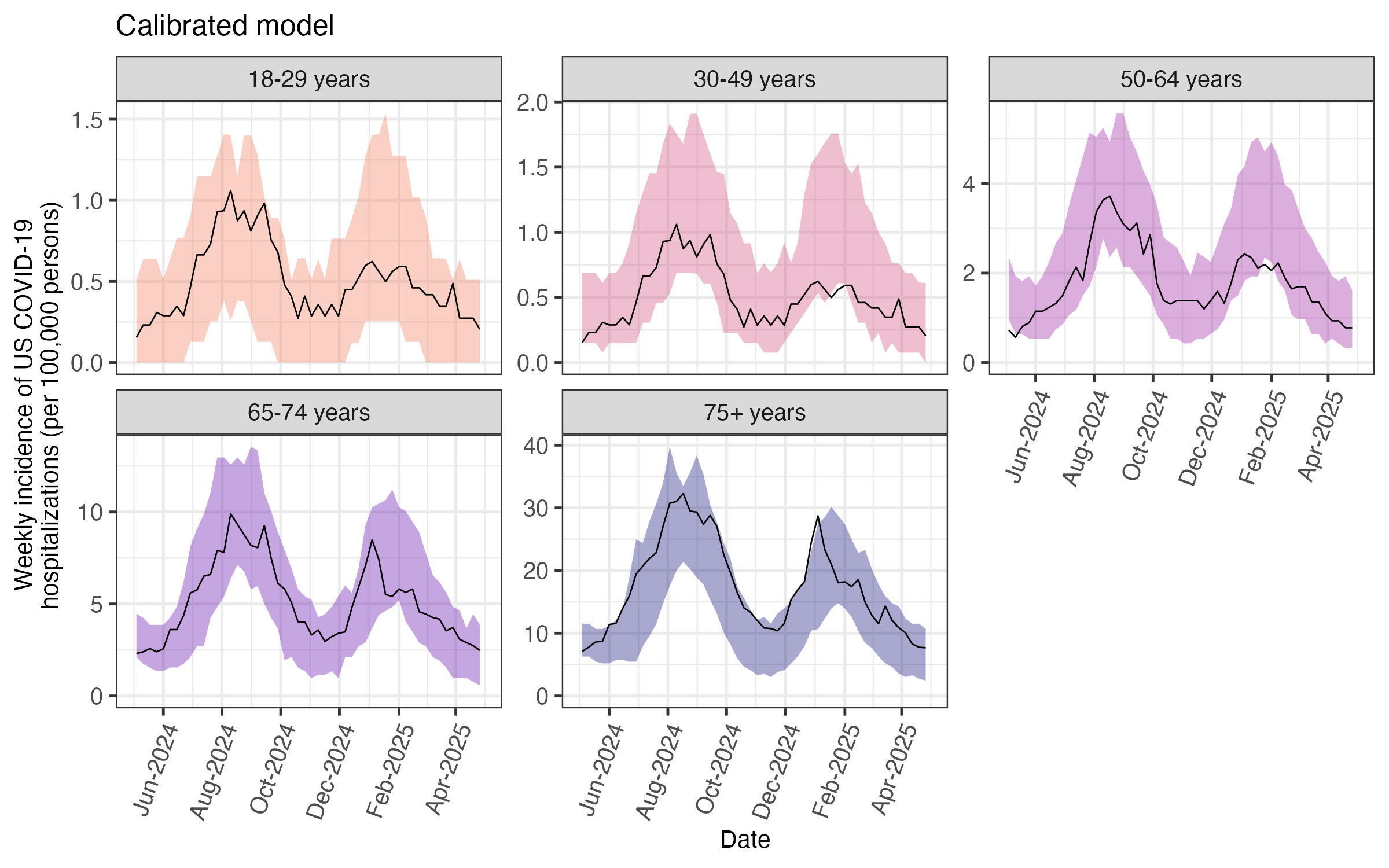

**Figure A3: Calibrated model fits compared to observed hospitalization incidence by age.** Black lines show observed weekly hospitalization rates per 100,000 persons. Shaded regions represent 5th–95th percentile of model predictions from 100 stochastic simulations using 10 calibrated parameter sets.

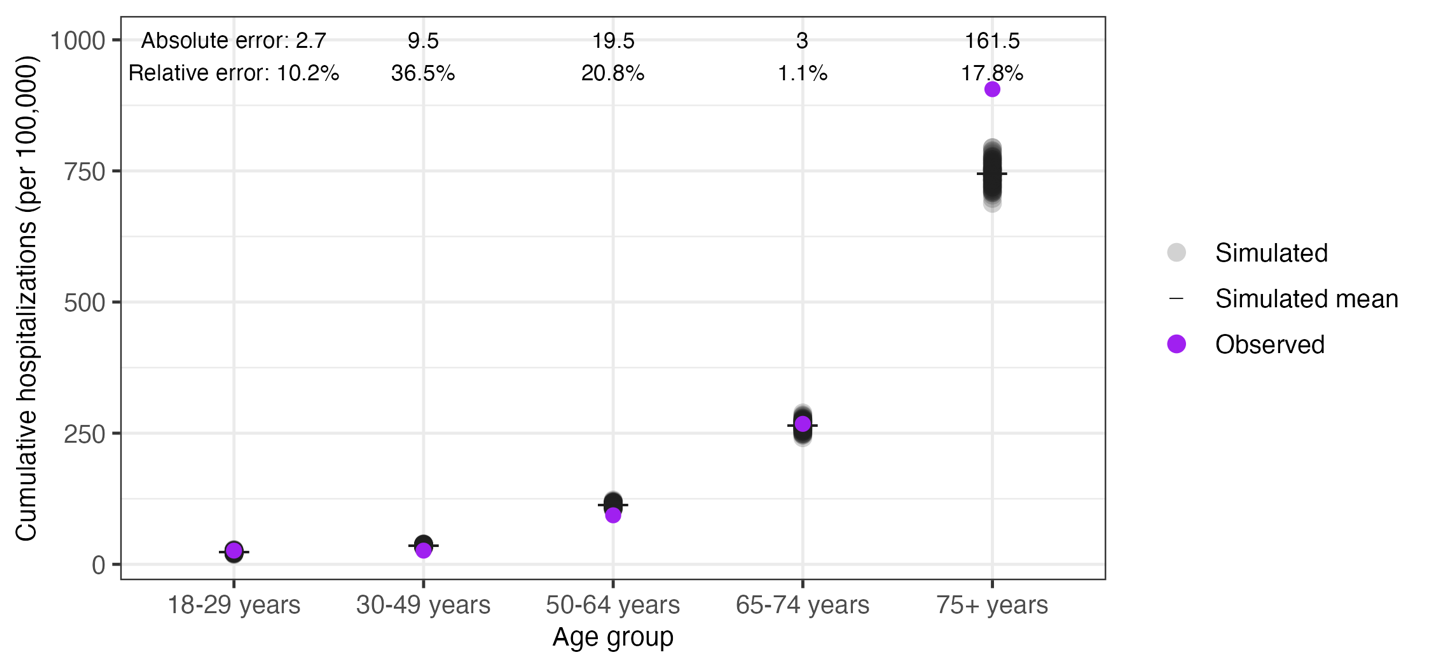

**Figure A4: Calibrated model fits with cumulative COVID-19 hospitalizations over the study period, model versus estimated observed values by age group.** Purple points show estimated observed cumulative hospitalizations per 100,000 persons over May 2024–April 2025. Grey points show each of 100 stochastic model iterations; black crossbars show their mean. Annotations give the absolute error (hospitalizations per 100,000) and relative error (%) of the model mean relative to the estimated observed value.

##### Model validation

Calibration targets comprised age-stratified hospitalization incidence in adults only. Risk-group-specific incidence was not among the summary statistics used to calibrate the study period, but did contribute to model initialization, used to set the age- and risk-specific severity ratios $\thetaᵢ,ᵣ$. Therefore, agreement by risk group over the study period tests whether these initial ratios propagate correctly under group-specific vaccination coverage and waning immunity, rather than providing independent validation. Simulated incidence broadly captured observed trends (Figure A5). In some cases, cumulative incidence estimates moderately differed from observed values. For example, cumulative incidence was overestimated in immunocompromised adults (the largest discrepancy in the model) and underestimated in higher-risk adults in the age groups carrying most of the burden (magnitudes reported in Figure A6; population-weighting these values by the prevalences in Table S1 reproduces the age-level observed incidence). This reflects how severity and protection are sourced for these groups. The severity ratios $\thetaᵢ,ᵣ$ derive from surveillance data in which immunocompromised status is defined broadly, whereas protective effectiveness against hospitalization is parameterized from studies of moderate to severe immunocompromising conditions (Sections S1, S2). The model therefore combines the relative severity of a broader, lower-risk group with the reduced vaccine response of a narrower, higher-risk one, over-predicting hospitalizations in immunocompromised adults; because βᵢ is solved to reproduce age-level incidence, this over-attribution is offset by under-attribution to higher-risk adults. The immunocompromised share of hospitalizations in the reference data is also assumed constant across the study period from a single published estimate, so these data cannot establish whether that share changed. Absolute risk estimates should therefore be treated as an upper bound in immunocompromised adults and a lower bound in higher-risk adults, while comparisons across vaccination timing and strategy within a risk group are unaffected, since the same severity ratios apply to every scenario.

We additionally compared two model outputs against external estimates not used in calibration. First, the relative effectiveness of an additional vaccine dose implied by our fitted waning curves was consistent with published estimates stratified by prior infection status (Figure S2, Section S2). Additionally, projected hospitalizations under a counterfactual scenario without vaccination exceeded observed incidence throughout the study period by a plausible margin (Figure A7). Second, the proportion of infections that were non-severe in the calibrated model (99.8%) compared to published estimates for the Omicron era (97.9%^24^), equivalent to an infection-severe rate of 0.2% versus 2.1%. Two factors contribute: first, the published estimates are likely biased due to under-ascertainment of asymptomatic infections, which would narrow the apparent difference; second, hospitalization is a stricter endpoint than the severe clinical disease measured in that study, which would further reduce the difference.

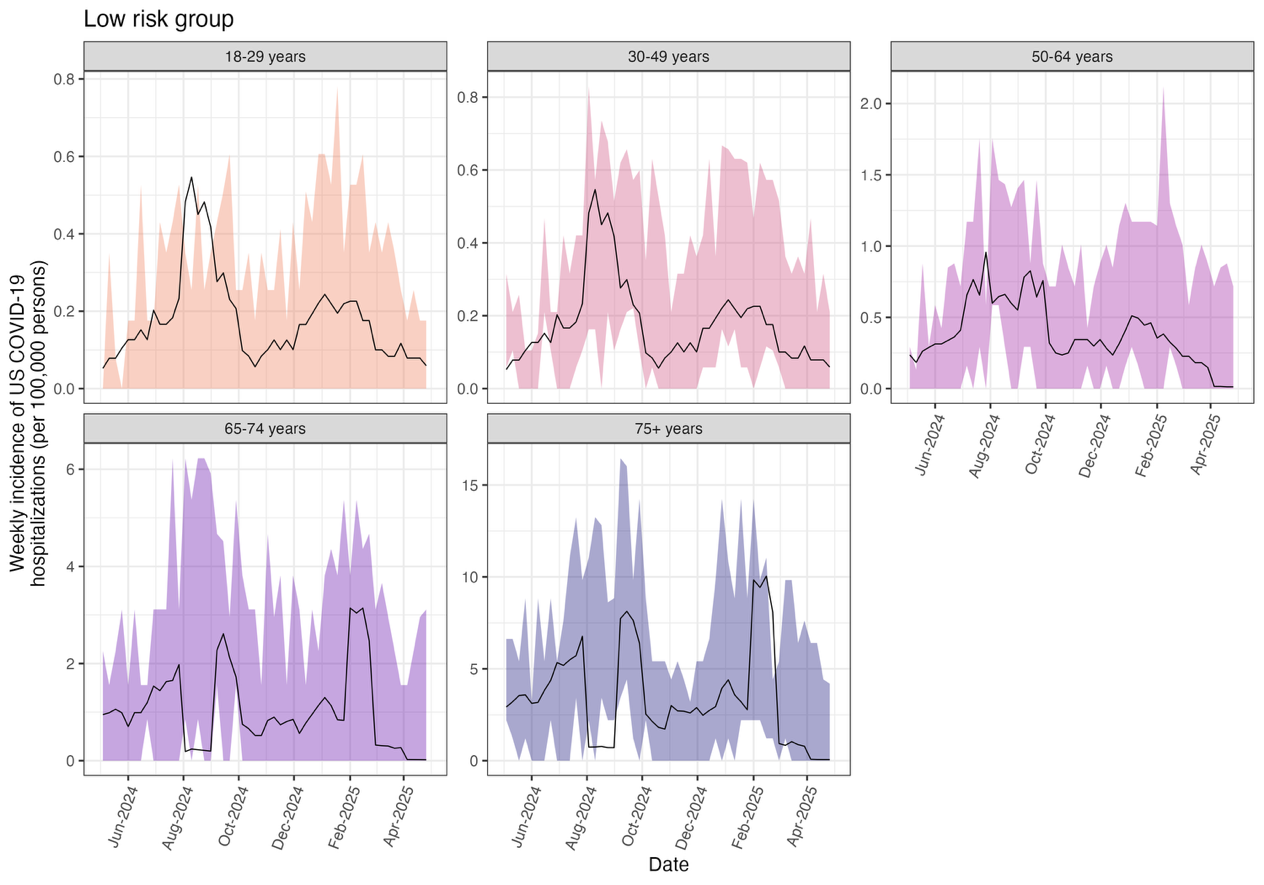

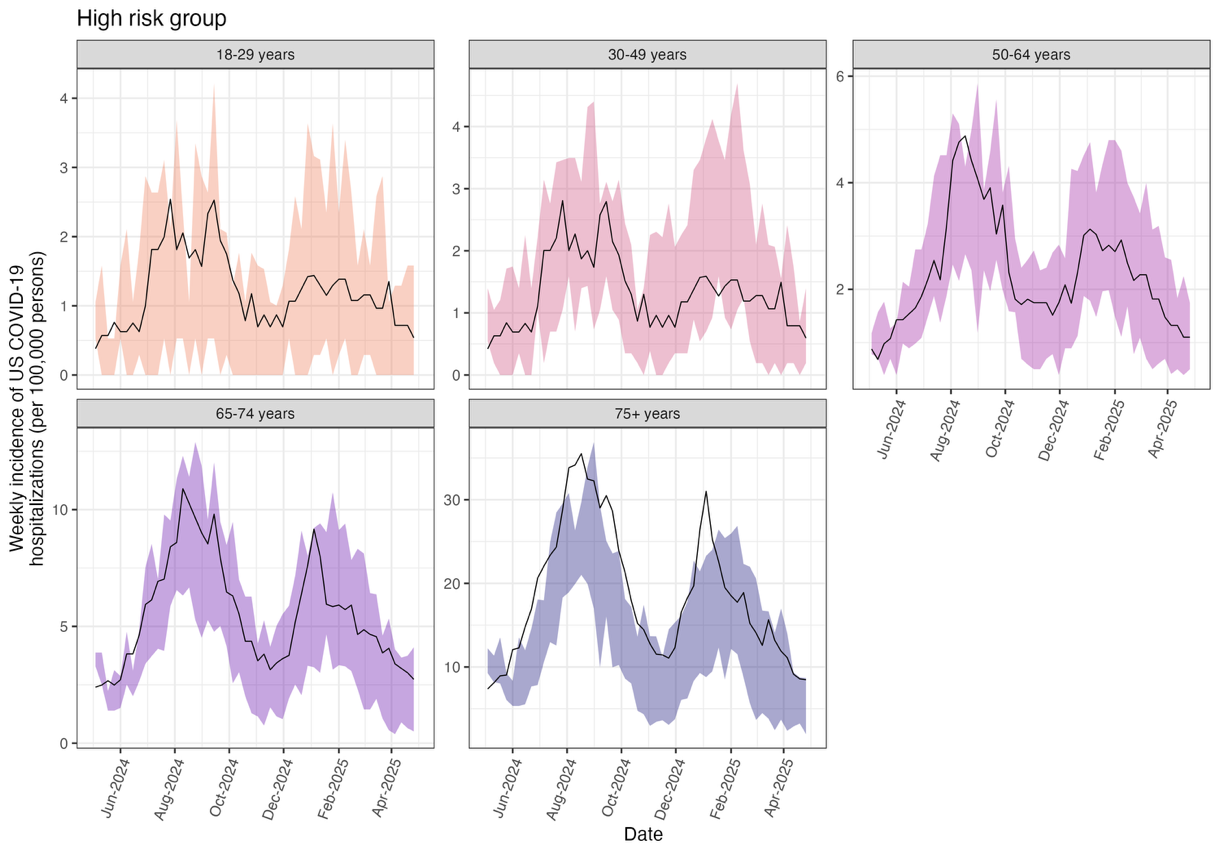

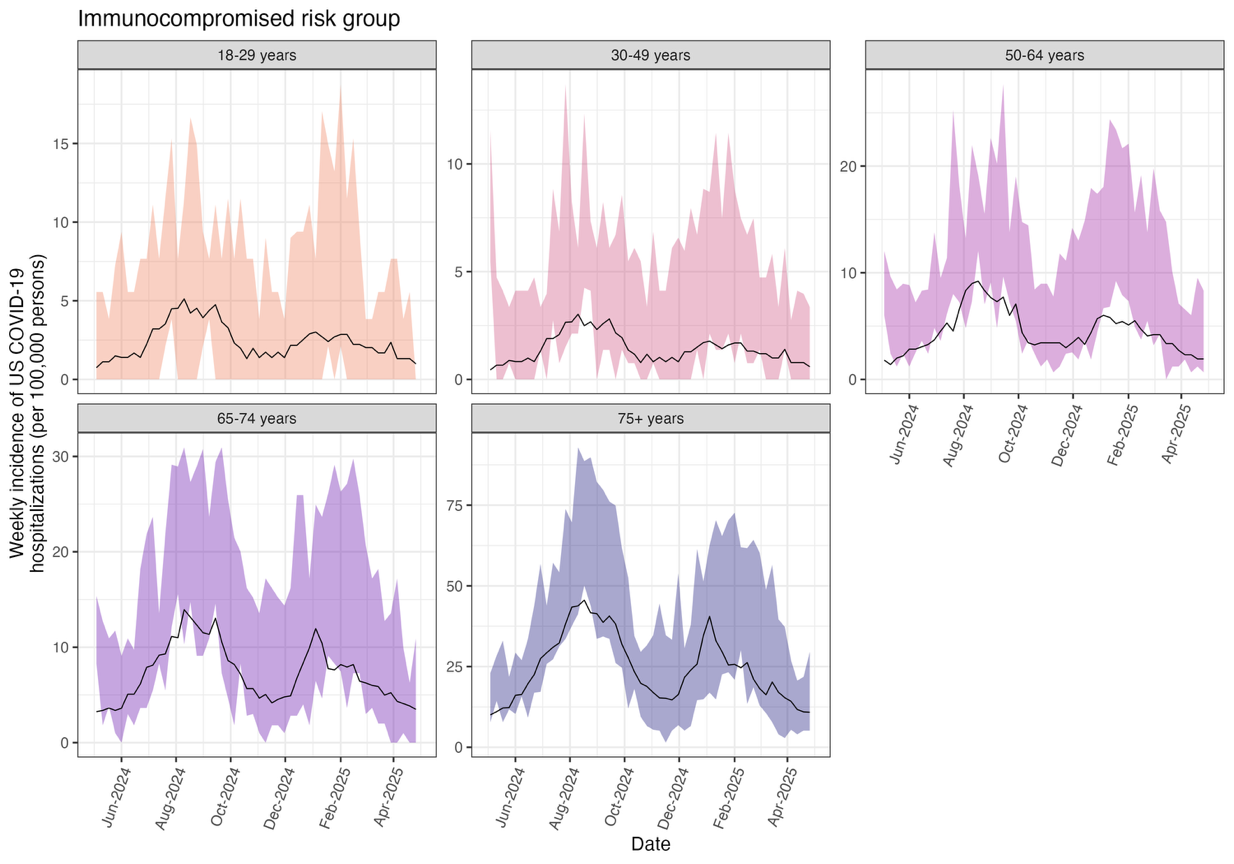

**Figure A5: Model fits against observed hospitalizations in age-risk stratified groups.** Black lines show observed hospitalization rates per 100,000 persons for low risk (top), high risk (middle), and immunocompromised individuals by age group (bottom). Shaded regions represent 5th–95th percentile of model predictions from 100 stochastic simulations. Model was calibrated to overall age-specific hospitalization data; this comparison validates performance when stratified by risk group. Note the differing y-axis scales across panels.

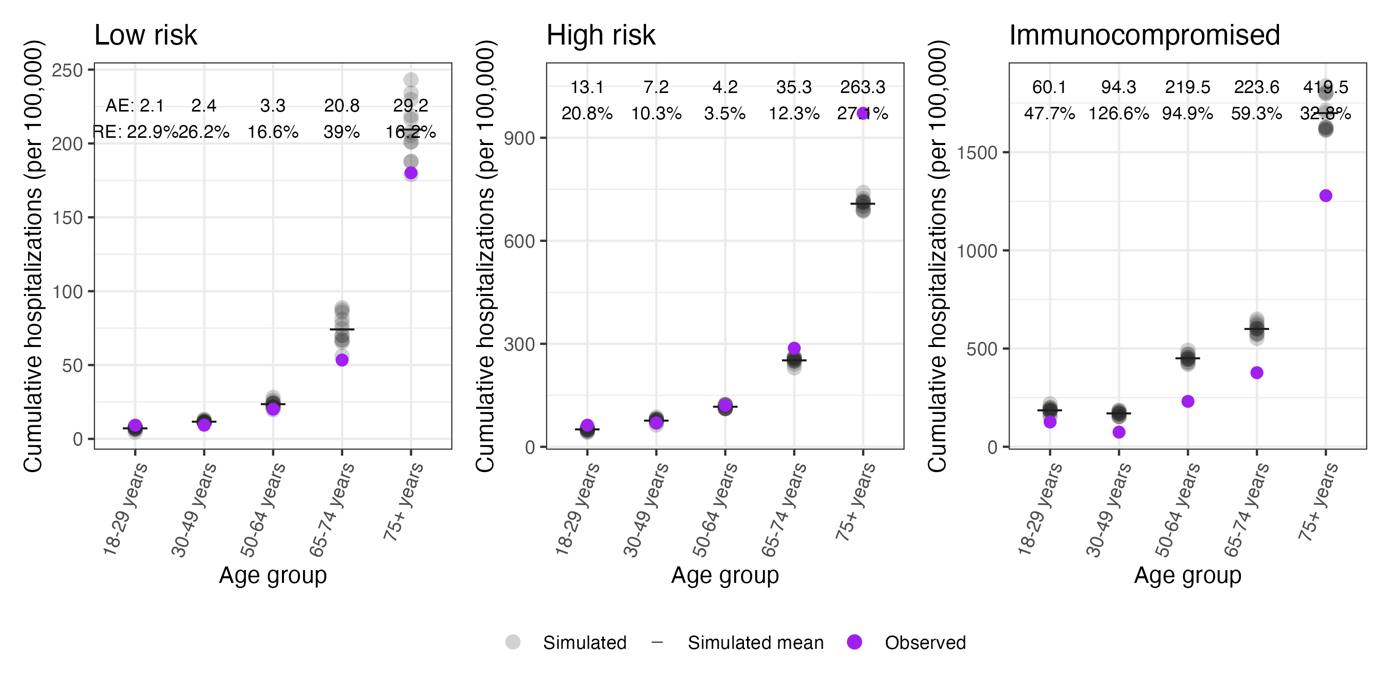

**Figure A6: Simulated versus observed cumulative COVID-19 hospitalizations by age-risk group, May 2024 to April 2025.** Observed (purple circle) and stochastic simulated (individual model run: grey circle, mean: black bar) cumulative hospitalizations per 100,000 persons. Annotations give absolute error (AE) and relative error (RE) comparing simulated mean to observed values. Observed values by risk group were estimated rather than measured directly (Section S1). Note the differing y-axis scales across panels.

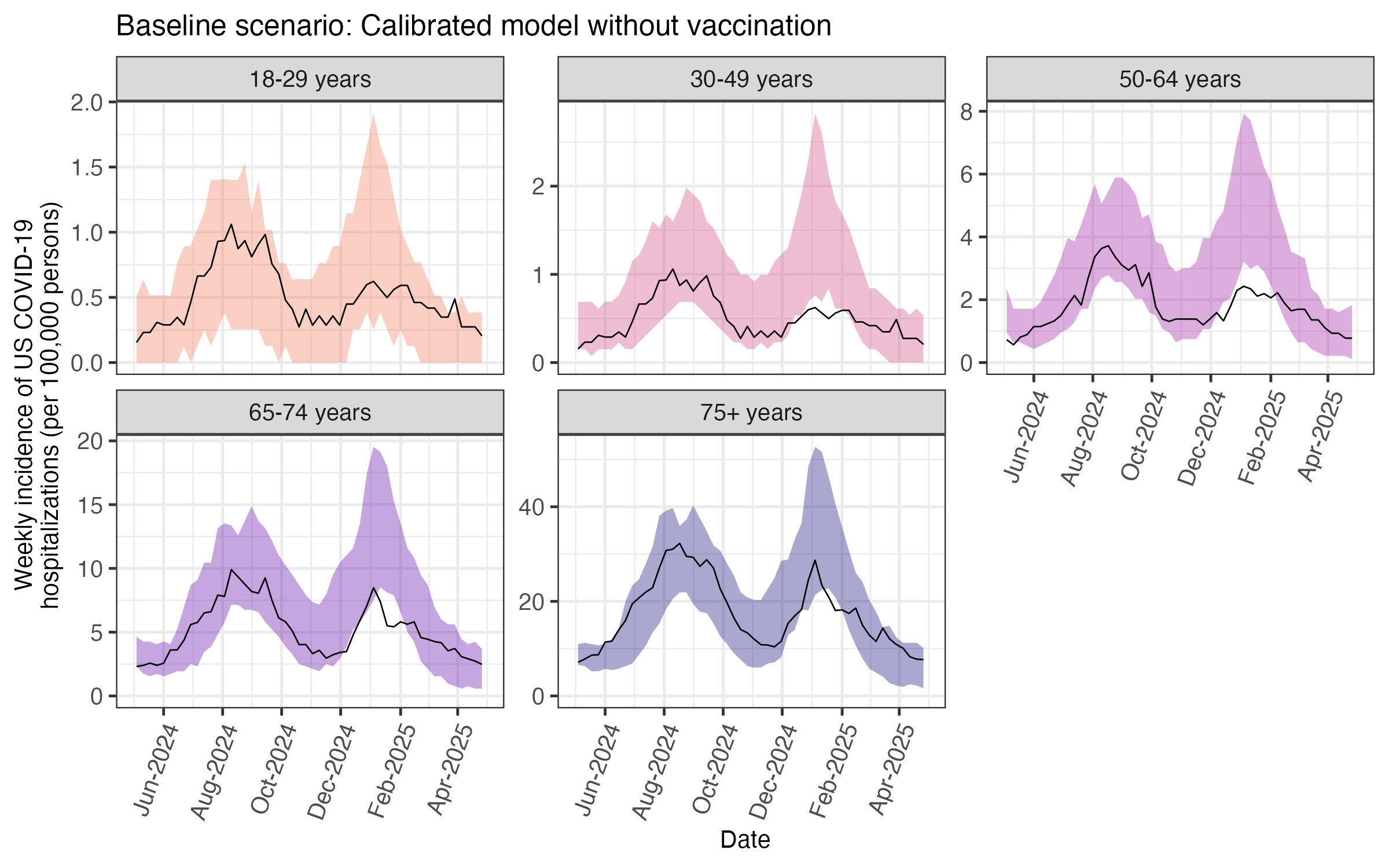

**Figure A7: Projected COVID-19 hospitalization incidence by age group with no additional vaccination during the study period.** Black lines show observed hospitalization rates per 100,000 persons, which reflect actual vaccination. Shaded regions show model projections (5th–95th percentile across 100 stochastic simulations) of rates that would have occurred without additional vaccination. Divergence between the two therefore represents the estimated impact of vaccination and is not a measure of model fit. This scenario serves as the reference for absolute and relative risk reduction calculations (Section S4) and corresponds to the reference row in Table S14.

#### Section S4: Study outcomes definitions and calculations

We focused on COVID-19 related hospitalizations rather than mortality because hospitalizations occur more frequently, are better captured by surveillance systems, and provide more stable estimates across age and risk strata.

Each vaccination strategy run was paired with the matched no vaccination administration reference run that shared the same random seed. In addition to risk of COVID-19 hospitalization (described in the Main Text), we calculated three quantities. These are deliberately defined on different denominators, reflecting two distinct questions: the benefit to an individual who is vaccinated, and the impact of a strategy on the population as a whole.

Absolute risk reduction (ARR) was defined, within each age–risk group, as the difference between severe risk across the entire group over the 12-month study period in the no-vaccination scenario and severe risk among vaccinated individuals in the vaccination scenario. We define vaccinated individuals here as those who received all doses for which they were eligible under the given strategy. Number needed to vaccinate (NNV) was calculated as $1/ARR$, giving the number of persons vaccinated (not doses administered) required to prevent one hospitalization. ARR and NNV therefore describe benefit among those vaccinated, consistent with conventional use of number needed to treat.

Relative risk reduction (RRR) was defined as the proportional reduction in population severe risk across the entire age–risk group under vaccination relative to the matched no-vaccination scenario, $\mathrm{RRR}=\left( R_{b}-R_{v} \right)/R_{b}$, where $R_{b}$ and $R_{v}$ are population severe risks without and with vaccination, respectively. RRR is defined on the total population so that it reflects both direct protection of vaccinated individuals and indirect protection of those unvaccinated.

Both metrics are calculated from paired simulations differing only in vaccination. In groups with low absolute risk or low vaccine coverage, the vaccination effect can be smaller than stochastic variation between paired runs, so individual simulations may show no benefit or apparent harm. This is why RRR uncertainty intervals include zero in some groups even where the corresponding ARR interval does not.

To assess broader societal impacts, we also estimated workdays lost due to infection and vaccination among adults 18-64 years old (Table A6). These estimates are conservative compared to other modeling studies (e.g., Fitzpatrick et al. assumed 5 days missed from school for mild illness and 10 days for severe illness or hospitalization^25^). We adjusted calculations by multiplying by 5/7 to account for weekends and multiplying by 0.96 to account for an estimated 4% unemployment rate^26^.

To evaluate if one vaccine strategy performed better than another, we compared the difference in outcomes between scenarios within matched simulations, pairing on seed and parameter set, because uncertainty intervals for individual scenarios reflect shared parameter uncertainty and frequently overlap. From this distribution of differences, a scenario was considered superior if it was more favorable than each alternative in a majority of paired simulations. To incorporate the relative magnitude as well as the direction of each difference, we also applied paired Wilcoxon signed-rank tests with Holm-adjusted p-values and report the number of pairwise comparisons reaching statistical significance in the Supplementary Tables.

**Table A6: Parameters for number of workdays missed due to SARS-CoV-2 infection or vaccine side effects**

| Event | Proportion | Number of workdays missed | Notes from references | Ref. |
| --- | --- | --- | --- | --- |
| Nonsevere infection, asymptomatic | 30% of nonsevere | 0 |  | ^27^ |
| Nonsevere infection,  mild symptoms | 50% of nonsevere | 0.5 |  | ^27^ |
| Nonsevere infection, moderate symptoms | 20% of nonsevere | 2.5 | Mean duration of absence in 2022-2023 was 5.8 days, chose parameter to reflect decreasing severity in recent years | ^27,28^ |
| Severe disease (hospitalization) | 100% of severe | 7 | Omicron-phase stays were typically 5-8 days | ^29^ |
| Vaccination | 11% of vaccinated | 1 | 11% had side-effects from boosters typically lasting 1-2 days^30^, 17% of HCWs missed at least 1 day^31^ | ^30,31^ |

#### Section S5: Archetypes representing state-level heterogeneity

We characterized regional variation in COVID-19 seasonality using hospitalization data from 12 states in COVID-NET with complete data for May 2024-April 2025. For each state we calculated the ratio of cumulative hospitalization incidence in May–October 2024 to that in November 2024–April 2025, using all-ages rates reported directly by COVID-NET rather than age-stratified estimates scaled to US demographics. Ratios ranged from 0.83 in Michigan (favoring larger winter peak) to 3.40 in California (favoring larger summer peak), with a national ratio of 1.28 (Table A7).

#### Rather than fitting separate models to individual states, we constructed two archetypes to bracket this observed range, holding all other model inputs (population structure, vaccine coverage and uptake timing, and waning immunity) at their national values. Archetypes were generated by applying multiplicative adjustments to the calibrated monthly transmission coefficients $\boldsymbol{\kappa}\boldsymbol{ₜ}$(Table A8), yielding seasonality ratios of 2.83 for the summer-peak archetype and 0.91 for the winter-peak archetype. Archetype adjustments were selected to reproduce the seasonality of the informing states; total transmission was not constrained to match the national scenario. These archetypes represent plausible seasonality patterns spanning the range observed across states rather than models calibrated to any individual state; the informing states shown in Figure 3 and S4 are provided for reference.

**Table A7: State-level heterogeneity in timing of COVID-19 burden.** Ratio of cumulative hospitalization incidence in May–October 2024 versus November 2024–April 2025. Ratios >1 indicate summer-dominant burden; ratios <1 indicate winter-dominant burden. National US ratio is highlighted for reference.

| **Region** | CA | OR | UT | NY | CO | US | GA | NM | MD | MN | CT | TN | MI |
| --- | --- | --- | --- | --- | --- | --- | --- | --- | --- | --- | --- | --- | --- |
| **Ratio of cumulative incidence** | 3.40 | 2.61 | 1.66 | 1.34 | 1.31 | 1.28 | 1.18 | 1.10 | 1.08 | 1.04 | 0.98 | 0.98 | 0.83 |

**Table A8: Multiplicative adjustments to transmission coefficients (**$\kappaₜ$**) for each archetype.**

|  | **May** | **Jun** | **Jul** | **Aug** | **Sep** | **Oct** | **Nov** | **Dec** | **Jan** | **Feb** | **Mar** | **Apr** |
| --- | --- | --- | --- | --- | --- | --- | --- | --- | --- | --- | --- | --- |
| Archetype 1: Summer-peak | +10% | +10% | +10% | +15% | +22% | +20% | — | −10% | −10% | −10% | −10% | −10% |
| Archetype 2: Winter-peak | — | — | — | — | +10% | +10% | +15% | +22% | +27% | +27% | +22% | +22% |

#### Section S6: Sensitivity analyses

We conducted sensitivity analyses varying the waning immunity (protective effectiveness) curves, vaccine coverage, the vaccine uptake pattern, and the evaluation window.

For waning immunity, we repeated the primary analysis using faster waning of protection against hospitalization (i.e., waning model fit to 95% lower bound of literature estimates, Figure S1) while holding protection against infection at baseline values. The ranking of the top strategies was unchanged, with July-timed universal annual vaccination remaining most favorable to reduce hospitalization burden in adults 65+ years (Figure A8).

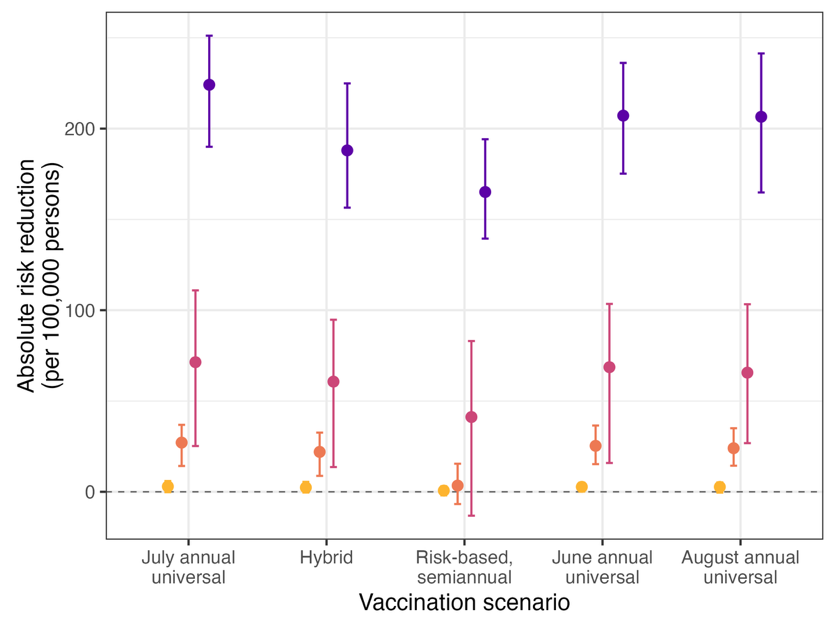

**Figure A8: Sensitivity analysis using faster waning of protection against COVID-19 hospitalization.** Absolute risk reduction per 100,000 persons relative to no additional vaccination during the study period, by vaccination scenario and age-risk group: ≥65 years (purple), 18–64 years immunocompromised (pink), 18–64 years high risk (orange), and 18–64 years low risk (yellow). Protection against hospitalization followed the 95% lower bound of the literature-based estimates, while protection against infection was held at baseline values. Universal annual scenarios assume one-month uptake beginning in the indicated month; hybrid and risk-based semiannual vaccination assume historical fall timing. All scenarios assume historical vaccine coverage. Points show medians with 95% uncertainty intervals across 100 simulation runs.

For vaccine coverage, we considered a higher coverage scenario in the Main Text when comparing vaccine strategies varying by eligibility and frequency (Table S2). We also considered reduced coverage by 5 and 10 absolute percentage points for vaccination occurring in June–August when comparing vaccine strategies by timing, for national results (Figure S6, Table S12) and for both state-level archetypes (Figures S8, S9).

For vaccine uptake, we compared timing under the historical four-month uptake pattern rather than one-month compressed uptake (Figure S5, Table S7); the uptake pattern did not meaningfully change results.

In the Main Text, outcomes were accumulated over the full 12-month study period (May 2024–April 2025). Because vaccines given earlier in that window have longer to accrue benefit, we also evaluated outcomes over the 180 days following each scenario's vaccination start. Neither window is neutral: the 12-month window equalizes calendar exposure but not time since vaccination, while the 180-day window equalizes time since vaccination but exposes scenarios to different epidemic conditions (July vaccination captures July–December; November captures November–April). We treat the 12-month window as primary, since an annual program is evaluated over the same calendar year regardless of when doses are administered.

Under the 180-day window, optimal timing at the national level shifted toward later months for adults ≥65 years: November vaccination achieved the lowest hospitalization risk, September the highest population-level absolute risk reduction, and August the highest absolute risk reduction among vaccinated individuals (Figure A9, Table S13). Similar patterns emerged in younger age groups, with August–November outperforming summer months. The advantage of summer timing therefore reflects greater coverage of the year's total burden rather than higher protection per unit time since vaccination. However, this comparison is confounded by different underlying seasonal risk patterns during each follow-up period which makes this comparison more challenging to interpret.

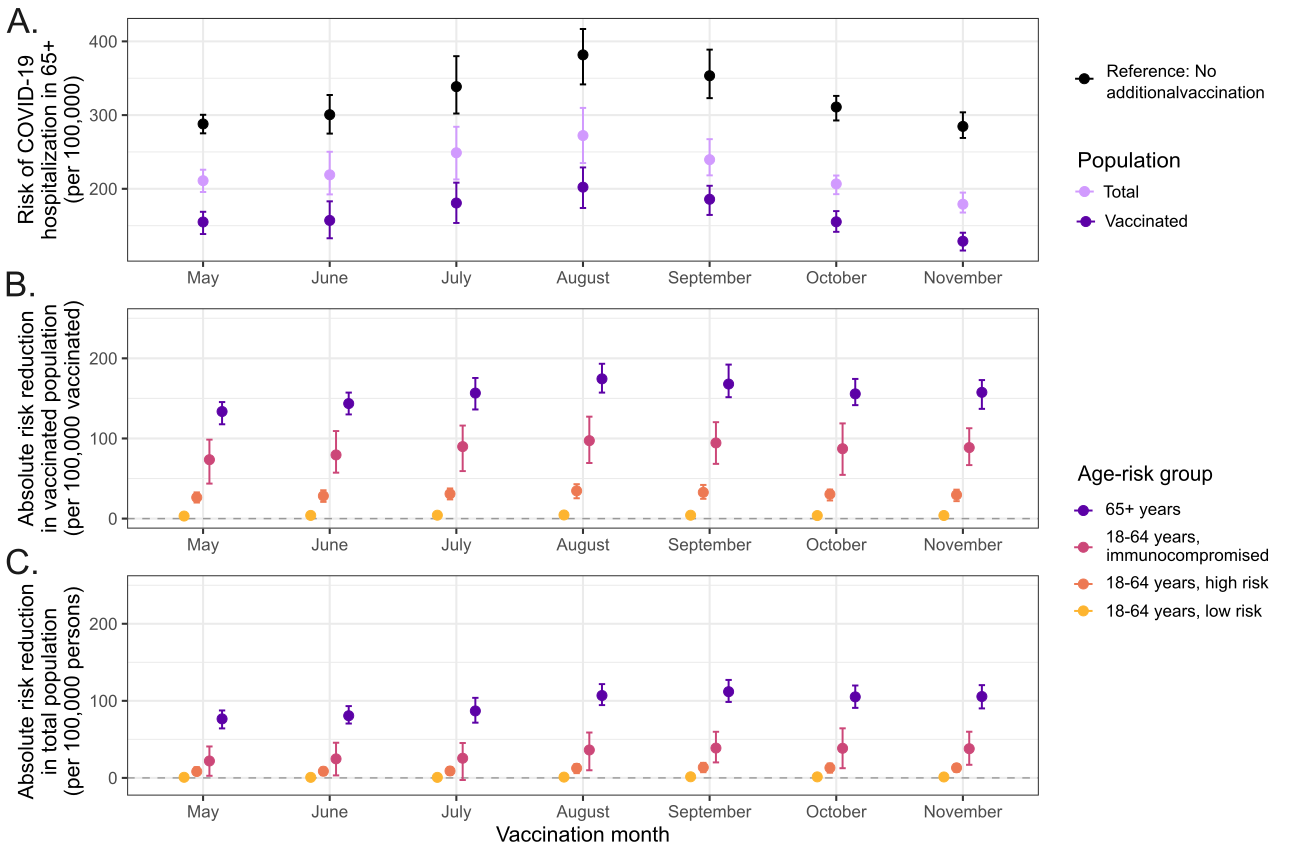

**Figure A9: Impact of vaccination timing with one-month compressed uptake over a 6-month follow-up window.** We modeled universal vaccination in individuals ≥6 months of age, with observed coverage levels but uptake compressed into a single month (x-axis label indicates the vaccination month). We assessed cumulative hospitalization risk over the 6-month period following the start of vaccination. This scenario differs from Figure 2 by using a shorter follow-up window rather than the full 12-month study period. (A) Annual hospitalization risk per 100,000 in adults ≥65 years, shown for vaccinated individuals (dark purple), the total population including unvaccinated (light purple), and a no-vaccination scenario (gray). (B) Absolute risk reduction per 100,000 among vaccinated individuals and (C) the total population, stratified by age-risk group: ≥65 years (purple), 18–64 years immunocompromised (pink), 18–64 years with comorbidities (orange), and 18–64 years low risk (yellow). Error bars represent 95% uncertainty intervals (i.e., 2.5^th^-97.5^th^ range of 100 stochastic simulations).

### Supplementary Tables and Figures

##### Table S1: Demographic characteristics of simulated population, by age and risk group.

References include the 2025 Census^32^, 2018 and 2021 National Health Interview Survey^6,33^, and CDC Chronic Disease Indicators^8^

| Risk group | Prevalence in US population (%) | Risk group prevalence in each age group (%) | Reference |
| --- | --- | --- | --- |
| **0-17 years** | **21.76** | -- | ^32^ |
| Low risk | 19.19 | 88.2 | -- |
| High risk | 2.00 | 9.2 | ^34^ |
| Immunocompromised | 0.56 | 2.6 | ^35^ |
| **18-29 years** | **15.70** | -- | ^32^ |
| Low risk | 11.40 | 72.6 | -- |
| High risk | 3.78 | 24.1 | ^6^ |
| Immunocompromised | 0.52 | 3.3 | ^33^ |
| **30-49 years** | **26.21** | -- | ^32^ |
| Low risk | 19.03 | 72.6 | -- |
| High risk | 5.71 | 21.8 | ^6,7^ |
| Immunocompromised | 1.47 | 5.6 | ^33^ |
| **50-64 years** | **18.68** | -- | ^32^ |
| Low risk | 6.83 | 36.6 | -- |
| High risk | 10.18 | 54.5 | ^6^ |
| Immunocompromised | 1.66 | 8.9 | ^33^ |
| **65-74 years** | **10.35** | -- | ^32^ |
| Low risk | 1.28 | 12.4 | -- |
| High risk | 7.95 | 77.0 | ^6^ |
| Immunocompromised | 1.10 | 10.6 | ^33,35^ |
| **75+ years** | **7.30** | -- | ^21,23^ |
| Low risk | 0.91 | 12.4 | -- |
| High risk* | 5.65 | 77.0 | ^5,6^ |
| Immunocompromised* | 0.78 | 10.6 | ^33,35^ |

*Assumed the same risk group prevalence as 65-74 years

**Table S2: Historical and higher vaccination coverage scenarios, by age and risk group.**

| Risk group | Historical coverage scenario  (%) | | | | Higher coverage scenario  (%) | | | |
| --- | --- | --- | --- | --- | --- | --- | --- | --- |
|  | 2023-2024 vaccine | | 2024-2025 vaccine | | 2023-2024 vaccine | | 2024-2025 vaccine | |
|  | Dose 0 | Dose 1 | Dose 2 | Dose 3* | Dose 0 | Dose 1 | Dose 2 | Dose 3 |
| **0-17 years** | **14.3** | **--** | **13.4** | **--** | Same as historical | **--** | **50** | **--** |
| Low risk | 13.1 | -- | 11.9 | -- |  | -- | 45.7 | -- |
| High risk | 23.5 | -- | 23.9 | -- |  | -- | 82.2 | -- |
| Immunocompromised | 23.5 | -- | 23.9 | -- |  | -- | 82.2 | -- |
| **18-29 years** | **11.3** | **--** | **9.7** | **--** |  | **--** | **60** | **--** |
| Low risk | 9.3 | -- | 7.6 | -- |  | -- | 49.2 | -- |
| High risk | 16.7 | 5.4** | 15.2 | 3.3** |  | 60 | 88.6 | 60 |
| Immunocompromised | 16.7 | 5.4 | 15.2 | 3.3 |  | 60 | 88.6 | 60 |
| **30-49 years** | **15.9** | **--** | **15.4** | **--** |  | **--** | **60** | **--** |
| Low risk | 13.1 | -- | 12.1 | -- |  | -- | 49.2 | -- |
| High risk | 23.5 | 5.4** | 24.3 | 3.3** |  | 60 | 88.6 | 60 |
| Immunocompromised | 23.5 | 5.4 | 24.3 | 3.3 |  | 60 | 88.6 | 60 |
| **50-64 years** | **21.7** | **--** | **24.1** | **--** |  | **--** | **70** | **--** |
| Low risk | 14.4 | -- | 14.7 | -- |  | -- | 46.4 | -- |
| High risk | 25.9 | 5.4** | 29.5 | 3.3** |  | 70 | 83.6 | 70 |
| Immunocompromised | 25.9 | 5.4 | 29.5 | 3.3 |  | 70 | 83.6 | 70 |
| **65-74 years** | **37.9** | **8.9** | **43.9** | **5.7** |  | **80** | **80** | **80** |
| Low risk | 22.3 | 8.9 | 23.3 | 5.7 |  | 47 | 47 | 47 |
| High risk | 40.1 | 8.9 | 46.8 | 5.7 |  | 84.7 | 84.7 | 84.7 |
| Immunocompromised | 40.1 | 8.9 | 46.8 | 5.7 |  | 84.7 | 84.7 | 84.7 |
| **75+ years** | **37.6** | **8.9** | **47.8** | **5.7** |  | **80** | **80** | **80** |
| Low risk | 22.1 | 8.9 | 25.4 | 5.7 |  | 47 | 47 | 47 |
| High risk | 39.8 | 8.9 | 50.9 | 5.7 |  | 84.7 | 84.7 | 84.7 |
| Immunocompromised | 39.8 | 8.9 | 50.9 | 5.7 |  | 84.7 | 84.7 | 84.7 |

Doses 0 and 2 represent the annual fall doses during the study period, doses 1 and 3 represent the semi-annual spring doses. Historical coverage assumptions are based on historical vaccine administration data in the United States from CDC^21,36–38^, with risk-stratified coverage estimated from published data^39^. The higher coverage scenario is hypothetical.

*The published data estimating coverage of the semiannual dose is from the 2023-2024 season, so we assume dose 3 coverage in the following year is the same as dose 1^37^. However, the study period only includes half the time the 4^th^ dose would have been distributed, so the coverage estimates reflect that difference.

**Assumed historical coverage since these age-risk groups were not eligible for semiannual vaccination historically but are included in some vaccination scenarios

##### Table S3: Summary of literature on vaccine effectiveness against COVID-19

| **Vaccine effectiveness end point** | **Type of vaccine effectiveness** | **Immunity type** | **Follow up time** | **References** |
| --- | --- | --- | --- | --- |
| Severe disease/  hospitalization |  | Hybrid immunity | 6 months  11 months  12 months | Bobrovitz et al.^40^  Carazo et al.^41^  Lee et al.^42^ |
|  | Absolute | Vaccine-induced  (no prior infection) | 8 months  16 months  4 months | Bobrovitz et al.^40^  Ferdinands et al.^11^  Link-Gelles et al.^43^ |
|  |  | Infection-acquired  (no prior vaccination) | 16 months  13 months  15 months | Bobrovitz et al.^40^  COVID Forecasting Team^44^  Lee et al.^42^ |
|  | Relative | Hybrid immunity | 10 months  10 months  8 months | Link-Gelles et al.^45^  (2023-2024 formulation)  Wiegand et al.^46^  (2024-2025 formulation)  Ma et al.^47^  (2024-2025 formulation) |
| Infection |  | Hybrid immunity | 6 months | Bobrovitz et al.^40^ |
|  | Absolute | Vaccine-induced  (no prior infection) | 8 months  7 months | Bobrovitz et al.^40^  Lau et al.^48^ |
|  |  | Infection-acquired  (no prior vaccination) | 12 months | Bobrovitz et al.^40^ |
|  | Relative | Hybrid immunity | 4 months  6 months | CDC MMWR^49^  (2023-2024 formulation)  Kirwan^50^  (2023-2024 formulation) |

##### Table S4: Vaccination strategies included in the model

| **Scenario** | **Description** |
| --- | --- |
| No additional vaccination (reference) | *Strategy 0:* No additional vaccination |
| Universal | *Strategy 1*: Annual vaccine 6+ months  *Strategy 2**: Annual vaccine 18+ years |
| Risk-based (traditional) | *Strategy 3*: Annual vaccine 65+ years and those with immunocompromising conditions  *Strategy 4*: Semiannual vaccine 65+ years and those with immunocompromising conditions |
| Risk-based (expanded) | *Strategy 5*: Annual vaccine 65+ years and to high risk or immunocompromised individuals 18-64 years  *Strategy 6**: Semiannual vaccine 65+ years and to high risk or immunocompromised individuals 18-64 years |
| Hybrid | *Strategy 7*: Annual vaccine 6+ months with second dose in 65+ years and those with immunocompromising conditions  *Strategy 8**: Annual vaccine 18+ years with second dose in 65+ years and those with immunocompromising conditions |

*These strategies are not included in the main text

##### Table S5: COVID-19 hospitalization risk at US national level by vaccination start month under annual universal vaccination with one-month uptake, accumulated over the 12-month study period (May 2024–April 2025).

| Vaccination  timing scenario | Age-risk group | Total hospitalizations | Absolute risk, total population  (per 100,000 persons) | Absolute risk, vaccinated  (per 100,000 vaccinated) | Absolute risk reduction ^a^  (per 100,000 vaccinated) | NNV to avert one hospitalization ^a^ | Relative risk reduction ^b^  (%) |
| --- | --- | --- | --- | --- | --- | --- | --- |
| May | 18-64 years, low risk | 244 [220, 276] | 13.1 [11.8, 14.8] | 9.6 [6.5, 14.6] | 4.5 [0.2, 8.9] | 21,956  [11,161, 158,959] | 8 [-7.6, 20.3] |
|  | 18-64 years, high risk | 931 [866, 992] | 94.5 [88, 100.7] | 68.5 [58, 78.5] | 38.1 [25.3, 50.2] | 2,628  [1,990, 3,966] | 11.9 [3.9, 18.3] |
|  | 18-64 years, immunocompromised | 537 [497, 592] | 294.9 [272.9, 324.9] | 218.9 [179.4, 264.9] | 112.8 [55.3, 153.8] | 887  [650, 1,816] | 9.7 [-0.5, 19.3] |
|  | 65+ years | 4,170 [3,933, 4,382] | 472.5 [445.7, 496.6] | 398.7 [368.3, 423.1] | 180.5 [154.9, 203.3] | 554  [492, 646] | 18.2 [14.9, 21.5] |
| June | 18-64 years, low risk | 241 [209, 264] | 12.9 [11.2, 14.2] | 8.6 [5, 12.3] | 5.8 [1.5, 9.6] | 17,277  [10,465, 67,231] | 10.5 [-2.9, 23.7] |
|  | 18-64 years, high risk | 898 [849, 959] | 91.2 [86.2, 97.4] | 63.4 [54.2, 74.3] | 43.5 [32.6, 55.2] | 2,299  [1,812, 3,065] | 14.2 [7.1, 20.4] |
|  | 18-64 years, immunocompromised | 512 [474, 561] | 281.4 [260.6, 307.8] | 202.4 [163.1, 239] | 128.2 [72.3, 172.3] | 780  [581, 1,382] | 14.1 [2.7, 21.6] |
|  | 65+ years | 3,987 [3,746, 4,181] | 451.8 [424.5, 473.8] | 366.6 [343.2, 392.8] | 210.5 [183.8, 234.8] | 475  [426, 544] | 21.8 [18.8, 24.5] |
| July | 18-64 years, low risk | 235 [209, 260] | 12.6 [11.2, 13.9] | 8.2 [4.5, 11.1] | 6.1 [1.8, 9.8] | 16,145  [10,177, 46,490] | 12.4 [-2.1, 24.9] |
|  | 18-64 years, high risk | 892 [821, 965] | 90.6 [83.4, 98] | 60.5 [49.8, 68.5] | 46.8 [33.2, 55.6] | 2,136  [1,799, 3,011] | 15.9 [7, 22.8] |
|  | 18-64 years, immunocompromised | 512 [462, 556] | 281.4 [253.7, 305.4] | 193.4 [158.7, 239.3] | 135.9 [85.1, 176.1] | 736  [568, 1,176] | 14.1 [4.3, 24.5] |
|  | 65+ years | 3,893 [3,731, 4,081] | 441.1 [422.8, 462.5] | 353 [330.9, 379.1] | 223.8 [199.5, 248.2] | 447  [403, 501] | 23.6 [20.1, 26.5] |
| August | 18-64 years, low risk | 244 [216, 270] | 13.1 [11.6, 14.5] | 9.1 [5, 12.9] | 5.4 [1.2, 9.1] | 18,479  [10,943, 64,754] | 8.4 [-5.5, 21.7] |
|  | 18-64 years, high risk | 909 [840, 969] | 92.3 [85.3, 98.3] | 63.6 [52.6, 73.8] | 43.9 [31.5, 54.9] | 2,280  [1,823, 3,179] | 13.8 [6.5, 20.9] |
|  | 18-64 years, immunocompromised | 520 [477, 560] | 285.3 [262.2, 307.6] | 202.6 [163.6, 239.1] | 127.9 [73.8, 176.4] | 782  [567, 1,355] | 13.1 [1.7, 24.1] |
|  | 65+ years | 3,994 [3,786, 4,275] | 452.5 [429, 484.5] | 372.6 [333.8, 407.3] | 206.2 [176.4, 238.5] | 485  [419, 567] | 21.5 [17.9, 25.8] |
| September | 18-64 years, low risk | 246 [211, 280] | 13.2 [11.3, 15] | 9.1 [5.5, 13] | 5.1 [1.6, 8.7] | 19,447  [11,550, 63,134] | 8.6 [-7, 20.8] |
|  | 18-64 years, high risk | 930 [855, 996] | 94.4 [86.8, 101.1] | 67.8 [57, 78.4] | 39.3 [26.8, 53.2] | 2,547  [1,879, 3,732] | 11.4 [3.8, 20.5] |
|  | 18-64 years, immunocompromised | 534 [488, 578] | 293.5 [268.2, 317.2] | 215.2 [174.5, 255.6] | 110.5 [79.9, 156.2] | 905  [641, 1,252] | 10.7 [-0.3, 19.3] |
|  | 65+ years | 4,136 [3,916, 4,470] | 468.7 [443.7, 506.5] | 394 [368.9, 438.8] | 178.8 [149.5, 207] | 559  [483, 669] | 18.5 [13.8, 22.6] |
| October | 18-64 years, low risk | 247 [222, 280] | 13.3 [11.9, 15] | 9.6 [6.5, 13.4] | 5 [0.6, 8.5] | 19,863  [11,819, 123,006] | 7.9 [-8.8, 18.9] |
|  | 18-64 years, high risk | 934 [862, 1043] | 94.8 [87.5, 105.9] | 69.3 [59, 81.4] | 37.6 [25.6, 50.3] | 2,662  [1,986, 3,905] | 11.9 [1, 18.7] |
|  | 18-64 years, immunocompromised | 537 [489, 587] | 294.9 [268.7, 322.4] | 221.2 [185.1, 269.6] | 107.5 [56.3, 150.7] | 930  [664, 1,782] | 10.7 [-1.1, 19.8] |
|  | 65+ years | 4,199 [4,000, 4,495] | 475.8 [453.3, 509.4] | 412.7 [382.9, 446.5] | 165.1 [136.4, 190.4] | 606  [525, 733] | 17.4 [13.2, 21.2] |
| November | 18-64 years, low risk | 244 [215, 280] | 13.1 [11.6, 15] | 9.6 [5.8, 14.2] | 4.8 [-0.1, 8.7] | 20,657  [11,425, 118,479] | 9.8 [-7.6, 19.5] |
|  | 18-64 years, high risk | 921 [839, 991] | 93.5 [85.2, 100.5] | 69 [59.5, 80.4] | 37.2 [25.6, 49.3] | 2,686  [2,027, 3,903] | 13.1 [4.1, 18.6] |
|  | 18-64 years, immunocompromised | 535 [474, 588] | 293.8 [260.6, 322.9] | 215.7 [174.5, 259.9] | 111.4 [63.3, 157.5] | 897  [635, 1,582] | 11.2 [0.7, 20.6] |
|  | 65+ years | 4,170 [3,958, 4,377] | 472.5 [448.5, 496] | 406.8 [379.2, 431.2] | 170.8 [143.5, 197.8] | 585  [506, 697] | 18.4 [14.4, 21.1] |

All scenarios assume annual universal vaccination at historical vaccine coverage. Values are medians with 95% uncertainty intervals across 100 simulation runs.

^a^ Absolute risk reduction and NNV compare the no-vaccination reference risk (Table S14) with risk among the vaccinated subgroup.
^b^ Relative risk reduction compares the no-vaccination reference with risk in the entire age-risk group, so it does not equal absolute risk reduction divided by the reference risk.

**Table S6: Statistical comparison of hospitalization risk at US national level** **by vaccination month under annual universal vaccination with one-month uptake.**

| Age-risk group | Population | Most favorable month (lowest median hospitalization risk) | Superior in majority of runs vs each other month ^a^ | Significant pairwise comparisons ^b^ |
| --- | --- | --- | --- | --- |
| 18-64 years, low risk | Total | July | Yes | 5/6 |
|  | Vaccinated | July | Yes | 4/6 |
| 18-64 years, high risk | Total | July | Yes | 5/6 |
|  | Vaccinated | July | Yes | 6/6 |
| 18-64 years, immunocompromised | Total | July | Yes | 4/6 |
|  | Vaccinated | July | Yes | 4/6 |
| 65+ years | Total | July | Yes | 6/6 |
|  | Vaccinated | July | Yes | 6/6 |

Corresponds to results in Table S5.

^a^ indicates whether the most favorable month had lower risk in >50% of pairwise comparisons, compared to each of the six other months.

^b^ the number of other months for which the most favorable month had significantly lower hospitalization risk by a Holm-corrected paired Wilcoxon test (p < 0.05).

##### Table S7: Model-based estimates of COVID-19 hospitalization risk at US national level under different vaccination timing with historical vaccine uptake over four months, evaluating cumulative risk over 12-month study period.

| Vaccination  timing scenario | Age-risk group | Total hospitalizations | Absolute risk, total population  (per 100,000 persons) | Absolute risk, vaccinated  (per 100,000 vaccinated) | Absolute risk reduction ^a^  (per 100,000 vaccinated) | NNV to avert one hospitalization ^a^ | Relative risk reduction ^b^  (%) |
| --- | --- | --- | --- | --- | --- | --- | --- |
| May | 18-64 years, low risk | 238  [204, 265] | 12.8 [11, 14.2] | 8.6 [5.3, 12.7] | 5.7 [1.5, 8.5] | 17,539 [11,768, 67,876] | 11.2 [-2.9, 22.5] |
|  | 18-64 years,  high risk | 906 [835, 966] | 92 [84.7, 98.1] | 63.2 [54.8, 71.9] | 44.2 [30.9, 52.7] | 2,264 [1,896, 3,233] | 14 [6.8, 20] |
|  | 18-64 years, immunocompromised | 518 [469, 569] | 284.7 [257.5, 312.6] | 203.7 [154.1, 239.1] | 124.8 [83.9, 173.2] | 801 [577, 1,192] | 13.2 [3.3, 21.9] |
|  | 65+ years | 3,994 [3,775, 4,162] | 452.6 [427.8, 471.6] | 369.5 [345.9, 396.4] | 208 [187.9, 234.6] | 481 [426, 532] | 21.6 [18.8, 24.9] |
| June | 18-64 years, low risk | 238 [205, 273] | 12.8 [11, 14.7] | 8.2 [4.8, 12.5] | 5.9 [1.9, 9.1] | 16,818 [11,045, 51,597] | 10.8 [-3.7, 23.9] |
|  | 18-64 years,  high risk | 894 [838, 948] | 90.8 [85.1, 96.2] | 62 [52.7, 74.3] | 43.8 [31.1, 57.1] | 2,284 [1,753, 3,214] | 15.4 [7.9, 21.7] |
|  | 18-64 years, immunocompromised | 516 [470, 573] | 283.6 [258.4, 314.5] | 199.6 [155.2, 234.3] | 135.5 [73, 175] | 738 [571, 1,371] | 14.4 [0.2, 22.7] |
|  | 65+ years | 3,924 [3,742, 4,138] | 444.7 [424, 468.9] | 356.4 [330.4, 380.5] | 219.9 [197.8, 245.6] | 455 [407, 505] | 22.6 [20.2, 25.9] |
| July | 18-64 years, low risk | 242 [214, 271] | 13 [11.5, 14.5] | 8.6 [4.5, 12.7] | 5.8 [2, 9.5] | 17,281 [10,547, 46,499] | 10.5 [-5.3, 22.4] |
|  | 18-64 years,  high risk | 906 [851, 973] | 92 [86.4, 98.7] | 64.7 [53.9, 72.9] | 42.5 [32.3, 53.7] | 2,351 [1,864, 3,100] | 13.3 [6.3, 21] |
|  | 18-64 years, immunocompromised | 519 [478, 587] | 285 [262.4, 322.4] | 194.6 [154.3, 246.5] | 131.6 [71.5, 182.1] | 760 [549, 1,435] | 12.4 [0.4, 22.5] |
|  | 65+ years | 3,977 [3,795, 4,197] | 450.7 [430.1, 475.5] | 366.5 [338.2, 394.3] | 213 [181.7, 231.3] | 470 [432, 550] | 21.8 [18.1, 24.9] |
| August | 18-64 years, low risk | 243 [210, 280] | 13 [11.3, 15] | 9.6 [5.8, 13.4] | 5 [1, 8.5] | 19,962 [11,726, 75,804] | 9.4 [-5.4, 20.6] |
|  | 18-64 years,  high risk | 921 [858, 985] | 93.5 [87.1, 100] | 66.8 [55.9, 78.5] | 40.8 [25.5, 52.1] | 2,451 [1,920, 3,934] | 12.5 [4, 21.1] |
|  | 18-64 years, immunocompromised | 528 [490, 571] | 290 [269.1, 313.4] | 207.4 [164.6, 247.5] | 123.2 [71.4, 165.9] | 811 [603, 1403] | 11.3 [-1.4, 21.9] |
|  | 65+ years | 4,071 [3,887, 4,335] | 461.3 [440.4, 491.2] | 383.2 [356.9, 417.1] | 192.4 [164.6, 218.9] | 520 [457, 608] | 19.7 [16, 23.3] |
| September | 18-64 years, low risk | 244 [219, 278] | 13.1 [11.8, 14.9] | 9.4 [6.2, 13] | 5.1 [0.9, 8.3] | 19,595 [12,093, 112,764] | 9.5 [-5.8, 19.4] |
|  | 18-64 years,  high risk | 932 [841, 998] | 94.7 [85.4, 101.3] | 67.5 [59.3, 78.5] | 38.2 [27.7, 48.8] | 2,621 [2,048, 3,611] | 11.8 [4.5, 20.5] |
|  | 18-64 years, immunocompromised | 536 [490, 581] | 294.1 [269.3, 318.8] | 213.6 [171.7, 254.9] | 114.6 [66.2, 159.4] | 872 [627, 1512] | 10.9 [-1.1, 20.7] |
|  | 65+ years | 4,126 [3,928, 4,405] | 467.5 [445.1, 499.1] | 397.4 [375.5, 432.6] | 177.7 [150.3, 205.8] | 563 [486, 665] | 18.7 [14.8, 22] |
| October | 18-64 years, low risk | 242 [215, 274] | 13 [11.6, 14.7] | 9.6 [6.5, 13.2] | 4.7 [0.7, 8.2] | 21,258 [12,147, 107,431] | 10.1 [-4.9, 20.9] |
|  | 18-64 years,  high risk | 922 [848, 982] | 93.6 [86.1, 99.7] | 68.1 [60.4, 81.9] | 38.6 [26.6, 48] | 2,588 [2,082, 3,763] | 12.3 [5.5, 21.1] |
|  | 18-64 years, immunocompromised | 532 [484, 575] | 292.4 [265.8, 315.8] | 214.3 [174.9, 253.2] | 111.4 [76.7, 155.1] | 898 [645, 1,304] | 11.1 [0.9, 20.6] |
|  | 65+ years | 4,142 [3,939, 4,374] | 469.3 [446.4, 495.6] | 404.7 [381.6, 431.1] | 169.8 [143.9, 199.4] | 589 [501, 695] | 18.5 [15.3, 22.4] |
| November | 18-64 years, low risk | 244 [210, 272] | 13.1 [11.3, 14.6] | 10.1 [6.7, 13.9] | 4.4 [0.2, 7.9] | 22,551 [12,693, 140,951] | 9 [-4.9, 23.4] |
|  | 18-64 years,  high risk | 930 [860, 990] | 94.5 [87.3, 100.4] | 71.8 [62.2, 83.7] | 35 [22.9, 46] | 2,857 [2,176, 4,373] | 11.7 [3.1, 19.1] |
|  | 18-64 years, immunocompromised | 535 [493, 589] | 293.8 [270.7, 323.4] | 223.3 [180.8, 272] | 104.2 [49.6, 154.3] | 960 [648, 2,017] | 10.5 [-3.9, 20.1] |
|  | 65+ years | 4,226 [4,035, 4,430] | 478.8 [457.3, 502] | 420 [398.3, 447.3] | 154.8 [125.5, 182.7] | 646 [547, 797] | 17.2 [13.8, 20.9] |

All scenarios assume annual universal vaccination at historical vaccine coverage. Values are medians with 95% uncertainty intervals across 100 simulation runs. NNV, number needed to vaccinate to prevent one hospitalization.

^a^ Absolute risk reduction and NNV compare the no-vaccination reference risk (Table S14) with risk among the vaccinated subgroup.
^b^ Relative risk reduction compares the no-vaccination reference with risk in the entire age-risk group, so it does not equal absolute risk reduction divided by the reference risk.

##### Table S8: Model-based estimates of COVID-19 hospitalization risk for Archetype 1 under different vaccination timing with one-month vaccine uptake.

| Vaccination  timing scenario | Age-risk group | Total hospitalizations | Absolute risk, total population  (per 100,000 persons) | Absolute risk, vaccinated  (per 100,000 vaccinated) | Absolute risk reduction ^a^  (per 100,000 vaccinated) | NNV to avert one hospitalization ^a^ | Relative risk reduction ^b^  (%) |
| --- | --- | --- | --- | --- | --- | --- | --- |
| Reference ^c^ | 18-64 years, low risk | 265 [233, 291] | 14.2 [12.5, 15.6] | - | - | - | - |
|  | 18-64 years, high risk | 1059 [995, 1123] | 107.5 [101, 114] | - | - | - | - |
|  | 18-64 years, immunocompromised | 598 [547, 652] | 328.7 [300.4, 358] | - | - | - | - |
|  | 65+ years | 5,089 [4,884, 5,339] | 576.7 [553.4, 604.9] | - | - | - | - |
| May | 18-64 years, low risk | 248 [217, 283] | 13.3 [11.7, 15.2] | 8.6 [4.8, 13.5] | 5.3 [1, 9.8] | 18,675  [10,155, 68,367] | 7 [-9, 20.9] |
|  | 18-64 years, high risk | 932 [841, 1,016] | 94.6 [85.4, 103.1] | 65.7 [53.3, 76.1] | 41.6 [29.7, 54.5] | 2,401  [1,837, 3,363] | 11.8 [4.9, 19.2] |
|  | 18-64 years, immunocompromised | 538 [480, 590] | 295.4 [263.8, 324] | 205.2 [165.8, 247.4] | 123.3 [70.7, 169.5] | 811  [591, 1,414] | 9.9 [-0.2, 19.2] |
|  | 65+ years | 4,117 [3,849, 4,330] | 466.5 [436.1, 490.7] | 375.9 [342.3, 403.5] | 202.4 [176.3, 226] | 494  [442, 567] | 19.3 [15.3, 22.9] |
| June | 18-64 years, low risk | 242 [209, 271] | 13 [11.2, 14.6] | 8.2 [5.3, 13] | 5.7 [1.2, 9.6] | 17,421  [10,415, 55,213] | 9.1 [-5.9, 20.7] |
|  | 18-64 years, high risk | 902 [851, 973] | 91.6 [86.4, 98.8] | 60.4 [49.8, 69.9] | 48.1 [36.8, 58.7] | 2,081  [1,705, 2,715] | 14.3 [6.5, 21.1] |
|  | 18-64 years, immunocompromised | 520 [479, 570] | 285.3 [263, 312.8] | 188.8 [155, 231.7] | 141.3 [87.6, 180.8] | 708  [553, 1,142] | 13.3 [3.5, 20.5] |
|  | 65+ years | 3,922 [3,714, 4,123] | 444.5 [420.8, 467.2] | 351.2 [319.6, 375.6] | 228.2 [203.2, 256] | 438  [391, 492] | 23 [20, 27.1] |
| July | 18-64 years, low risk | 242 [208, 272] | 13 [11.2, 14.6] | 8.1 [4.3, 12.3] | 6.1 [2, 9.8] | 16,176  [10,160, 45,102] | 9.7 [-9.7, 21.4] |
|  | 18-64 years, high risk | 907 [841, 975] | 92.1 [85.4, 99] | 60.1 [50.9, 70.9] | 46.9 [34.6, 57.3] | 2,132  [1,744, 2,903] | 14.3 [6, 20.5] |
|  | 18-64 years, immunocompromised | 518 [476, 568] | 284.2 [261.6, 311.7] | 195.6 [158.6, 229.5] | 135 [92.1, 179.9] | 741  [556, 1,086] | 13.4 [3, 21.9] |
|  | 65+ years | 3,936 [3,693, 4,136] | 445.9 [418.5, 468.7] | 356.6 [318.8, 378.5] | 224.6 [187.1, 262.9] | 445  [380, 534] | 22.9 [18.6, 27.2] |
| August | 18-64 years, low risk | 242 [217, 274] | 13 [11.7, 14.7] | 9.1 [5.8, 13.4] | 5.3 [0.2, 9.2] | 18,565  [10,863, 168,328] | 8.7 [-7, 20.5] |
|  | 18-64 years, high risk | 933 [854, 1,003] | 94.7 [86.7, 101.8] | 68 [54.6, 79.8] | 39.2 [24.9, 54.4] | 2,551  [1,839, 4,022] | 12.6 [4.5, 19.5] |
|  | 18-64 years, immunocompromised | 535 [487, 580] | 293.8 [267.7, 318.2] | 208.8 [173.4, 256.4] | 117.7 [71.7, 161.2] | 850  [621, 1,394] | 11 [0.7, 20] |
|  | 65+ years | 4,124 [3,919, 4,407] | 467.3 [444.1, 499.4] | 396.2 [357.8, 435.1] | 182.6 [141.6, 219.6] | 548  [455, 706] | 18.5 [13.3, 23.2] |
| September | 18-64 years, low risk | 250 [223, 279] | 13.5 [12, 15] | 10.5 [6.2, 14.1] | 4.1 [0.6, 8.5] | 24,528  [11,753, 143,953] | 5.7 [-10, 18.5] |
|  | 18-64 years, high risk | 951 [883, 1022] | 96.5 [89.7, 103.8] | 73.5 [63.8, 85.7] | 34.3 [21.3, 46.8] | 2,918  [2,138, 4,687] | 10.3 [4.4, 16.9] |
|  | 18-64 years, immunocompromised | 536 [487, 590] | 294.1 [267.3, 323.7] | 229.5 [190.9, 272.3] | 97.9 [53.1, 148.2] | 1,021  [675, 1,884] | 10.5 [0.5, 19.8] |
|  | 65+ years | 4,306 [4,079, 4,626] | 488 [462.2, 524.2] | 428.5 [396.7, 466.7] | 146.4 [116.9, 179] | 683  [559, 855] | 15.4 [9.7, 18.9] |
| October | 18-64 years, low risk | 248 [219, 285] | 13.3 [11.8, 15.3] | 10.1 [5.8, 14.9] | 4.1 [-1.4, 7.9] | 21,942  [12,416, 196,342] | 6.1 [-9.5, 22.7] |
|  | 18-64 years, high risk | 956 [878, 1025] | 97 [89.2, 104] | 76.3 [63.6, 90.1] | 31.9 [12.7, 43.9] | 3,135  [2,279, 7,897] | 9.8 [2.7, 17] |
|  | 18-64 years, immunocompromised | 548 [509, 602] | 300.7 [279.8, 330.6] | 233.2 [190.6, 270.3] | 97.9 [49.7, 139] | 1,022  [720, 2,025] | 9.2 [-2.9, 17] |
|  | 65+ years | 4,402 [4,109, 4,602] | 498.8 [465.7, 521.5] | 445.6 [410.8, 471.7] | 129.2 [108.2, 162.8] | 774  [614, 925] | 13.6 [10.4, 17.6] |
| November | 18-64 years, low risk | 247 [222, 283] | 13.3 [11.9, 15.2] | 10.1 [6.7, 14.1] | 3.8 [0.5, 8.2] | 25,783  [12,188, 113,358] | 7.5 [-8.7, 19.3] |
|  | 18-64 years, high risk | 954 [879, 1016] | 96.8 [89.2, 103.1] | 76.7 [65.2, 88.6] | 30.6 [19.6, 43.2] | 3,266  [2,318, 5,095] | 9.7 [3, 18.8] |
|  | 18-64 years, immunocompromised | 534 [485, 582] | 293.2 [266.6, 319.6] | 236.4 [199.7, 280.3] | 92 [38.6, 142.2] | 1,087  [704, 2,597] | 11.2 [-0.7, 19.7] |
|  | 65+ years | 4,416 [4,187, 4,590] | 500.4 [474.5, 520.1] | 453.4 [423.5, 478.7] | 120.6 [95.8, 158.2] | 829  [632, 1,044] | 13.5 [9.7, 17.1] |

Assumed historical vaccine coverage in one-month uptake. Reports median and 95% uncertainty intervals over 100 simulations.

^a^ Absolute risk reduction and NNV compare the no-vaccination reference risk (Table S14) with risk among the vaccinated subgroup.
^b^ Relative risk reduction compares the no-vaccination reference with risk in the entire age-risk group, so it does not equal absolute risk reduction divided by the reference risk.

^c^ Reference group: No additional vaccination during the study period

**Table S9: Statistical comparison of hospitalization risk by vaccination month for Archetype 1 under annual universal vaccination with one-month uptake.**

| Age-risk group | Population | Most favorable month (lowest median hospitalization risk) | Superior in majority of runs vs each other month ^a^ | Significant pairwise comparisons ^b^ |
| --- | --- | --- | --- | --- |
| 18-64 years, low risk | Total | August | No | 2/6 |
|  | Vaccinated | July | Yes | 4/6 |
| 18-64 years, high risk | Total | June | Yes | 5/6 |
|  | Vaccinated | July | No | 5/6 |
| 18-64 years, immunocompromised | Total | July | Yes | 5/6 |
|  | Vaccinated | June | Yes | 5/6 |
| 65+ years | Total | June | No | 5/6 |
|  | Vaccinated | June | Yes | 6/6 |

Corresponds to results in Table S8.

^a^ indicates whether the most favorable month had lower risk in >50% of pairwise comparisons, compared to each of the six other months.

^b^ the number of other months for which the most favorable month had significantly lower hospitalization risk, by a Holm-corrected paired Wilcoxon test (p < 0.05).

##### Table S10: Model-based estimates of COVID-19 hospitalization risk for Archetype 2 under different vaccination timing with one-month vaccine uptake.

| Vaccination  timing scenario | Age-risk group | Total hospitalizations | Absolute risk, total population  (per 100,000 persons) | Absolute risk, vaccinated  (per 100,000 vaccinated) | Absolute risk reduction ^a^  (per 100,000 vaccinated) | NNV to avert one hospitalization ^a^ | Relative risk reduction ^b^  (%) |
| --- | --- | --- | --- | --- | --- | --- | --- |
| Reference ^c^ | 18-64 years, low risk | 342 [308, 379] | 18.4 [16.5, 20.3] | - | - | - | - |
|  | 18-64 years, high risk | 1366 [1278, 1461] | 138.7 [129.7, 148.3] | - | - | - | - |
|  | 18-64 years, immunocompromised | 781 [718, 830] | 428.9 [394.1, 455.9] | - | - | - | - |
|  | 65+ years | 6,636 [6,350, 6,878] | 751.9 [719.6, 779.3] | - | - | - | - |
| May | 18-64 years, low risk | 326 [293, 374] | 17.5 [15.7, 20.1] | 12.9 [9.1, 18.2] | 5.3 [0.1, 9.8] | 18,753  [10,154, 120,338] | 3.7 [-11.6, 15.5] |
|  | 18-64 years, high risk | 1276 [1186, 1375] | 129.6 [120.4, 139.5] | 99.7 [86.2, 111.1] | 39.5 [27.7, 56.8] | 2,532  [1,761, 3,622] | 6.2 [-2.6, 16.1] |
|  | 18-64 years, immunocompromised | 734 [676, 804] | 402.8 [371.2, 441.5] | 313.2 [263.5, 363.1] | 119 [63.6, 163.7] | 840  [611, 1,574] | 6.2 [-4.3, 13.8] |
|  | 65+ years | 5,733 [5,494, 6,114] | 649.6 [622.6, 692.8] | 571 [537.9, 616.2] | 180.1 [134.5, 212.8] | 555  [470, 744] | 13.1 [8.3, 16.6] |
| June | 18-64 years, low risk | 319 [287, 365] | 17.1 [15.4, 19.6] | 12.5 [7.9, 17.8] | 5.9 [0.3, 11.2] | 16,485  [8,877, 57,385] | 5.8 [-10.8, 19.5] |
|  | 18-64 years, high risk | 1232 [1161, 1353] | 125 [117.9, 137.3] | 92.1 [80.1, 106.6] | 47.1 [31, 60.4] | 2,122  [1,655, 3,223] | 9.8 [-0.4, 17.6] |
|  | 18-64 years, immunocompromised | 711 [649, 784] | 390.4 [356.3, 430.3] | 288.5 [246.5, 341.4] | 139.1 [89.9, 183.2] | 719  [546, 1,113] | 9.1 [1, 17.5] |
|  | 65+ years | 5,563 [5,301, 5,888] | 630.4 [600.7, 667.2] | 529.9 [497, 576.2] | 218 [189.1, 252.3] | 459  [396, 529] | 16.5 [12, 20] |
| July | 18-64 years, low risk | 314 [273, 343] | 16.9 [14.7, 18.4] | 12 [7.9, 16.8] | 6.6 [1.3, 10.4] | 15,108  [9,604, 41,160] | 7.7 [-4.7, 21] |
|  | 18-64 years, high risk | 1,188 [1,096, 1,289] | 120.6 [111.2, 130.8] | 85.4 [75.9, 95.9] | 53.3 [40, 71.9] | 1,878  [1,391, 2,504] | 13.3 [4.4, 21.2] |
|  | 18-64 years, immunocompromised | 675 [625, 741] | 370.7 [343.5, 407] | 262.4 [223, 314.4] | 162.6 [115.9, 205.5] | 615  [487, 863] | 13.3 [4.5, 20.6] |
|  | 65+ years | 5,322 [5,031, 5,522] | 603.1 [570.1, 625.8] | 498.2 [466.6, 524.5] | 256.1 [223, 283.7] | 390  [352, 449] | 20.3 [15.8, 23.3] |
| August | 18-64 years, low risk | 308 [278, 343] | 16.6 [14.9, 18.4] | 11.5 [7.4, 16.8] | 6.8 [1.4, 11.1] | 14,614  [9,028, 73,130] | 10.2 [-4.5, 18.9] |
|  | 18-64 years, high risk | 1,191 [1,096, 1,262] | 120.9 [111.2, 128.1] | 84.2 [72.3, 93.9] | 55.1 [41.2, 67.8] | 1,816  [1,475, 2,425] | 13.3 [5.4, 19.3] |
|  | 18-64 years, immunocompromised | 678 [623, 749] | 372.3 [342.4, 411.1] | 262.9 [218.7, 313.7] | 165.3 [110, 209.6] | 605  [477, 909] | 13.9 [2.6, 21.7] |
|  | 65+ years | 5,286 [5,055, 5,479] | 599 [572.8, 620.9] | 491.7 [462.3, 525.3] | 256.1 [225.3, 302.1] | 391  [331, 444] | 20.3 [17.2, 24.4] |
| September | 18-64 years, low risk | 312 [286, 349] | 16.8 [15.4, 18.7] | 12 [8.1, 16.8] | 6.8 [0.8, 10] | 14,760  [9,972, 40,634] | 8.3 [-3.9, 17.9] |
|  | 18-64 years, high risk | 1210 [1137, 1286] | 122.9 [115.4, 130.5] | 87.2 [75.8, 99.2] | 51.1 [35.9, 66.3] | 1,959  [1,508, 2,789] | 11.9 [4.1, 18.4] |
|  | 18-64 years, immunocompromised | 694 [649, 735] | 380.8 [356.6, 403.6] | 277.1 [231.3, 328] | 148.5 [101.2, 204.4] | 674  [489, 988] | 11.5 [2.9, 18.4] |
|  | 65+ years | 5,439 [5,144, 5,633] | 616.3 [582.9, 638.3] | 515.5 [475.8, 545] | 235.9 [198.6, 279.2] | 424  [358, 503] | 18.2 [14.6, 22.3] |
| October | 18-64 years, low risk | 320 [289, 355] | 17.2 [15.5, 19] | 12 [7.9, 17.7] | 6.2 [0.4, 10.3] | 15,914  [9,752, 153,476] | 6.2 [-6.8, 17.1] |
|  | 18-64 years, high risk | 1,227 [1,135, 1,311] | 124.6 [115.2, 133] | 89.7 [78.1, 104.6] | 48.4 [32.5, 61.7] | 2,065  [1,620, 3,075] | 10.1 [3.3, 18.2] |
|  | 18-64 years, immunocompromised | 704 [650, 762] | 386.9 [357.1, 418.5] | 281.8 [239.6, 336.9] | 146.7 [93.2, 193.4] | 681  [517, 1,085] | 10.2 [-1.5, 18] |
|  | 65+ years | 5,541 [5,207, 5,762] | 627.9 [590, 653] | 536.5 [499.3, 572.7] | 214.4 [177.9, 246.3] | 466  [406, 562] | 16.9 [13.4, 20.3] |
| November | 18-64 years, low risk | 316 [287, 353] | 17 [15.4, 19] | 12.7 [7.4, 17.8] | 5.8 [0.4, 10.9] | 16,778  [9,171, 77,527] | 7.1 [-4, 19.1] |
|  | 18-64 years, high risk | 1,218 [1,128, 1,279] | 123.7 [114.5, 129.8] | 89.7 [78, 103] | 49 [33.4, 62.1] | 2,042  [1,609, 3,002] | 11 [4.6, 18.2] |
|  | 18-64 years, immunocompromised | 700 [650, 761] | 384.7 [356.9, 417.9] | 285.6 [234.7, 352.7] | 144.4 [87.6, 194.5] | 693  [514, 1,142] | 10.1 [1.9, 17.2] |
|  | 65+ years | 5,468 [5,240, 5,792] | 619.5 [593.8, 656.3] | 530.7 [494.6, 579.9] | 221.1 [171.8, 257.6] | 452  [388, 582] | 17.4 [12.8, 20.9] |

Assumed historical vaccine coverage in one-month uptake. Reports median and 95% uncertainty intervals over 100 simulations.

^a^ Absolute risk reduction and NNV compare the no-vaccination reference risk (Table S14) with risk among the vaccinated subgroup.
^b^ Relative risk reduction compares the no-vaccination reference with risk in the entire age-risk group, so it does not equal absolute risk reduction divided by the reference risk.

^c^ Reference group: No additional vaccination during the study period

**Table S11: Statistical comparison of hospitalization risk by vaccination month for Archetype 2 under annual universal vaccination with one-month uptake.**

| Age-risk group | Population | Most favorable month (lowest median hospitalization risk) | Superior in majority of runs vs each other month ^a^ | Significant pairwise comparisons ^b^ |
| --- | --- | --- | --- | --- |
| 18-64 years, low risk | Total | August | Yes | 4/6 |
|  | Vaccinated | August | No | 2/6 |
| 18-64 years, high risk | Total | July | No | 5/6 |
|  | Vaccinated | August | Yes | 5/6 |
| 18-64 years, immunocompromised | Total | July | Yes | 5/6 |
|  | Vaccinated | July | No | 5/6 |
| 65+ years | Total | August | No | 5/6 |
|  | Vaccinated | August | Yes | 5/6 |

Corresponds to results in Table S10.

^a^ indicates whether the most favorable month had lower risk in >50% of pairwise comparisons, compared to each of the six other months.

^b^ the number of other months for which the most favorable month had significantly lower hospitalization risk, by a Holm-corrected paired Wilcoxon test (p < 0.05).

**Table S12: Statistical comparison of hospitalization risk at the US national level with reduced summer vaccination coverage.**

| Coverage scenario | Age-risk group | Population | Most favorable month (lowest median hospitalization risk) | Superior in majority of runs vs each other month ^a^ | Significant pairwise comparisons ^b^ |
| --- | --- | --- | --- | --- | --- |
| 5 absolute percentage point decrease | 18-64 years, low risk | Total | November | No | 0/6* |
|  |  | Vaccinated | July | No | 0/6* |
|  | 18-64 years, high risk | Total | November | Yes | 3/6 |
|  |  | Vaccinated | July | Yes | 5/6 |
|  | 18-64 years,  immunocompromised | Total | July | No | 0/6* |
|  |  | Vaccinated | July | Yes | 5/6 |
|  | 65+ years | Total | July | Yes | 6/6 |
|  |  | Vaccinated | July | Yes | 6/6 |
| 10 absolute percentage point decrease | 18-64 years, low risk | Total | November | No | 3/6 |
|  |  | Vaccinated | September | Yes | 1/6** |
|  | 18-64 years, high risk | Total | November | Yes | 6/6 |
|  |  | Vaccinated | July | Yes | 6/6 |
|  | 18-64 years,  immunocompromised | Total | September | No | 3/6 |
|  |  | Vaccinated | July | Yes | 1/6** |
|  | 65+ years | Total | September | No | 4/6 |
|  |  | Vaccinated | July | Yes | 6/6 |

##### All scenarios use annual universal vaccination with one-month uptake, evaluating risk accumulated over the 12-month study period (May 2024–April 2025). Coverage was reduced by 5 or 10 absolute percentage points for vaccination occurring in June–August only; all other months retain historical coverage.

^a^ indicates whether the most favorable month had lower risk in >50% of pairwise comparisons, compared to each of the six other months.

^b^ the number of other months for which the most favorable month had significantly lower hospitalization risk, by a Holm-corrected paired Wilcoxon test (p < 0.05).

***** a value of 0/6 indicates that the most favorable month did not have significantly lower hospitalization risk than any other month.

****** the majority criterion and significance counts can diverge, since a month may be more favorable than another in slightly more than half of paired simulations without the difference being statistically significant.

##### Table S13: Model-based estimates of COVID-19 hospitalization risk at US national level under different vaccination timing with one-month uptake, evaluating cumulative risk over the 180-days post start of vaccination.

| Vaccination  timing scenario | Age-risk group | Total hospitalizations | Absolute risk,  total population  (per 100,000 persons) | Absolute risk, vaccinated  (per 100,000 vaccinated) | Absolute risk reduction ^a^  (per 100,000 vaccinated) | NNV to avert one hospitalization ^a^ | Relative risk reduction ^b^  (%) |
| --- | --- | --- | --- | --- | --- | --- | --- |
| May | 18-64 years, low risk | 122 [98, 143] | 6.6 [5.3, 7.7] | 4.1 [1.9, 6.7] | 3.2 [0.4, 5.6] | 31,103  [17,857, 209,356] | 10.7 [-11.2, 29.3] |
|  | 18-64 years, high risk | 442 [397, 490] | 44.9 [40.3, 49.8] | 26.9 [20.7, 33.6] | 26.4 [19.2, 33.5] | 3,787  [2,981, 5,215] | 15.6 [5.5, 26.3] |
|  | 18-64 years, immunocompromised | 252 [224, 288] | 138.4 [123.2, 158.2] | 88.8 [60.7, 111.1] | 73.5 [39.2, 100.5] | 1,360  [995, 2,557] | 14.2 [0.2, 25.2] |
|  | 65+ years | 1,862 [1,709, 2,000] | 211 [193.7, 226.6] | 155 [137.1, 170.1] | 133.6 [116, 147] | 748  [680, 862] | 26.6 [22.2, 31] |
| June | 18-64 years, low risk | 128 [103, 147] | 6.9 [5.6, 7.9] | 3.6 [1.7, 6.3] | 4 [0.9, 6.7] | 24,928  [14,835, 71,253] | 9.6 [-9, 28] |
|  | 18-64 years, high risk | 457 [388, 544] | 46.4 [39.4, 55.2] | 27 [19.1, 36] | 28.3 [19.9, 36.5] | 3,533  [2,741, 5,032] | 15.8 [4.6, 24.8] |
|  | 18-64 years, immunocompromised | 260 [221, 319] | 142.8 [121.3, 175.3] | 88.8 [61.9, 122.2] | 79.5 [54.7, 111.4] | 1,258  [898, 1,829] | 14.9 [0.9, 26.8] |
|  | 65+ years | 1,932 [1,670, 2,223] | 219 [189.2, 251.9] | 157.1 [131.6, 185.2] | 143.6 [126.6, 161.8] | 697  [618, 790] | 27 [22.6, 32.3] |
| July | 18-64 years, low risk | 142 [116, 168] | 7.6 [6.3, 9] | 4.3 [2.1, 7.2] | 4.3 [0.5, 6.7] | 23,403  [14,839, 141,487] | 8.5 [-15.9, 23.5] |
|  | 18-64 years, high risk | 530 [443, 606] | 53.8 [45, 61.5] | 31 [24.1, 38.7] | 30.9 [23.5, 38.2] | 3,234  [2,620, 4,255] | 14.4 [5.2, 24.4] |
|  | 18-64 years, immunocompromised | 302 [253, 359] | 165.8 [139, 197.2] | 101.9 [66.1, 140] | 89.9 [41.4, 124] | 1,113  [807, 2,420] | 13.4 [-2.7, 26] |
|  | 65+ years | 2,196 [1,849, 2,567] | 248.9 [209.5, 290.8] | 180.7 [148.3, 213.2] | 156.6 [133.9, 181.9] | 638  [550, 747] | 25.9 [20.5, 31.6] |
| August | 18-64 years, low risk | 152 [122, 182] | 8.2 [6.5, 9.8] | 4.8 [1.9, 7.9] | 4.5 [1.3, 6.9] | 22,084  [14,453, 79,733] | 10.8 [-8.8, 25.9] |
|  | 18-64 years, high risk | 567 [467, 652] | 57.6 [47.4, 66.1] | 35.6 [26.4, 44.9] | 34.6 [24, 44.8] | 2,887  [2,231, 4,170] | 18.3 [8.1, 26.5] |
|  | 18-64 years, immunocompromised | 324 [277, 389] | 177.9 [152.4, 213.4] | 115 [85.6, 147.7] | 97.3 [56.2, 134.2] | 1,028  [745, 1,779] | 17.4 [4, 28.4] |
|  | 65+ years | 2,402 [2,059, 2,763] | 272.2 [233.3, 313.1] | 202.2 [170.9, 236.4] | 174.4 [153.9, 195.5] | 573  [512, 650] | 28.7 [24.4, 33.4] |
| September | 18-64 years, low risk | 132 [108, 163] | 7.1 [5.8, 8.8] | 4.3 [1.9, 7] | 4.2 [1.7, 7] | 23,673  [14,228, 59,440] | 17.1 [-7.7, 34.5] |
|  | 18-64 years, high risk | 504 [450, 563] | 51.2 [45.7, 57.2] | 32.1 [23.9, 41.1] | 32.8 [23.1, 43.4] | 3,052  [2,303, 4,322] | 20.7 [11, 30.1] |
|  | 18-64 years, immunocompromised | 292 [251, 332] | 160.1 [137.8, 182.4] | 104 [78.2, 138.7] | 94.4 [66.1, 120.9] | 1,059  [827, 1,513] | 19.4 [7.6, 28.8] |
|  | 65+ years | 2,114 [1,886, 2,370] | 239.5 [213.8, 268.5] | 185.8 [163.3, 210.4] | 167.9 [147.1, 193.7] | 596  [516, 680] | 31.7 [27.9, 35.5] |
| October | 18-64 years, low risk | 118 [96, 142] | 6.3 [5.2, 7.6] | 3.8 [1.9, 5.8] | 3.8 [1, 6.6] | 26,265  [15,097, 96,416] | 17 [-6, 32.4] |
|  | 18-64 years, high risk | 437 [385, 484] | 44.4 [39.1, 49.1] | 27.3 [21.7, 33.4] | 30.5 [21.6, 37.5] | 3,281  [2,671, 4,629] | 23.2 [10.4, 32.5] |
|  | 18-64 years, immunocompromised | 254 [221, 293] | 139.8 [121.3, 161] | 89.3 [58.4, 122.9] | 87.2 [52.7, 122.6] | 1,147  [816, 1,896] | 21.8 [6.7, 34.6] |
|  | 65+ years | 1,824 [1,666, 1,949] | 206.6 [188.8, 220.8] | 155.2 [139.7, 171.3] | 155.7 [139.6, 175.1] | 642  [571, 716] | 33.5 [29.4, 37.8] |
| November | 18-64 years, low risk | 108 [88, 130] | 5.8 [4.8, 7] | 3.1 [1, 5.8] | 3.9 [0.9, 6.2] | 25,758  [16,148, 84,066] | 17.3 [-5.8, 31.7] |
|  | 18-64 years, high risk | 392 [345, 434] | 39.8 [35.1, 44.1] | 23.4 [17.1, 29.3] | 29.6 [20.8, 36.6] | 3,375  [2,731, 4,817] | 24.4 [14.4, 32.7] |
|  | 18-64 years, immunocompromised | 233 [194, 262] | 128 [106.5, 143.9] | 75.9 [51, 105.5] | 88.6 [53.3, 116.3] | 1,129  [860, 1,881] | 22.9 [8.5, 35.3] |
|  | 65+ years | 1,580 [1,462, 1,733] | 179.1 [165.6, 196.4] | 128.9 [114.2, 143] | 157.6 [133.5, 178.1] | 635  [562, 749] | 37.3 [32.5, 41.4] |

##### Assumed historical vaccine coverage. Reports median and 95% uncertainty intervals over 100 simulations. Scenarios have different reference groups for risk reduction calculations (i.e., for May and June, the reference group is hospitalization incidence under no additional vaccination from May-October and June-November, respectively).

^a^ Absolute risk reduction and NNV compare risk among the vaccinated subgroup.
^b^ Relative risk reduction compares in the entire age-risk group, so it does not equal absolute risk reduction divided by the reference risk.

##### Table S14: Model-based estimates of COVID-19 hospitalization risk at the US national level under different vaccination eligibility and dose frequency strategies with historical vaccine coverage and uptake timing, evaluating cumulative risk over 12 months.

|  | Age-risk group | Total hospitalizations | Absolute risk,  total population  (per 100,000 persons) | Absolute risk, vaccinated  (per 100,000 vaccinated) | Absolute risk reduction ^a^  (per 100,000 vaccinated) | NNV to avert one hospitalization ^a^ | Relative risk reduction ^b^  (%) |
| --- | --- | --- | --- | --- | --- | --- | --- |
| **Reference** | *Strategy 0:* No additional vaccination during the study period | | |  |  |  |  |
|  | 18-64 years, low risk | 267  [237, 296] | 14.3  [12.8, 15.9] | - | - | - | - |
|  | 18-64 years, high risk | 1,056  [962, 1,125] | 107.1  [97.7, 114.2] | - | - | - | - |
|  | 18-64 years, immunocompromised | 598  [538, 643] | 328.4  [295.7, 352.9] | - | - | - | - |
|  | 65+ years | 5,100  [4,879, 5,304] | 577.9  [552.8, 601] | - | - | - | - |
| **Universal** | *Strategy 1:* Annual vaccine 6+ months | | | | | |  |
|  | 18-64 years, low risk | 246  [215, 274] | 13.2 [11.5, 14.7] | 9.6 [5.8, 14.1] | 4.8 [1.1, 8.8] | 20,591  [11,369, 47,284] | 8 [-3.7, 23.3] |
|  | 18-64 years, high risk | 925 [842, 991] | 93.9 [85.5, 100.6] | 66.9 [57.9, 76.5] | 40.3 [29.1, 49] | 2,483  [2,039, 34,41] | 13 [3.6, 21.2] |
|  | 18-64 years, immunocompromised | 531 [483, 582] | 291.6 [265.5, 319.8] | 210.3 [164.4, 258.8] | 118.7 [53.8, 164.1] | 842  [610, 1,862] | 11.1 [0.2, 20.5] |
|  | 65+ years | 4,122 [3,952, 4,372] | 467 [447.9, 495.4] | 393.6 [371.3, 423.2] | 180.8 [164, 205.4] | 553  [487, 610] | 19.3 [15.6, 22] |
|  | *Strategy 2:* Annual vaccine to 18+ years | | |  |  |  |  |
|  | 18-64 years, low risk | 246 [218, 280] | 13.2 [11.7, 15] | 8.9 [5.8, 13.9] | 5.3 [0.8, 9.6] | 18,689  [10,429, 101,291] | 8.8 [-9.4, 19.9] |
|  | 18-64 years, high risk | 929 [859, 991] | 94.3 [87.2, 100.6] | 67.5 [57.8, 77.6] | 38.7 [25.4, 50.9] | 2,583  [1,965, 3,949] | 11.9 [3.3, 19.3] |
|  | 18-64 years, immunocompromised | 536 [493, 586] | 294.6 [270.7, 321.8] | 212.4 [170.4, 254.7] | 113 [70.6, 164.2] | 885  [609, 1,416] | 10.3 [-0.6, 19.4] |
|  | 65+ years | 4152 [3,996, 4,377] | 470.5 [452.8, 496] | 398.8 [372.8, 424.7] | 178.2 [156.7, 204.1] | 561  [490, 638] | 18.2 [15.4, 21] |
| **Risk-based (traditional)** | *Strategy 3:* Annual vaccine 65+ years and those with immunocompromising conditions | | | | |  |  |
|  | 18-64 years, low risk | 258 [230, 289] | 13.9 [12.4, 15.5] | - | - | - | 4.2 [-11.9, 15.9] |
|  | 18-64 years, high risk | 1,031 [951, 1,092] | 104.7 [96.5, 110.8] | - | - | - | 2.4 [-5.5, 9.3] |
|  | 18-64 years, immunocompromised | 556 [502, 591] | 305.3 [275.9, 324.6] | 218.5 [182.5, 267.9] | 107.3 [48, 147.1] | 932  [680, 2,149] | 8 [-4.8, 18.4] |
|  | 65+ years | 4,256 [4,064, 4,423] | 482.2 [460.6, 501.2] | 406.3 [380.3, 429.6] | 171.2 [148.2, 195.2] | 584  [512, 675] | 16.5 [13.8, 19.6] |
|  | *Strategy 4:* Semiannual vaccine 65+ years and those with immunocompromising conditions | | | | | |  |
|  | 18-64 years, low risk | 258 [232, 291] | 13.9 [12.5, 15.6] | - | - | - | 2.4 [-13.7, 16.6] |
|  | 18-64 years, high risk | 1,024 [940, 1097] | 103.9 [95.4, 111.4] | - | - | - | 2.8 [-7.5, 9.5] |
|  | 18-64 years, immunocompromised | 554 [503, 607] | 304 [276.2, 333.4] | 166.8 [90.5, 281.7] | 157.9 [48, 239.5] | 633  [417, 1,733] | 7.2 [-3.5, 19.1] |
|  | 65+ years | 4,209 [4,008, 4,401] | 476.9 [454.1, 498.7] | 307.2 [252.2, 357.5] | 270.3 [216.4, 329.6] | 370  [303, 462] | 17.4 [13.9, 20.4] |
| **Risk-based (expanded)** | *Strategy 5:* Annual vaccine 65+ years and “high risk” or those with immunocompromising conditions | | | | | |  |
|  | 18-64 years, low risk | 255 [224, 299] | 13.7 [12, 16.1] | - | - | - | 3.9 [-14.1, 19] |
|  | 18-64 years, high risk | 950 [888, 1009] | 96.5 [90.2, 102.5] | 69.2 [58.9, 79.9] | 38 [25.9, 47.9] | 2,630  [2,087, 3,859] | 10 [1.7, 17.7] |
|  | 18-64 years, immunocompromised | 540 [505, 601] | 296.3 [277.3, 329.8] | 217 [173.7, 261.9] | 110.7 [67.1, 159.3] | 903  [628, 1,491] | 9.5 [-1.9, 19] |
|  | 65+ years | 4,208 [4,035, 4,393] | 476.8 [457.3, 497.8] | 401.7 [378.6, 426.3] | 174.9 [155.4, 197.8] | 572  [505, 644] | 17.3 [14.7, 21] |
|  | *Strategy 6:* Semiannual vaccine 65+ years and “high risk” or those with immunocompromising conditions | | | | | |  |
|  | 18-64 years, low risk | 257 [221, 288] | 13.8 [11.9, 15.5] | - | - | - | 4.8 [-9.5, 16.7] |
|  | 18-64 years, high risk | 934 [863, 995] | 94.8 [87.7, 101] | 52.1 [27.7, 73.5] | 55.8 [35.5, 79.5] | 1,793  [1,257, 2,820] | 11.8 [5.4, 18.1] |
|  | 18-64 years, immunocompromised | 538 [484, 587] | 295.4 [266, 322.1] | 181.8 [99, 281.8] | 154.4 [52.2, 239.7] | 648  [417, 1,923] | 10.1 [-1.3, 19.5] |
|  | 65+ years | 4,148 [3,965, 4,338] | 470 [449.3, 491.6] | 294.9 [249.2, 345.9] | 284.9 [231.7, 328.9] | 351  [304, 432] | 18.5 [15.7, 21.5] |
| **Hybrid** | *Strategy 7:* Annual vaccine 6+ months with second dose in 65+ years and those with immunocompromising conditions ^c^ | | | | | |  |
|  | 18-64 years, low risk | 247 [215, 281] | 13.3 [11.5, 15.1] | 9.1 [5.8, 13.2] | 5.1 [0.7, 8.7] | 19,724  [11,500, 94,780] | 7.1 [-6, 22.4] |
|  | 18-64 years, high risk | 921 [859, 992] | 93.5 [87.2, 100.7] | 67.5 [58.1, 77.3] | 39.3 [27.6, 51.3] | 2,544  [1,950, 3,633] | 12.5 [5, 19.4] |
|  | 18-64 years, immunocompromised | 528 [481, 571] | 289.7 [264.4, 313.6] | 168.7 [90.9, 281.5] | 157.4 [37.9, 246.5] | 635  [406, 2,678] | 11.5 [0.3, 21.7] |
|  | 65+ years | 4,056 [3,886, 4,296] | 459.6 [440.3, 486.8] | 288.2 [255.5, 347.5] | 287.8 [228.8, 320.2] | 347  [312, 437] | 20 [16.9, 23.6] |
|  | *Strategy 8:* Annual vaccine 18+ years with second dose in 65+ years and those with immunocompromising conditions | | | | | |  |
|  | 18-64 years, low risk | 247 [214, 282] | 13.3 [11.5, 15.1] | 9.6 [5.5, 13] | 4.8 [0.8, 9.2] | 20,849  [10,881, 72,949] | 7.6 [-8.1, 19.2] |
|  | 18-64 years, high risk | 927 [879, 993] | 94.1 [89.3, 100.8] | 69.5 [59.4, 79] | 37.8 [25.2, 50.5] | 2,647  [1,980, 3,973] | 11.8 [3.7, 18.3] |
|  | 18-64 years, immunocompromised | 530 [488, 569] | 290.8 [267.9, 312.5] | 181.2 [82.9, 276.1] | 151.5 [52.2, 257.5] | 660  [388, 1,944] | 11.5 [1.7, 21.8] |
|  | 65+ years | 4,103 [3,940, 4,325] | 464.9 [446.5, 490.1] | 298.7 [241.8, 350.5] | 278.5 [224, 336.2] | 359  [297, 446] | 18.9 [16.9, 22.1] |

Reports median and 95% uncertainty intervals over 100 simulations.

^a^ Absolute risk reduction and NNV compare the no-vaccination reference risk (Table S14) with risk among the vaccinated subgroup.
^b^ Relative risk reduction compares the no-vaccination reference with risk in the entire age-risk group, so it does not equal absolute risk reduction divided by the reference risk.

^c^ Hybrid vaccination (Strategy 7) corresponds to US COVID-19 vaccine policy during the study period.

**Table S15: Statistical comparison of hospitalization risk under different vaccination strategies by eligibility and dose frequency, with historical vaccine coverage and timing.**

| Age-risk group | Population | Most favorable month (lowest median hospitalization risk) | Superior in majority of runs vs each other month ^a^ | Significant pairwise comparisons ^b^ |
| --- | --- | --- | --- | --- |
| 18-64 years, low risk | Total | Strategy 1 (Universal) | No | 4/7 |
|  | Vaccinated | Strategy 2 (Universal) | No | 0/7 |
| 18-64 years, high risk | Total | Strategy 7 (Hybrid) | No | 3/7 |
|  | Vaccinated | Strategy 6 (Risk-based, expanded semiannual) | No | 5/7 |
| 18-64 years,  immunocompromised | Total | Strategy 7 (Hybrid) | Yes | 3/7 |
|  | Vaccinated | Strategy 4 (Risk-based, semiannual) | No | 4/7 |
| 65+ years | Total | Strategy 7 (Hybrid) | Yes | 7/7 |
|  | Vaccinated | Strategy 7 (Hybrid) | Yes | 4/7 |

Corresponds to results in Table S14.

^a^ indicates whether the most favorable month had lower risk in >50% of pairwise comparisons, compared to each of the six other months.

^b^ the number of other months for which the most favorable month had significantly lower hospitalization risk, by a Holm-corrected paired Wilcoxon test (p < 0.05).

##### Table S16: Model-based estimates of COVID-19 hospitalization risk at the US national level under different vaccination eligibility and dose frequency strategies under the higher vaccine coverage scenario, evaluating cumulative risk over 12 months.

|  | Age-risk group | Total hospitalizations | Absolute risk,  total population  (per 100,000 persons) | Absolute risk, vaccinated  (per 100,000 vaccinated) | Absolute risk reduction ^a^  (per 100,000 vaccinated) | NNV to avert one hospitalization ^a^ | Relative risk reduction ^b^  (%) |
| --- | --- | --- | --- | --- | --- | --- | --- |
| **Reference** | *Strategy 0:* No additional vaccination during the study period | | |  |  |  |  |
|  | 18-64 years, low risk | 267  [237, 296] | 14.3  [12.8, 15.9] | - | - | - | - |
|  | 18-64 years, high risk | 1,056  [962, 1,125] | 107.1  [97.7, 114.2] | - | - | - | - |
|  | 18-64 years, immunocompromised | 598  [538, 643] | 328.4  [295.7, 352.9] | - | - | - | - |
|  | 65+ years | 5,100  [4,879, 5,304] | 577.9  [552.8, 601] | - | - | - | - |
| **Universal** | *Strategy 1:* Annual vaccine 6+ months | | | | | |  |
|  | 18-64 years, low risk | 182 [148, 214] | 9.8 [8, 11.5] | 7.8 [5.9, 9.5] | 6.5 [4.5, 8.7] | 15,374 [11,536, 22,487] | 32.3 [17.5, 44.4] |
|  | 18-64 years, high risk | 637 [570, 727] | 64.7 [57.8, 73.8] | 58.4 [51.8, 65.9] | 48.6 [38.7, 58.1] | 2,058 [1,722, 2,583] | 39.5 [31.2, 46.5] |
|  | 18-64 years, immunocompromised | 366 [326, 421] | 201.3 [179, 230.9] | 186.6 [165.1, 215.5] | 143.6 [104.8, 177] | 696 [565, 954] | 38.8 [29.2, 47] |
|  | 65+ years | 2,978 [2,712, 3,366] | 337.5 [307.3, 381.4] | 327 [296.5, 374.3] | 246.5 [214.6, 277.7] | 406 [360, 466] | 41 [34.8, 45.9] |
|  | *Strategy 2:* Annual vaccine to 18+ years | |  |  |  |  |  |
|  | 18-64 years, low risk | 200 [170, 233] | 10.8 [9.1, 12.5] | 8.9 [7, 10.5] | 5.4 [3.4, 8.3] | 18,590 [12,120, 29,872] | 25.1 [6.8, 38.9] |
|  | 18-64 years, high risk | 700 [635, 778] | 71 [64.5, 79] | 64 [58, 72.1] | 43 [32.8, 50.4] | 2,324 [1,985, 3,047] | 33.6 [25.7, 40.1] |
|  | 18-64 years, immunocompromised | 404 [354, 458] | 221.6 [194.6, 251.6] | 204.8 [178.1, 231.8] | 121.7 [85.9, 161.9] | 821 [618, 1165] | 31.9 [24.2, 41.5] |
|  | 65+ years | 3,285 [3,111, 3,643] | 372.2 [352.6, 412.8] | 357.5 [339.7, 403.3] | 217.2 [193.9, 240.6] | 460 [416, 516] | 35.4 [30.5, 38.6] |
| **Risk-based (traditional)** | *Strategy 3:* Annual vaccine 65+ years and those with immunocompromising conditions | | | | |  |  |
|  | 18-64 years, low risk | 256 [222, 285] | 13.7 [11.9, 15.3] | - | - | - | 5.7 [-7.8, 17.5] |
|  | 18-64 years, high risk | 1,002 [922, 1,060] | 101.8 [93.6, 107.6] | - | - | - | 4.7 [-3.5, 13.3] |
|  | 18-64 years, immunocompromised | 452 [399, 494] | 248.2 [219.1, 271.1] | 227.7 [199.1, 248.6] | 103.7 [62.1, 137.9] | 964 [725, 1,611] | 24.6 [14.3, 33.6] |
|  | 65+ years | 3,627 [3,457, 3,818] | 411 [391.8, 432.6] | 393 [373.8, 414.2] | 182.3 [164.9, 206.9] | 549 [483, 606] | 28.6 [25.7, 31.7] |
|  | *Strategy 4:* Semiannual vaccine 65+ years and those with immunocompromising conditions | | | | |  |  |
|  | 18-64 years, low risk | 249 [217, 282] | 13.4 [11.6, 15.1] | - | - | - | 6.9 [-7.9, 18.5] |
|  | 18-64 years, high risk | 972 [908, 1,040] | 98.6 [92.2, 105.5] | - | - | - | 8 [-1.3, 16.1] |
|  | 18-64 years, immunocompromised | 360 [329, 398] | 198 [180.9, 218.3] | 162.2 [139.7, 185.4] | 165.5 [128.7, 197.4] | 604 [507, 777] | 39.4 [30.2, 45.6] |
|  | 65+ years | 2,506 [2,361, 2,668] | 284 [267.6, 302.3] | 257.9 [240.8, 275.7] | 319.2 [300.4, 339] | 313 [295, 333] | 50.7 [48.7, 53.1] |
| **Risk-based (expanded)** | *Strategy 5:* Annual vaccine 65+ years and “high risk” or those with immunocompromising conditions | | | | | |  |
|  | 18-64 years, low risk | 239 [213, 272] | 12.8 [11.4, 14.6] | - | - | - | 9.9 [-0.6, 21.7] |
|  | 18-64 years, high risk | 746 [696, 803] | 75.7 [70.6, 81.5] | 67.9 [62.3, 73.9] | 39.5 [29.3, 47.3] | 2,531 [2,114, 3,415] | 29.2 [22.8, 34.8] |
|  | 18-64 years, immunocompromised | 425 [389, 477] | 233.4 [213.6, 261.8] | 217.3 [194, 240.9] | 112.7 [74, 146.8] | 887 [681, 1352] | 28.4 [17.2, 36.9] |
|  | 65+ years | 3,452 [3,271, 3,681] | 391.2 [370.6, 417.1] | 376.2 [355.1, 403.6] | 200.6 [175.1, 221.4] | 498 [452, 571] | 32 [28.8, 35.1] |
|  | *Strategy 6:* Semiannual vaccine 65+ years and “high risk” or those with immunocompromising conditions | | | | | |  |
|  | 18-64 years, low risk | 219 [191, 251] | 11.8 [10.3, 13.5] | - | - | - | 16.6 [6.8, 30.7] |
|  | 18-64 years, high risk | 532 [481, 584] | 54.1 [48.9, 59.3] | 41 [35.1, 47.2] | 65.3 [57.7, 74.5] | 1,531 [1,343, 1,734] | 49.2 [43.3, 53.8] |
|  | 18-64 years, immunocompromised | 318 [280, 350] | 174.4 [153.8, 192.2] | 141.9 [116.8, 160.6] | 186.4 [153.9, 220.2] | 536 [454, 650] | 47.2 [40, 53.4] |
|  | 65+ years | 2,172 [2,038, 2,346] | 246.1 [230.9, 265.9] | 222.3 [206.9, 242.8] | 352.9 [333.5, 372.5] | 283 [268, 300] | 57 [54.5, 59.3] |
| **Hybrid** | *Strategy 7:* Annual vaccine 6+ months with second dose in 65+ years and those with immunocompromising conditions | | | | | |  |
|  | 18-64 years, low risk | 171 [151, 202] | 9.2 [8.1, 10.8] | 7.4 [5.8, 9.7] | 6.9 [4.4, 9.6] | 14,498 [10,431, 22,542] | 35.6 [23.2, 46] |
|  | 18-64 years, high risk | 598 [534, 683] | 60.7 [54.2, 69.3] | 54.7 [48.8, 62.9] | 52.3 [42.1, 60.4] | 1,911 [1,655, 2,378] | 43.3 [35.4, 50.4] |
|  | 18-64 years, immunocompromised | 268 [229, 309] | 146.9 [125.7, 169.8] | 115 [92.7, 134.5] | 213.5 [177.4, 249.5] | 468 [401, 564] | 54.9 [47.5, 62.2] |
|  | 65+ years | 1,794 [1,662, 2,011] | 203.3 [188.4, 227.8] | 181.7 [164.6, 204.3] | 396 [373.3, 414.7] | 253 [241, 268] | 64.5 [61.1, 67.4] |
|  | *Strategy 8:* Annual vaccine 18+ years with second dose in 65+ years and those with immunocompromising conditions | | | | | |  |
|  | 18-64 years, low risk | 190 [163, 220] | 10.2 [8.8, 11.8] | 8.2 [6.5, 10.4] | 6.1 [3.8, 8.5] | 16,325 [11,827, 26,135] | 28.2 [16.1, 39.5] |
|  | 18-64 years, high risk | 660 [605, 731] | 67 [61.5, 74.2] | 60.2 [54.8, 66.6] | 46.9 [37.6, 53.9] | 2,133 [1,856, 2,659] | 37.6 [29.6, 43] |
|  | 18-64 years, immunocompromised | 305 [263, 352] | 167.5 [144.7, 193.3] | 133.6 [110.7, 160.2] | 195.6 [154.9, 225.4] | 511 [444, 646] | 49 [39.5, 56.1] |
|  | 65+ years | 2,114 [1,989, 2,338] | 239.5 [225.4, 265] | 215.8 [201.3, 237] | 358.6 [341.4, 378] | 279 [265, 293] | 58.2 [54.9, 60.7] |

##### Specific vaccine coverage for the higher vaccine coverage scenario are in Table S2. Assuming historical uptake timing. Reports median and 95% uncertainty intervals over 100 simulations.

^a^ Absolute risk reduction and NNV compare the no-vaccination reference risk (Table S14) with risk among the vaccinated subgroup.
^b^ Relative risk reduction compares the no-vaccination reference with risk in the entire age-risk group, so it does not equal absolute risk reduction divided by the reference risk.

**Table S17.** **Simulated number of workdays missed in adults 18-64 years due to infection and vaccination under each vaccination eligibility and frequency strategy.**

| Vaccination Strategy | **Historical coverage scenario** | | | **Higher coverage scenario** | | |
| --- | --- | --- | --- | --- | --- | --- |
|  | Workdays missed  due to infection | Workdays missed  due to vaccine side effects | Total workdays  missed | Workdays missed  due to infection | Workdays missed due to vaccine side effects | Total workdays  missed |
| No additional  vaccination during the study period | 1,062,510  [1,027,359, 1,086,832] | 0  [0, 0] | 1,062,510  [1,027,359, 1,086,832] | 1,062,510  [1,027,359, 1,086,832] | 0  [0, 0] | 1,062,510  [1,027,359, 1,086,832] |
| Strategy 1  (Universal, 6mo+) | 990,335  [959,852, 1,025,784] | 37,953  [37,880, 38,018] | 1,028,285  [997,838, 1,063,706] | 736,411  [694,799, 828,917] | 144,093  [143,963, 144,216] | 880,580  [838,925, 973,012] |
| Strategy 2  (Universal, 18y+) | 993,891  [964,226, 1,025,339] | 37,953  [37,880, 38,018] | 1,031,908  [1,002,200, 1,063,284] | 812,106  [782,599, 876,221] | 144,093  [143,963, 144,216] | 956,211  [926,766, 1,020,267] |
| Strategy 3  (Annual  risk-based traditional) | 1,046,775  [1,012,372, 1,071,762] | 3,477  [3,459, 3,484] | 1,050,245  [1,015,852, 1,075,236] | 1,024,260  [992,990, 1,051,709] | 11,844  [11,814, 11,861] | 1,036,091  [1,004,838, 1,063,558] |
| Strategy 4  (Semiannual risk-based traditional) | 1,044,894  [1,011,498, 1,070,070] | 5,220  [5,202, 5,229] | 1,050,117  [1,016,713, 1,075,274] | 996,556  [965,265, 1,023,067] | 35,885  [35,849, 35,898] | 1,032,443  [1,001,155, 1,058,937] |
| Strategy 5  (Annual  risk-based expanded) | 1,018,844  [987,117, 1,046,955] | 22,209  [22,198, 22,276] | 1,041,082  [1,009,324, 1,069,193] | 925,079  [897,565, 964,044] | 75,734  [75,693, 75,812] | 1,000,819  [973,327, 1,039,770] |
| Strategy 6  (Semiannual risk-based expanded) | 1,013,986  [982,231, 1,040,709] | 33,381  [33,363, 33,450] | 1,047,395  [1,015,658, 1,074,083] | 820,174  [794,457, 858,033] | 229,806  [229,758, 229,880] | 1,049,983  [1,024,304, 1,087,831] |
| Strategy 7  (Hybrid,  6mo+) | 988,430  [958,118, 1,024,146] | 39,696  [39,624, 39,765] | 1,028,095  [997,838, 1,063,815] | 705,268  [666,522, 793,125] | 168,137  [168,014, 168,243] | 873,388  [834,705, 961,227] |
| Strategy 8  (Hybrid,  18y+) | 994,688  [962,920, 1,024,461] | 39,694  [39,628, 39,765] | 1,034,368  [1,002,644, 1,064,120] | 779,197  [750,026, 837,836] | 168,137  [168,014, 168,243] | 947,268  [918,167, 1,005,916] |

Corresponding to a total population of 5 million people. Reporting the median and 95% uncertainty interval. Historical and higher vaccine coverage scenarios specified in Table S2.

**Table S18: Statistical comparison of July universal annual vaccination with hybrid and risk-based semiannual vaccination at historical fall timing.**

| Age-risk group | Population | Most favorable strategy (lowest median hospitalization risk) | Superior in majority of runs vs each other month ^a^ | Significant pairwise comparisons ^b^ |
| --- | --- | --- | --- | --- |
| 18-64 years, low risk | Total | July annual universal | Yes | 2/2 |
|  | Vaccinated* | July annual universal | No | 1/2 |
| 18-64 years, high risk | Total | July annual universal | Yes | 2/2 |
|  | Vaccinated* | July annual universal | No | 1/2 |
| 18-64 years, immunocompromised | Total | July annual universal | Yes | 2/2 |
|  | Vaccinated* | Risk-based semiannual | Yes | 1/2 |
| 65+ years | Total | July annual universal | Yes | 2/2 |
|  | Vaccinated* | Hybrid | Yes | 2/2 |

Corresponds to results from Tables S5 (July universal annual) and S14 (hybrid and risk-based semiannual with historical timing).

^a^ indicates whether the most favorable month had lower risk in >50% of pairwise comparisons, compared to each of the six other months.

^b^ the number of other months for which the most favorable month had significantly lower hospitalization risk, by a Holm-corrected paired Wilcoxon test (p < 0.05).

*Note that the vaccinated subgroup is defined as people who received the max number of doses they are eligible for during the study period, so the subgroup changes in size across these three strategies (e.g., for the 65+ years and immunocompromised adults 18-64 years, the vaccinated subgroup under hybrid and risk-based semiannual strategies refers to those who received two doses, while July annual universal refers to those who received one dose.)

**Table S19: Median paired differences in annual hospitalization risk (per 100,00 persons) pairwise between three strategies: July universal annual vaccination, hybrid vaccination with historical timing, and risk-based semiannual vaccination with historical timing.**

| Age-risk group | July universal vs hybrid | | July universal vs semiannual risk-based | | Hybrid vs semiannual risk-based | |
| --- | --- | --- | --- | --- | --- | --- |
|  | Difference in hospitalization risk  (per 100,000) | Lower risk in X percent of matched simulations | Difference in hospitalization risk  (per 100,000) | Lower risk in X percent of matched simulations | Difference in hospitalization risk  (per 100,000) | Lower risk in X percent of matched simulations |
| 18-64 years, low risk | 0.6  [-1.2, 2.4] | 71% | 1.3  [-1.2, 3.4] | 91% | 0.7  [-1.4, 3] | 74% |
| 18-64 years, high risk | 3.2  [-6.3, 13] | 74% | 13.8  [4.3, 21.6] | 100% | 10.5  [0.5, 20] | 97% |
| 18-64 years, immunocompromised | 9.3  [-22.8, 51.9] | 80% | 20.9  [-8.9, 53] | 91% | 13.2  [-24, 46.2] | 72% |
| ≥65 years | 19.9  [0.7, 40.1] | 97% | 36.1  [14.3, 54.1] | 100% | 16.7  [-6.3, 34.2] | 92% |

Hospitalization risk corresponds to cumulative hospitalization incidence over the 12-month study period. Differences were computed within each matched simulation (paired on seed and parameter set) before summarizing. Difference in hospitalization risk reports the median with the 2.5th to 97.5th percentiles of the paired differences in brackets. Positive values indicate lower risk under the first-named strategy. The lower risk in X percent of matched simulations is the percentage that the first-named strategy had lower hospitalization risk than the second-named strategy. July universal annual vaccination assumes one-month uptake (Table S5); hybrid and semiannual risk-based vaccination assume historical fall timing (Table S14). Statistical comparisons are reported separately in Table S18.

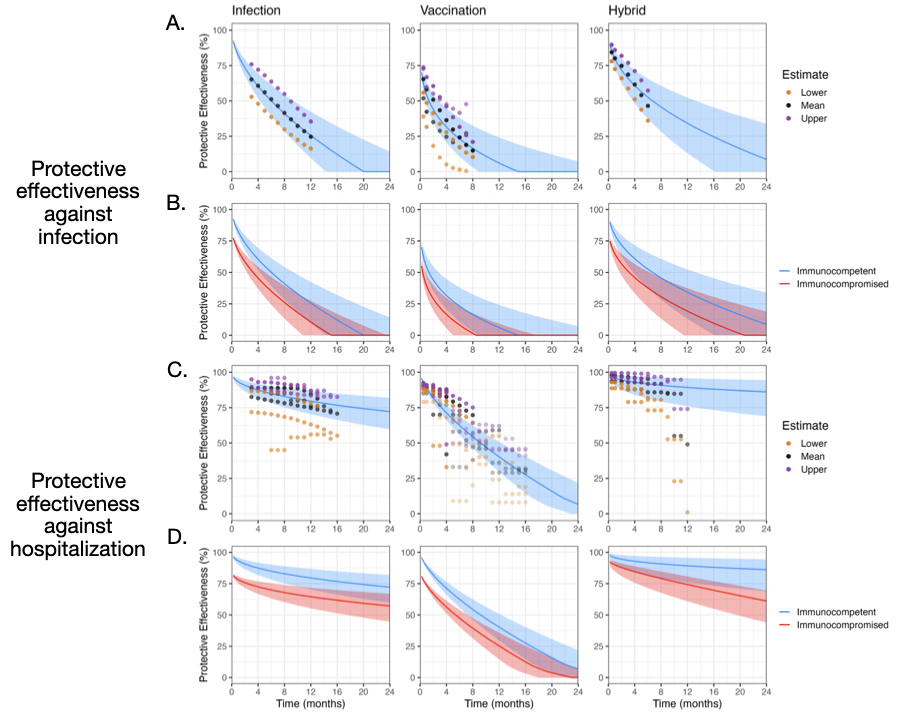

Figure S1: **Modeled waning of protective effectiveness against SARS-CoV-2 infection and COVID-19 hospitalization, by immunity type, outcome, and risk group.** Curves show absolute protective effectiveness relative to an immunologically naïve baseline over 24 months since the most recent immune event. Columns indicate immunity type: infection-acquired (left), vaccine-acquired (middle), and hybrid, i.e. both prior infection and vaccination (right). (A, C) Literature estimates (Table S3) and fitted waning curves for infection (A) and hospitalization (C). (B, D) Waning curves for immunocompetent (blue) and immunocompromised (red) individuals, for infection (B) and hospitalization (D). Curves were fitted by linear mixed-effects models, with central curves fitted to reported means and shaded regions to reported lower and upper 95% confidence limits. Assumed waning curves for immunocompromised persons were 15 percentage points lower than immunocompetent. See Section S2 for more details.

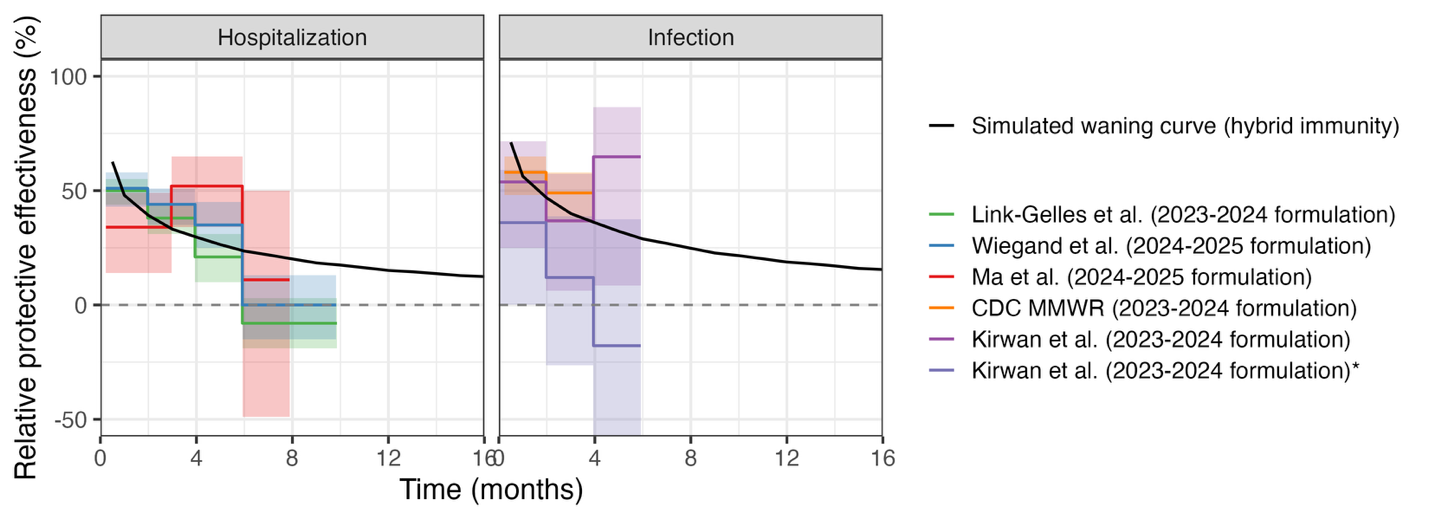

Figure S2: **Validation analysis comparing modeled and literature-based relative vaccine effectiveness of an additional COVID-19 dose by prior infection status**. Model predictions (black lines) show the relative vaccine effectiveness of an additional COVID-19 vaccine dose over 24 months, stratified by no prior infection (left) and prior infection (right) and outcome (top: hospitalization; bottom: infection). Models assume an average time of ~7 months since the last immune event. Colored lines represent empirical estimates from published studies, with some studies stratifying by prior infection status and others providing aggregate estimates. All effectiveness values are relative to no additional dose during the study period.

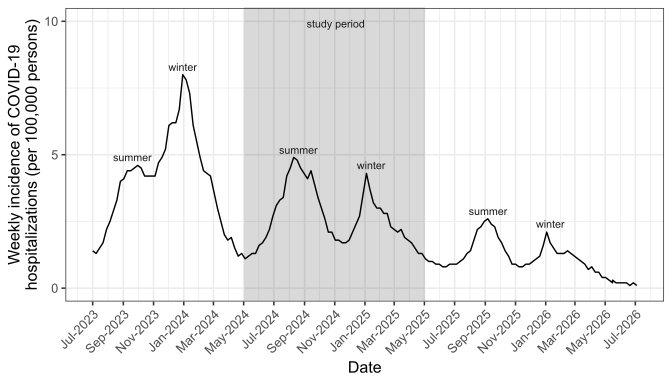

**Figure S3: Weekly COVID-19 hospitalization incidence in the United States, July 2023–June 2026.** Rate per 100,000 persons. Study period is indicated by grey shading. Source: COVID-Net surveillance system, accessed July 12, 2026^1^.

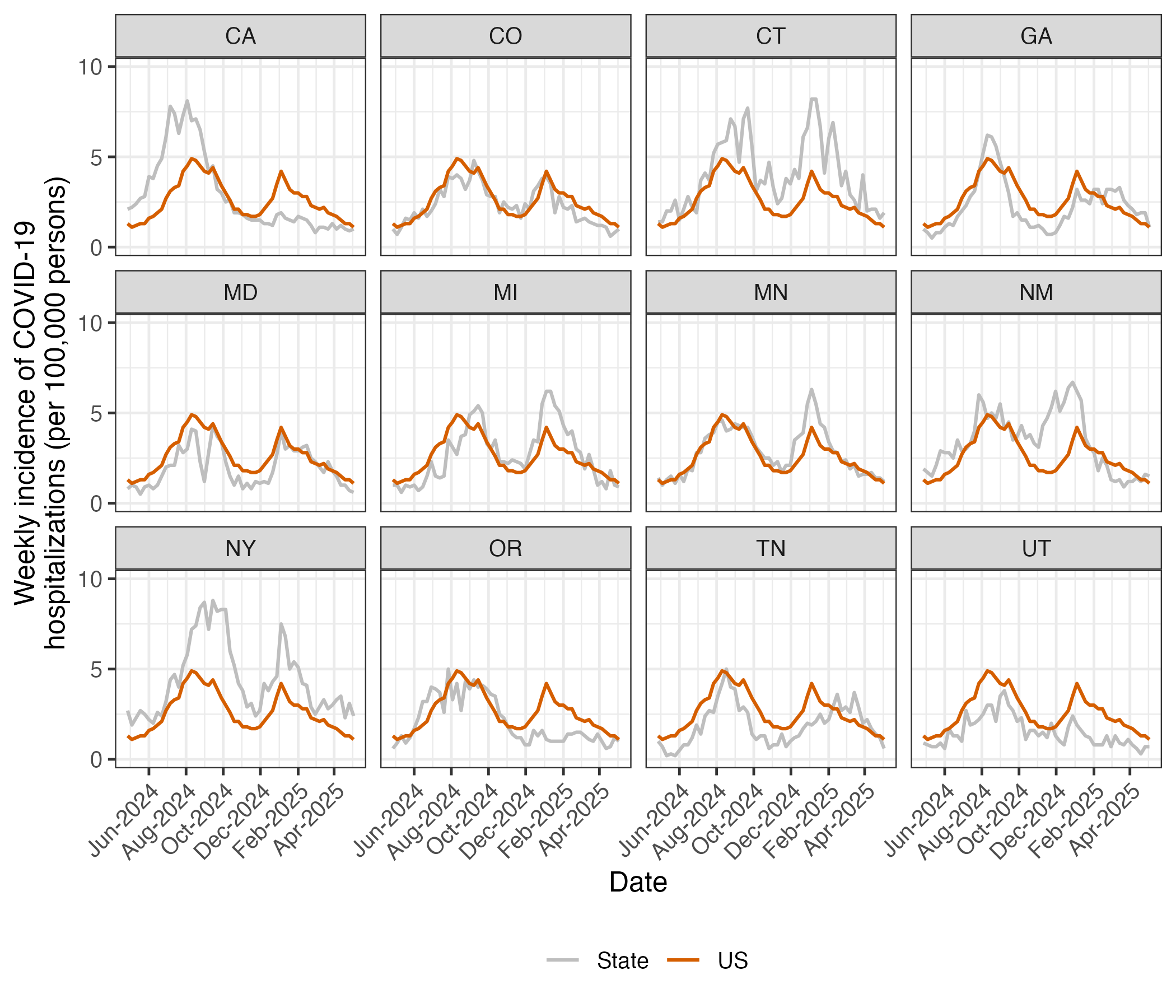

**Figure S4: State-level COVID-19 hospitalization incidence compared to national rates, May 2024–April 2025.** Weekly incidence per 100,000 persons for 12 states with reporting data from the COVID-NET database^1^. Gray lines show state-specific rates; orange line shows US national rate.

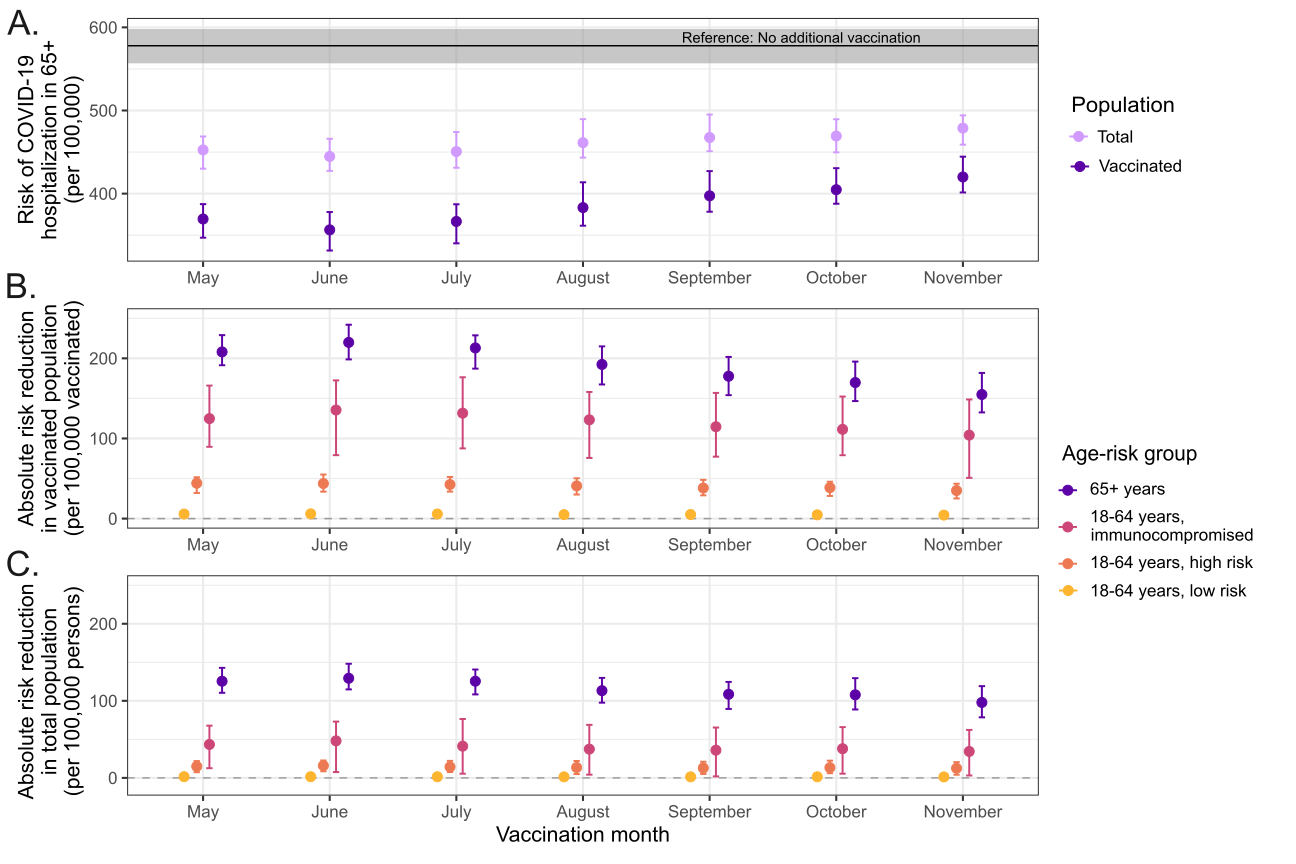

**Figure S5: Impact of vaccination timing under historical 4-month vaccination uptake, evaluating cumulative hospitalization risk over 12-month study period.** We modeled universal vaccination in individuals ≥6 months of age using the observed 2024-2025 vaccination uptake pattern (distributed over four months (Dose 2 curves in September-December, Figure 1B)). To evaluate different start times, we shifted this entire uptake distribution to begin in each labeled vaccination month (x-axis) and assessed cumulative hospitalization risk over the one-year study period. (A) Annual hospitalization risk per 100,000 in adults ≥65 years, shown for vaccinated individuals (dark purple), the total population including unvaccinated (light purple), and a no-vaccination scenario (gray). (B) Absolute risk reduction per 100,000 among vaccinated individuals and (C) the total population, stratified by age-risk group: ≥65 years (purple), 18–64 years immunocompromised (pink), 18–64 years with comorbidities (orange), and 18–64 years low risk (yellow). Error bars represent 95% uncertainty intervals (i.e., 2.5^th^-97.5^th^ range of 100 stochastic simulations).

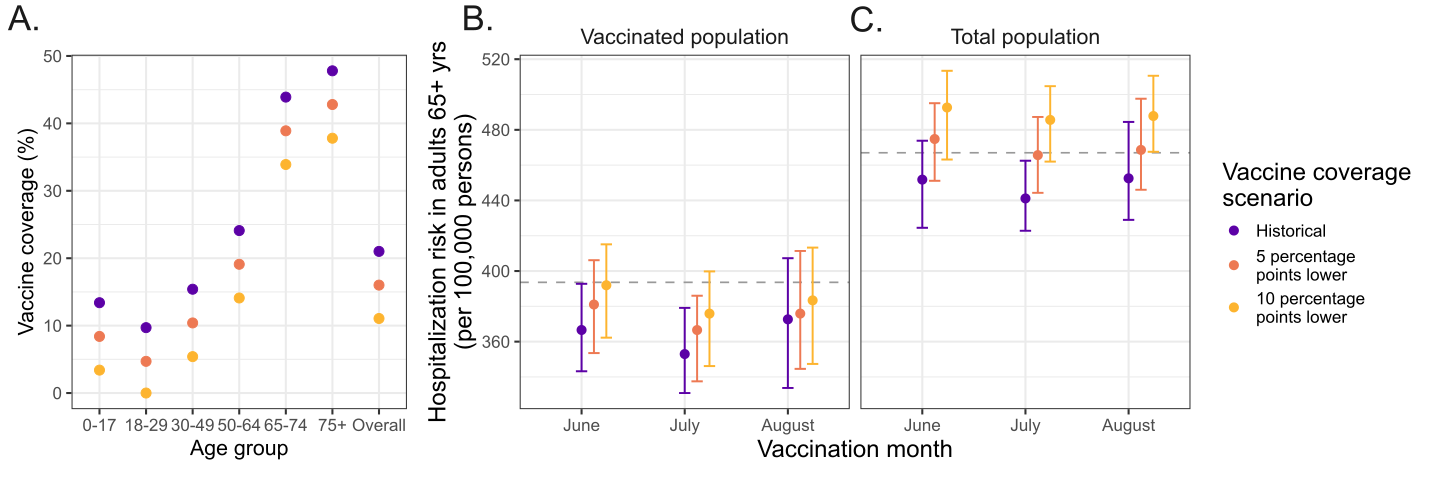

**Figure S6: Sensitivity of summer vaccination timing to reduced vaccine coverage.**

(A) Vaccine coverage (%) by age group and overall, under three vaccine coverage scenarios: historical (purple), 5 absolute percentage points lower (orange), and 10 absolute percentage points lower (yellow). (B-C) Annual COVID-19 hospitalization risk among vaccinated adults ≥65 years (B, per 100,000 persons vaccinated) and all adults among ≥65 years (C, per 100,000 persons), by vaccination month (x-axis) under the three coverage scenarios, assuming annual universal vaccination (≥6 months) with coverage achieved over one month. Gray dashed lines show risk under annual universal vaccination with historical coverage and uptake as a reference (closely matching risk under annual universal vaccination with one-month uptake in September). Points show medians with 95% uncertainty intervals.

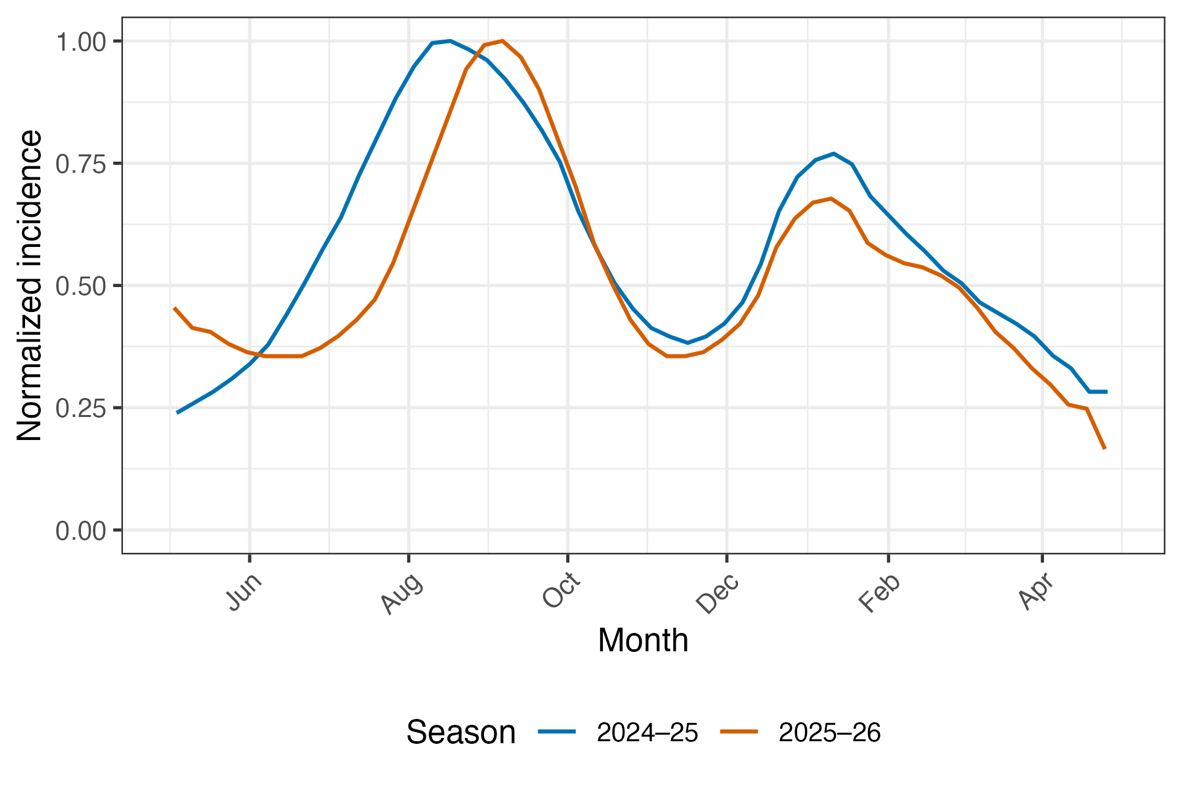

**Figure S7: Comparison of normalized national US COVID-19 hospitalization trends during the study period (2024-25) and the latest season (2025-26).** Curves are smoothed for visualization using a 5-week centered moving average. The ratio of cumulative hospitalization burden during May-October versus November-April was 1.28 (2024-25) and 1.31 (2025-2026).

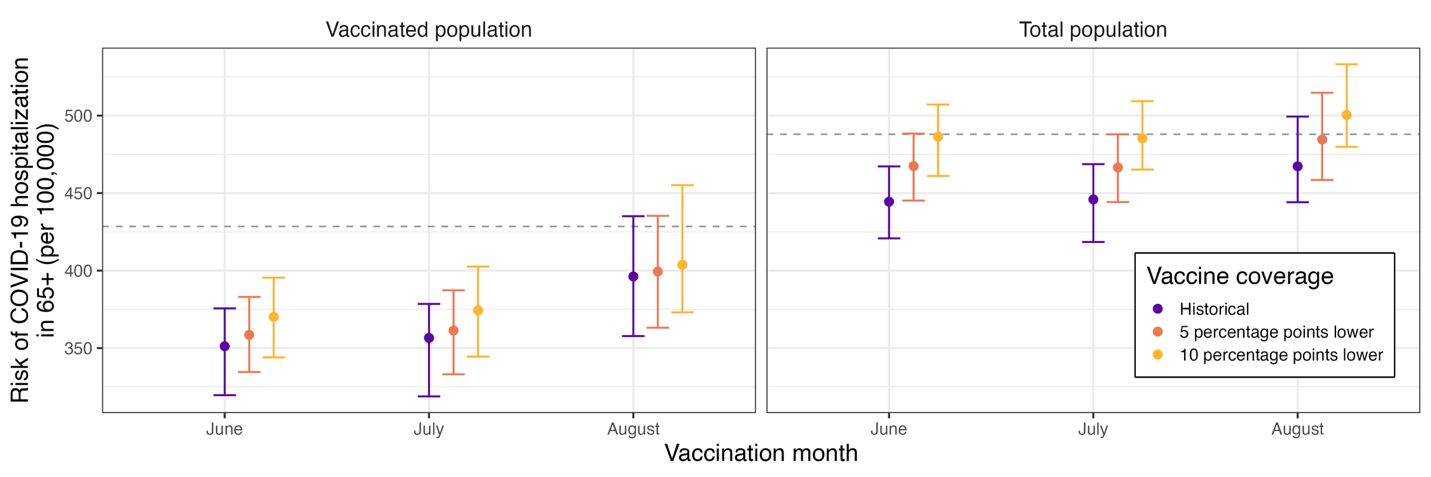

**Figure S8: Optimal vaccination timing for Archetype 1 under reduced summer coverage scenarios.** COVID-19 hospitalization risk per 100,000 adults ≥65 years by vaccination month under the three coverage scenarios: historical (purple), 5 absolute percentage points lower in each age-risk group in June–August (orange), and 10 absolute percentage points lower (yellow), showing risk among vaccinated individuals (left) and total population (right). Gray dashed line show annual universal vaccination with uptake in September as a reference. Points show medians with 95% uncertainty intervals.

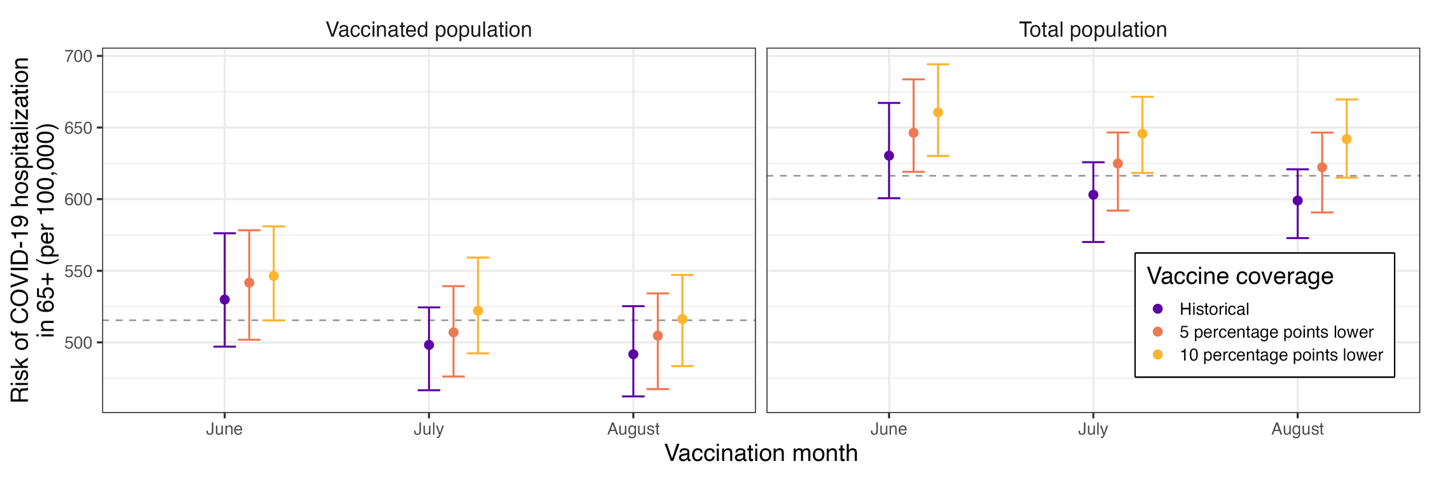

**Figure S9: Optimal vaccination timing for Archetype 2 under reduced summer coverage scenarios.** COVID-19 hospitalization risk per 100,000 adults ≥65 years by vaccination month under the three coverage scenarios: historical (purple), 5 absolute percentage points lower in each age-risk group in June–August (orange), and 10 absolute percentage points lower (yellow), showing risk among vaccinated individuals (left) and total population (right). Gray dashed line show annual universal vaccination with uptake in September as a reference. Points show medians with 95% uncertainty intervals.

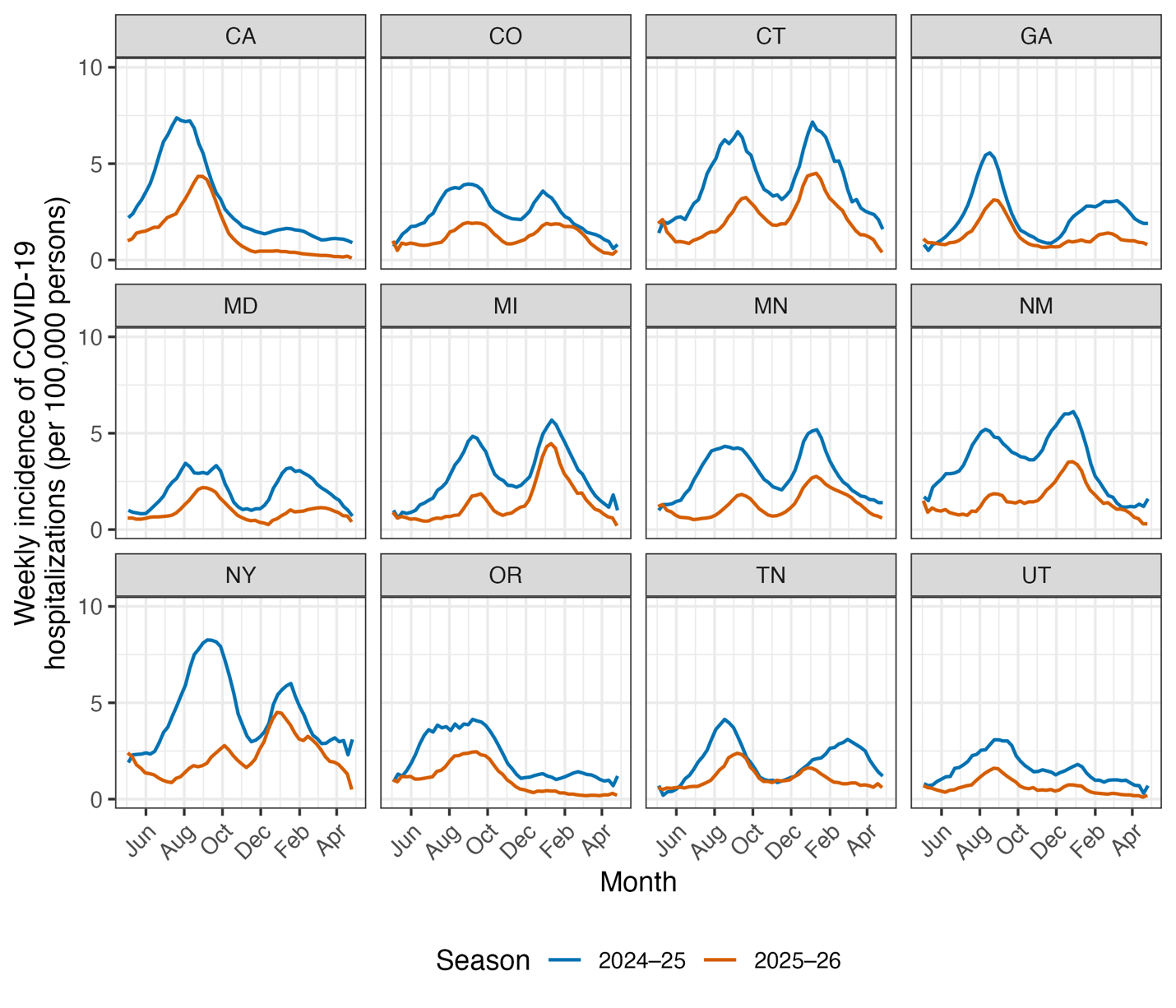

##### Figure S10: Comparison of state-level COVID-19 hospitalization trends during the study period 2024-25 and the latest 2025-26 season. Curves are smoothed for visualization using a 5-week centered moving average.

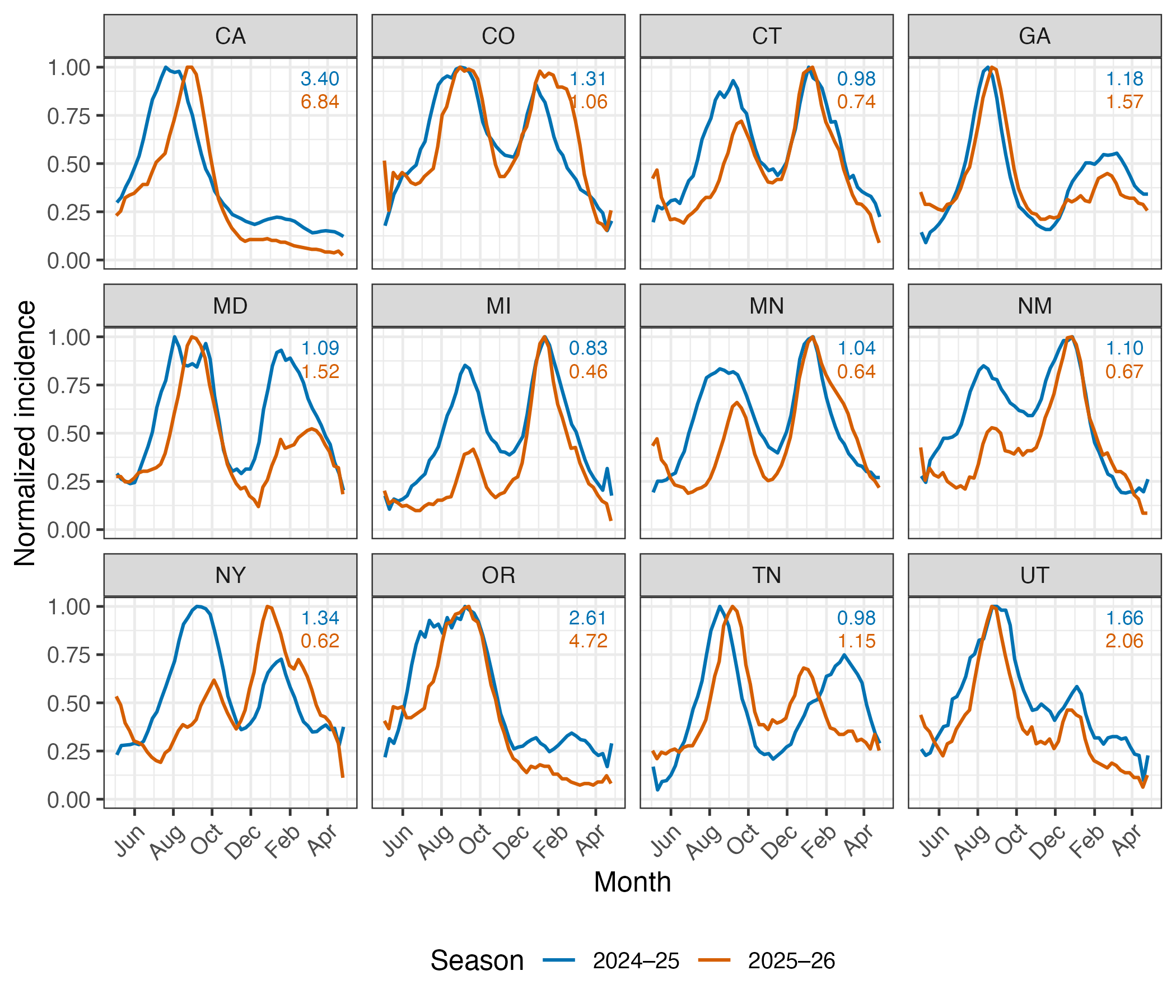

##### Figure S11: Comparison of normalized state-level COVID-19 hospitalization trends during the study period 2024-25 and the latest 2025-26 season. Curves are smoothed for visualization using a 5-week centered moving average. Annotations indicate the ratio of cumulative hospitalization burden occurring in the May-October versus November-April in each year, with the color corresponding to the season: 2024-25 (blue), 2025-26 (orange).

### References

1.	Centers for Disease Control and Prevention. Coronavirus Disease 2019 (COVID-19) Hospitalization Surveillance Network (COVID-NET). Published online January 30, 2026. Accessed June 24, 2026. https://www.cdc.gov/covid/php/covid-net/index.html

2.	Centers for Disease Control and Prevention. Patient Characteristics of Laboratory-Confirmed COVID-19 Hospitalizations from the COVID-NET Surveillance System. Accessed June 24, 2026. https://data.cdc.gov/Public-Health-Surveillance/Patient-Characteristics-of-Laboratory-Confirmed-CO/bigw-pgk2/about_data

3.	Herrera-Esposito D, de los Campos G. Age-specific rate of severe and critical SARS-CoV-2 infections estimated with multi-country seroprevalence studies. *BMC Infect Dis*. 2022;22:311. doi:10.1186/s12879-022-07262-0

4.	Mossadeghi B, Caixeta R, Ondarsuhu D, Luciani S, Hambleton I, Hennis AJM. Multimorbidity and social determinants of health in the US prior to the COVID-19 pandemic and implications for health outcomes: a cross-sectional analysis based on NHANES 2017–2018. *BMC Public Health*. 2023;23(887). doi:10.1186/s12889-023-15768-8

5.	Jindai K. Multimorbidity and Functional Limitations Among Adults 65 or Older, NHANES 2005–2012. *Prev Chronic Dis*. 2016;13. doi:10.5888/pcd13.160174

6.	Boersma P, Black LI, Ward BW. Prevalence of Multiple Chronic Conditions Among US Adults, 2018. *Prev Chronic Dis*. 2020;17:E106. doi:10.5888/pcd17.200130

7.	Centers for Disease Control and Prevention. Chronic Disease Indicator (CDI). National Center for Chronic Disease Prevention and Health Promotion. Accessed June 24, 2026. https://cdi.cdc.gov/?location=US&category=HEA&indicators=HEA05

8.	Centers for Disease Control and Prevention. People Who Are Immunocompromised. Centers for Disease Control and Prevention. February 11, 2020. Accessed July 18, 2026. https://archive.cdc.gov/www_cdc_gov/coronavirus/2019-ncov/need-extra-precautions/people-who-are-immunocompromised.html

9.	Taylor CA. COVID-19–Associated Hospitalizations Update — COVID-NET, July 2023–September 2024. Slide presentation presented at: Meeting of the Advisory Committee on Immunization Practices; October 23, 2024; Atlanta, GA. Accessed September 14, 2026. https://www.cdc.gov/acip/downloads/slides-2024-10-23-24/03-COVID-Taylor-508.pdf

10.	Park HJ, Gonsalves GS, Tan ST, et al. Comparing frequency of booster vaccination to prevent severe COVID-19 by risk group in the United States. *Nat Commun*. 2024;15(1):1883. doi:10.1038/s41467-024-45549-9

11.	Ferdinands JM, Rao S, Dixon BE, et al. Waning of vaccine effectiveness against moderate and severe covid-19 among adults in the US from the VISION network: test negative, case-control study. *BMJ*. 2022;379:e072141. doi:10.1136/bmj-2022-072141

12.	Britton A. Effectiveness of COVID-19 mRNA Vaccines Against COVID-19–Associated Hospitalizations Among Immunocompromised Adults During SARS-CoV-2 Omicron Predominance — VISION Network, 10 States, December 2021—August 2022. *MMWR Morb Mortal Wkly Rep*. 2022;71. doi:10.15585/mmwr.mm7142a4

13.	Wu Y, Guo Z, Yuan J, et al. Duration of viable virus shedding and polymerase chain reaction positivity of the SARS-CoV-2 Omicron variant in the upper respiratory tract: a systematic review and meta-analysis. *Int J Infect Dis*. 2023;129:228-235. doi:10.1016/j.ijid.2023.02.011

14.	Li Y, Jiang X, Qiu Y, et al. Latent and incubation periods of Delta, BA.1, and BA.2 variant cases and associated factors: a cross-sectional study in China. *BMC Infect Dis*. 2024;24(1):294. doi:10.1186/s12879-024-09158-7

15.	National Center for Immunization and Respiratory Diseases (U.S.) Division of Viral Diseases. Reinfections and COVID 19. September 9, 2022. Accessed August 24, 2026. https://stacks.cdc.gov/view/cdc/121105

16.	Prem K, Cook AR, Jit M. Projecting social contact matrices in 152 countries using contact surveys and demographic data. *PLOS Comput Biol*. 2017;13(9):e1005697. doi:10.1371/journal.pcbi.1005697

17.	Haynes JM, Dodd RY, Crowder LA, Notari EP, Stramer SL. Trajectory and Demographic Correlates of Antibodies to SARS-CoV-2 Nucleocapsid in Recently Infected Blood Donors, United States. *Emerg Infect Dis*. 2023;29(7). doi:10.3201/eid2907.230173

18.	Klaassen F, Chitwood MH, Cohen T, et al. Changes in Population Immunity Against Infection and Severe Disease From Severe Acute Respiratory Syndrome Coronavirus 2 Omicron Variants in the United States Between December 2021 and November 2022. *Clin Infect Dis*. 2023;77(3):355-361. doi:10.1093/cid/ciad210

19.	Centers for Disease Control and Prevention. Antibody Seroprevalence. COVID-19. September 5, 2025. Accessed August 13, 2026. https://www.cdc.gov/covid/php/antibody-seroprevalence/index.html

20.	Centers for Disease Control and Prevention. 2022–2023 Nationwide Blood Donor Seroprevalence Survey Combined Infection- and Vaccination-Induced Seroprevalence Estimates. Accessed August 13, 2026. https://data.cdc.gov/Laboratory-Surveillance/2022-2023-Nationwide-Blood-Donor-Seroprevalence-Su/ar8q-3jhn/data_preview

21.	Centers for Disease Control and Prevention. COVID-19 Vaccination Age and Sex Trends in the United States, National and Jurisdictional. Published online May 11, 2023. Accessed June 24, 2026. https://data.cdc.gov/Vaccinations/COVID-19-Vaccination-Age-and-Sex-Trends-in-the-Uni/5i5k-6cmh/about_data

22.	Del Moral P, Doucet A, Jasra A. An adaptive sequential Monte Carlo method for approximate Bayesian computation. *Stat Comput*. 2012;22(5):1009-1020. doi:10.1007/s11222-011-9271-y

23.	Jabot F, Faure T, Dumoulin N. EasyABC: performing efficient approximate Bayesian computation sampling schemes using R. *Methods Ecol Evol*. 2013;4(7):684-687. doi:10.1111/2041-210X.12050

24.	Yu W, Guo Y, Zhang S, Kong Y, Shen Z, Zhang J. Proportion of asymptomatic infection and nonsevere disease caused by SARS-CoV-2 Omicron variant: A systematic review and analysis. *J Med Virol*. 2022;94(12):5790-5801. doi:10.1002/jmv.28066

25.	Fitzpatrick MC, Moghadas SM, Vilches TN, Shah A, Pandey A, Galvani AP. Estimated US Pediatric Hospitalizations and School Absenteeism Associated With Accelerated COVID-19 Bivalent Booster Vaccination. *JAMA Netw Open*. 2023;6(5):e2313586. doi:10.1001/jamanetworkopen.2023.13586

26.	What is the unemployment rate in the US right now? USAFacts. July 2, 2026. Accessed July 10, 2026. https://usafacts.org/answers/what-is-the-unemployment-rate/country/united-states/

27.	Wang Y, So HC, Tsang NNY, et al. Clinical profile analysis of SARS-CoV-2 community infections during periods with omicron BA.2, BA.4/5, and XBB dominance in Hong Kong: a prospective cohort study. *Lancet Infect Dis*. 2025;25(3):276-289. doi:10.1016/S1473-3099(24)00574-7

28.	Maltezou HC, Gamaletsou MN, Lourida A, et al. Comparison of morbidity and absenteeism due to COVID-19 and seasonal influenza in a large cohort of health care personnel in the 2022 to 2023 season. *Am J Infect Control*. 2024;52(11):1248-1251. doi:10.1016/j.ajic.2024.05.015

29.	Choi M, Kim J, Kim H, Tobin RJ, Lee S. Multiscale Modeling of Hospital Length of Stay for Successive SARS-CoV-2 Variants: A Multi-State Forecasting Framework. *Viruses*. 2025;17(7):953. doi:10.3390/v17070953

30.	Domen J, Abrams S, Digregorio M, et al. Predictors of moderate-to-severe side-effects following COVID-19 mRNA booster vaccination: a prospective cohort study among primary health care providers in Belgium. *BMC Infect Dis*. 2024;24(1):1135. doi:10.1186/s12879-024-09969-8

31.	Politis M, Rachiotis G, Mouchtouri VA, Hadjichristodoulou C. The Global Burden of Absenteeism Related to COVID-19 Vaccine Side Effects Among Healthcare Workers: A Systematic Review and Meta-Analysis. *Vaccines*. 2024;12(10):1196. doi:10.3390/vaccines12101196

32.	United States Census Bureau. U.S. and World Population Clock: Population by Age and Sex. United States Census Bureau. Accessed June 24, 2026. https://www.census.gov/popclock/data_tables.php?component=pyramid

33.	Martinson ML, Lapham J. Prevalence of Immunosuppression Among US Adults. *JAMA*. 2024;331(10):880-882. doi:10.1001/jama.2023.28019

34.	Rezaee ME. Multiple Chronic Conditions Among Outpatient Pediatric Patients, Southeastern Michigan, 2008–2013. *Prev Chronic Dis*. 2015;12. doi:10.5888/pcd12.140397

35.	Patel M, Chen J, Kim S, et al. Analysis of MarketScan Data for Immunosuppressive Conditions and Hospitalizations for Acute Respiratory Illness, United States. *Emerg Infect Dis*. 2020;26(8). doi:10.3201/eid2608.191493

36.	Centers for Disease Control and Prevention. COVID-19 Vaccination Coverage, Overall and by Selected Demographics and Jurisdiction, Among Adults 18 Years and Older, by Season. Published online March 4, 2026. Accessed July 3, 2026. https://data.cdc.gov/Vaccinations/COVID-19-Vaccination-Coverage-Overall-and-by-Selec/ksfb-ug5d/about_data

37.	COVID-19 Vaccination Implementation. Accessed July 14, 2026. https://www.cdc.gov/acip/downloads/slides-2025-06-25-26/05-Peacock-COVID-508.pdf

38.	Centers for Disease Control and Prevention. Weekly Parental Intent for Vaccination and Cumulative Percentage of Children 6 Months-17 Years Who are Up to date with the COVID-19 Vaccines by Season, United States. Published online July 2, 2026. Accessed July 3, 2026. https://data.cdc.gov/Child-Vaccinations/Weekly-Parental-Intent-for-Vaccination-and-Cumulat/ker6-gs6z/data_preview

39.	Centers for Disease Control and Prevention. National Immunization Survey Adult COVID Module (NIS-ACM). Accessed July 8, 2026. https://data.cdc.gov/Vaccinations/National-Immunization-Survey-Adult-COVID-Module-NI/si7g-c2bs/data_preview

40.	Bobrovitz N, Ware H, Ma X, et al. Protective effectiveness of previous SARS-CoV-2 infection and hybrid immunity against the omicron variant and severe disease: a systematic review and meta-regression. *Lancet Infect Dis*. 2023;23(5):556-567. doi:10.1016/S1473-3099(22)00801-5

41.	Carazo S, Skowronski DM, Brisson M, et al. Effectiveness of previous infection-induced and vaccine-induced protection against hospitalisation due to omicron BA subvariants in older adults: a test-negative, case-control study in Quebec, Canada. *Lancet Healthy Longev*. 2023;4(8):e409-e420. doi:10.1016/S2666-7568(23)00099-5

42.	Lee N, Nguyen L, Austin PC, et al. Protection Conferred by COVID-19 Vaccination, Prior SARS-CoV-2 Infection, or Hybrid Immunity Against Omicron-Associated Severe Outcomes Among Community-Dwelling Adults. *Clin Infect Dis*. 2024;78(5):1372-1382. doi:10.1093/cid/ciad716

43.	Link-Gelles R, Levy ME, Natarajan K, et al. Estimation of COVID-19 mRNA Vaccine Effectiveness and COVID-19 Illness and Severity by Vaccination Status During Omicron BA.4 and BA.5 Sublineage Periods. *JAMA Netw Open*. 2023;6(3):e232598. doi:10.1001/jamanetworkopen.2023.2598

44.	Stein C, Nassereldine H, Sorensen RJD, et al. Past SARS-CoV-2 infection protection against re-infection: a systematic review and meta-analysis. *The Lancet*. 2023;401(10379):833-842. doi:10.1016/S0140-6736(22)02465-5

45.	Link-Gelles R. Effectiveness of COVID-19 vaccines. Slide presentation presented at: Meeting of the Advisory Committee on Immunization Practices; October 23, 2024; Atlanta, GA. https://www.cdc.gov/acip/downloads/slides-2024-10-23-24/04-COVID-Link-Gelles-508.pdf?exitCode=pfa

46.	Wiegand RE, Payne AB, Mak J, et al. Estimated Effectiveness of 2024-2025 COVID-19 Vaccines in Adults. *JAMA Intern Med*. Published online June 15, 2026. doi:10.1001/jamainternmed.2026.1936

47.	Ma KC, Webber A, Lauring AS, et al. Estimated Effectiveness of 2024-2025 COVID-19 Vaccination Against Severe COVID-19. *JAMA Netw Open*. 2026;9(2):e2557415. doi:10.1001/jamanetworkopen.2025.57415

48.	Lau JJ, Cheng SMS, Leung K, et al. Real-world COVID-19 vaccine effectiveness against the Omicron BA.2 variant in a SARS-CoV-2 infection-naive population. *Nat Med*. 2023;29(2):348-357. doi:10.1038/s41591-023-02219-5

49.	Ruth Link-Gelles P, Allison Avrich Ciesla P, Josephine Mak MPH, et al. Early Estimates of Updated 2023–2024 (Monovalent XBB.1.5) COVID-19 Vaccine Effectiveness Against Symptomatic SARS-CoV-2 Infection Attributable to Co-Circulating Omicron Variants Among Immunocompetent Adults — Increasing Community Access to Testing Program, United States, September 2023–January 2024. *MMWR Morb Mortal Wkly Rep*. 2024;73. doi:10.15585/mmwr.mm7304a2

50.	Kirwan PD, Foulkes S, Munro K, et al. Protection of vaccine boosters and prior infection against mild/asymptomatic and moderate COVID-19 infection in the UK SIREN healthcare worker cohort: October 2023 to March 2024. *J Infect*. 2024;89(5):106293. doi:10.1016/j.jinf.2024.106293
